# A randomised controlled pilot trial of oral 11β-HSD1 inhibitor AZD4017 for wound healing in adults with type 2 diabetes mellitus

**DOI:** 10.1101/2021.03.23.21254200

**Authors:** Ramzi Ajjan, Elizabeth MA Hensor, Kave Shams, Francesco Del Galdo, Afroze Abbas, Janet Woods, Rebecca J Fairclough, Lorraine Webber, Lindsay Pegg, Adrian Freeman, Ann Morgan, Paul M Stewart, Angela E Taylor, Wiebke Arlt, Abd Tahrani, David Russell, Ana Tiganescu

**Affiliations:** Leeds Institute of Cardiovascular and Metabolic Medicine, University of Leeds, Leeds, UK; Leeds Institute of Rheumatic and Musculoskeletal Medicine, University of Leeds, Leeds, UK; NIHR Biomedical Research Center, Leeds Teaching Hospitals, NHS Trust, Leeds, UK; Leeds Centre for Dermatology, Chapel Allerton Hospital, Leeds, UK; Leeds Vascular Institute, Leeds Teaching Hospitals NHS Trust, Leeds, UK; Emerging Innovations Unit, Discovery Sciences, BioPharmaceuticals R&D, AstraZeneca, Cambridge, UK; Emerging portfolio development, Late Oncology, Oncology R&D, AstraZeneca, Cambridge, UK; Faculty of Medicine and Health, University of Leeds, Leeds, UK; Institute of Metabolism and Systems Research (IMSR), University of Birmingham, Birmingham, UK; Centre of Endocrinology Diabetes and Metabolism (CEDAM), Birmingham Health partners, Birmingham, UK; Department of Diabetes and Endocrinology, University Hospitals Birmingham NIHS Foundation Trust, Birmingham, UK

**Keywords:** Glucocorticoid, Skin, Wound healing, 11 beta hydroxysteroid dehydrogenase type 1, Type 2 diabetes mellitus, Randomised controlled trial

## Abstract

Chronic wounds (e.g. diabetic foot ulcers) have a major impact on quality of life, yet treatments remain limited. Glucocorticoids impair wound healing; preclinical research suggests that blocking glucocorticoid activation by the enzyme 11β-hydroxysteroid dehydrogenase type 1 (11β-HSD1) improves wound repair. This investigator-initiated double-blind, randomised, placebo-controlled parallel-group phase 2b pilot trial investigated efficacy, safety and feasibility of 11β-HSD1 inhibition for 35 days by oral AZD4017 (AZD) treatment in adults with type 2 diabetes (n=14) compared to placebo (PCB, n=14) in a single-centre secondary care setting. Computer-generated 1:1 randomisation was pharmacy-administered. From 300 screening invitations, 36 attended, 28 were randomised. There was no proof-of-concept that AZD inhibited 24 hour skin 11β-HSD1 activity at day 28 (primary outcome: adjusted difference AZD-PCB 90% CI (diffCI)=-3.4,5.5) but systemic 11β-HSD1 activity (median urinary [THF+alloTHF]/THE ratio) was 87% lower with AZD at day 35 (PCB 1.00, AZD 0.13, diffCI=-1.04,-0.69). Mean wound gap diameter (mm) following baseline 2mm punch biopsy was 34% smaller at day 2 (PCB 1.51, AZD 0.98, diffCI=-0.95,-0.10) and 48% smaller after repeat wounding at day 30 (PCB 1.35, AZD 0.70, diffCI=-1.15,-0.16); results also suggested greater epidermal integrity but modestly impaired barrier function with AZD. AZD was well-tolerated with minimal side effects and comparable adverse events between treatments. Staff availability restricted recruitment (2.9/month); retention (27/28) and data completeness (95.3%) were excellent. These preliminary findings suggest that AZD may improve wound healing in patients with type 2 diabetes and warrant a fully-powered trial in patients with active ulcers. [Trial Registry: www.isrctn.com/ISRCTN74621291.

**Funding:** MRC Confidence in Concept and NIHR Senior Investigator Award.]

**Single Sentence Summary:** AZD4017 was safe; data suggested improved skin healing / integrity, and modestly reduced epidermal barrier function in patients with type 2 diabetes.

**Disclosure Summary:** I certify that neither I nor my co-authors have a conflict of interest as described above that is relevant to the subject matter or materials included in this Work.

## Introduction

Chronic, non-healing wounds e.g. diabetic foot ulcers are a common worldwide health problem that have substantial medical and socioeconomic importance and represent a major unmet clinical need (*1*). In Europe, 1-1.5% of the population has a problem wound at any one moment in time. The average cost per episode is 6,650€ for leg ulcers and 10,000€ for foot ulcers, accounting for 2-4% of the healthcare budget and likely to escalate with an increasingly elderly and diabetic population (*2*). In the West Yorkshire UK region the overall prevalence of wounds is 2.8-3.6 people per 1000 population (*3*), up to 50% of which are chronically inflamed, non-healing wounds. Costs for wound care in the UK are estimated at £2.03-3.8 million per 100,000 population (*4*) and diabetes currently accounts for approximately 10% of the total health resource expenditure and is projected to increase to 17% by 2035 (*5*).

The profound atrophic effects of glucocorticoids (GC) on human skin structure and function are well documented, causing decreased collagen content, increased transepidermal water loss (TEWL), dermal and epidermal thinning, telangiectasia, impaired wound healing (WH) and increased infection risk (*6-13*). Keratinocytes, melanocytes, fibroblasts and sebocytes play significant roles as GC targets in these processes (*11, 14*). These effects arise from GC excess including systemic (*7, 8*) and topical (*10*) GC therapy, Cushing’s disease (*6*) and psychological stress (*13, 15-17*).

The expertise of our group has focused on 11β-hydroxysteroid dehydrogenase (11β-HSD) isozymes which regulate local GC availability in many tissues largely independently of circulating levels (*18*). In skin, 11β-HSD1 generates cortisol from cortisone, is expressed in epidermal keratinocytes, hair follicles, sebaceous glands and dermal fibroblasts and is upregulated by GC in a forward-feedback manner (*19, 20*). Conversely, 11β-HSD2 converts cortisol to cortisone and, in skin, is predominantly expressed in eccrine (sweat) glands where it functions to protect the mineralocorticoid receptor from inappropriate activation by GC (as with other mineralocorticoid-responsive tissues e.g. kidney) (*21*). We recently demonstrated increased 11β-HSD1 activity during the inflammatory phase of wound healing in a mouse model (*22*) and faster healing in 11β-HSD1-null mice treated with oral corticosterone (active GC in mouse) and topical application of carbenoxolone (CBX), an 11β-HSD non-selective inhibitor (*23*). Strikingly, mice with global deletion of 11β-HSD1 were also protected from age-induced dermal atrophy, with improved collagen processing and accelerated wound healing (*19*). Others have also reported accelerated wound healing by 11β-HSD1 blockade in animal models of diabetes and GC excess (*24, 25*).

These findings suggest that 11β-HSD1 mediates the effects of circulating GC in skin and drives the cutaneous consequences of GC excess. However, the role of 11β-HSD1 in regulating skin function in man remains unexplored, despite evidence that 11β-HSD1 is upregulated by pro-inflammatory cytokines e.g. IL-1β and TNF (*26-28*) abundant in chronic wounds, and reports of increased systemic GC levels in patients with type 2 diabetes mellitus (T2DM) (*29, 30*). Moreover, pro-inflammatory cytokines and GC synergistically increase 11β-HSD1 expression (*31*) which may exacerbate GC availability and further impede wound healing.

Targeting 11β-HSD1 through selective inhibitors has been a focus of major pharma for the last 10 years, the main target indication being metabolic syndrome to improve glucose tolerance, reduce hepatic gluconeogenesis, steatosis and visceral adipogenesis. Proof of concept was established in a number of phase 2 studies but effect size prevented progression to phase 3 studies (*32-35*). However, the ability of systemic 11β-HSD1 inhibitors to target enzyme activity in skin and regulate skin function in humans is unknown.

Our trial objectives were to generate efficacy, safety and study feasibility data to inform a future confirmatory phase 2b trial in patients with diabetic foot ulcers. Our study variables were a combination of validated disease-related outcomes and skin-specific measures previously unexplored in T2DM that greatly improve our knowledge GC metabolism as a regulator of skin function.

## Results

A Consolidated Standards of Reporting Trials (CONSORT) flow diagram is presented in Figure 1.

**Figure 1.**
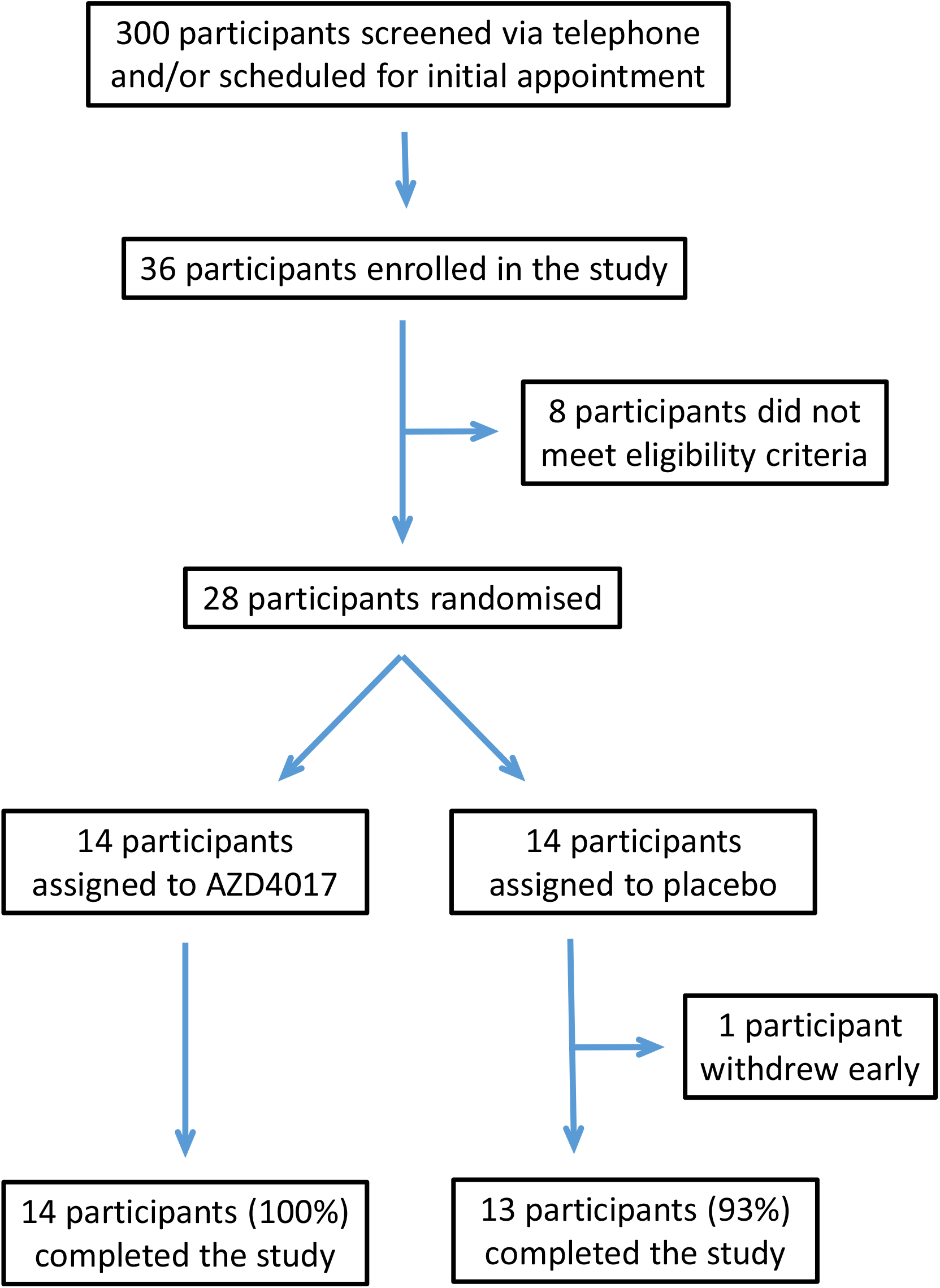
CONSORT flow diagram. Progress, from screening through to study completion, of participants in the double-blind randomised placebo-controlled trial testing a 35 day oral AZD4017 treatment versus placebo in adults with type 2 diabetes. A total of 300 prospective participants were screened. Eight of the 36 individuals enrolled in the study were not randomised owing to not meeting eligibility criteria. 27 out of 28 participants who were randomised completed the study.

### Feasibility

Assessment of recruitment, eligibility, consent and data completeness found conduct of a future confirmatory trial to be feasible. Further details are provided in Appendix 1.

### Participant demographics and baseline variables

Participant demographics, baseline efficacy and laboratory safety variables are presented in Table 1. Full descriptive summaries of efficacy and laboratory safety variables are presented in Tables S1 and S2, respectively. The two randomised arms were well balanced for efficacy and safety variables with the possible exception of sudomotor function in the feet and platelets, which were higher and diastolic BP which was lower in the AZD4017 arm.

**Table 1:** Baseline demographics, primary and secondary efficacy variables, laboratory safety variables. Population: Full analysis set.

| Variable | Summary | PCB | AZD | Total |
| --- | --- | --- | --- | --- |
|  |  | N=14 | N=14 | N=28 |
| Age | Mean (SD), range | 60.3 (13.4), 31.0 to 84.0 | 60.1 (14.5), 28.0 to 75.0 | 60.2 (13.7), 28.0 to 84.0 |
| Male | n/N (%) | 12/14 (86) | 10/14 (71) | 22/28 (79) |
| 11bHSD1 activity radioassay (% conv/24hrs) | Median (Q1, Q3) | 15.3 (11.6, 18.4) | 10.7 (9.4, 17.4) | 13.6 (9.8, 17.9) |
| 11bHSD1 activity ELISA (% conv/24hrs) | Median (Q1, Q3) | 6.8 (5.5, 15.6) | 6.4 (4.2, 8.5) | 6.8 (4.8, 10.7) |
| Sudomotor function Left Hand (micro S) | Median (Q1, Q3) | 54.0 (47.0, 63.0) | 57.5 (39.0, 71.0) | 56.0 (46.5, 68.5) |
| Sudomotor function Right Hand (micro S) | Median (Q1, Q3) | 54.0 (47.0, 60.0) | 56.5 (41.0, 67.0) | 56.0 (45.5, 61.5) |
| Sudomotor function Hands (micro S) | Median (Q1, Q3) | 53.8 (48.0, 60.5) | 56.8 (40.0, 70.5) | 56.8 (47.3, 65.0) |
| Sudomotor function Left Foot (micro S) | Median (Q1, Q3) | 65.5 (46.0, 78.0) | 79.0 (63.0, 84.0) | 74.5 (57.0, 81.5) |
| Sudomotor function Right Foot (micro S) | Median (Q1, Q3) | 69.5 (48.0, 79.0) | 77.0 (69.0, 82.0) | 74.0 (54.5, 79.0) |
| Sudomotor function Feet (micro S) | Median (Q1, Q3) | 67.5 (44.5, 76.5) | 78.0 (64.0, 83.5) | 75.0 (55.8, 79.8) |
| Sudomotor function Overall (micro S) | Median (Q1, Q3) | 61.0 (46.8, 67.8) | 66.6 (63.3, 71.3) | 64.6 (50.3, 69.8) |
| Skin hydration (A.U) | Median (Q1, Q3) | 40.5 (34.5, 46.2) | 40.4 (36.7, 45.5) | 40.5 (36.1, 45.8) |
| Epidermal thickness (micro m) | Median (Q1, Q3) | 62.8 (54.5, 69.2) | 65.7 (60.7, 69.3) | 63.8 (58.4, 69.3) |
| Cortisol (mcg/24h) | Median (Q1, Q3) | 68.5 (40.0, 101.0) | 75.5 (48.0, 84.0) | 74.5 (47.5, 94.0) |
| Urinary [THF+alloTHF]/THE ratio | Median (Q1, Q3) | 1.0 (0.8, 1.2) | 1.0 (0.8, 1.1) | 1.0 (0.8, 1.1) |
| Hour 0 TEWL (Set 1; Day 0) | Log scale Mean (SD), n | 2.1 (0.4), n=14 | 2.2 (0.3), n=13 | 2.2 (0.4), n=27 |
|  | Geometric Mean | 8.2 | 9.1 | 8.6 |
| N tapes required for barrier disruption | Log scale Mean (SD), n | 3.8 (0.4), n=14 | 3.9 (0.5), n=13 | 3.9 (0.4), n=27 |
|  | Geometric Mean | 44.5 | 50.5 | 47.5 |
| Body Mass Index (kg / m<sup>2</sup>) | Mean (SD) | 33.67 (13.47) | 35.05 (5.68) | 34.36 (10.17) |
| Waist-hip ratio | Mean (SD) | 0.98 (0.08) | 1.03 (0.08) | 1.01 (0.08) |
| Systolic blood pressure (mm Hg) | Mean (SD) | 135.71 (21.01) | 140.43 (12.02) | 138.07 (16.97) |
| Diastolic blood pressure (mm Hg) | Mean (SD) | 83.86 (8.91) | 72.64 (9.96) | 78.25 (10.89) |
| HbA1c (mmol/mol) | Mean (SD) | 72.29 (19.43) | 66.00 (14.91) | 69.14 (17.29) |
| High density lipoprotein (mmol/l) | Mean (SD) | 1.24 (0.31) | 1.19 (0.27) | 1.21 (0.29) |
| Cholesterol (mmol/l) | Mean (SD) | 4.36 (1.13) | 3.95 (0.75) | 4.15 (0.97) |
| Triglycerides (mmol/l) | Mean (SD) | 1.68 (0.74) | 2.02 (1.02) | 1.85 (0.89) |
| Haemoglobin (g/l) | Mean (SD) | 139.21 (14.12) | 139.43 (11.35) | 139.32 (12.57) |
| White cells (x10<sup>9</sup>/l) | Mean (SD) | 6.46 (2.55) | 7.34 (2.04) | 6.90 (2.31) |
| Platelets (x10<sup>9</sup>/l) | Mean (SD) | 218.71 (61.85) | 273.14 (81.37) | 245.93 (76.15) |
| Red cells (x10<sup>12</sup>/l) | Mean (SD) | 4.84 (0.49) | 4.71 (0.48) | 4.78 (0.48) |
|  |  | N=14 | N=14 | N=28 |
| Mean corpuscular volume (fl) | Mean (SD) | 88.43 (4.64) | 91.21 (6.96) | 89.82 (5.98) |
| Haematocrit (packed cell volume) | Mean (SD) | 0.43 (0.04) | 0.43 (0.03) | 0.43 (0.03) |
| Mean corpuscular haemoglobin (pg) | Mean (SD) | 28.82 (2.09) | 29.74 (2.47) | 29.28 (2.29) |
| Corpuscular hemoglobin concentration (g/l) | Mean (SD) | 325.57 (10.43) | 326.64 (8.29) | 326.11 (9.26) |
| Red blood cell distribution width (%) | Mean (SD) | 13.88 (0.98) | 14.12 (1.10) | 14.00 (1.03) |
| Albumin (g/l) | Mean (SD) | 39.43 (2.56) | 39.50 (2.44) | 39.46 (2.46) |
| Blirubin (umol/l) | Mean (SD) | 8.93 (2.97) | 8.14 (2.82) | 8.54 (2.87) |
| Alkaline phosphatase (U/l) | Mean (SD) | 80.43 (23.76) | 85.07 (26.08) | 82.75 (24.59) |
| Alanine aminotransferase (iu/l) | Mean (SD) | 24.64 (7.10) | 27.43 (10.91) | 26.04 (9.14) |
| Aspartate aminotransferase (iu/l) | Mean (SD) | 21.00 (4.95) | 22.64 (5.50) | 21.82 (5.20) |
| Gamma-glutamyl transpeptidase (iu/l) | Mean (SD) | 41.86 (33.87) | 39.29 (29.87) | 40.57 (31.37) |
| eGFR (ml/min/1.73m2) | Mean (SD) | 81.86 (9.69) | 79.21 (13.59) | 80.54 (11.66) |
| Sodium (mmol/l) | Mean (SD) | 138.93 (2.06) | 140.71 (5.47) | 139.82 (4.15) |
| Potassium (mmol/l) | Mean (SD) | 4.46 (0.33) | 4.63 (0.41) | 4.55 (0.37) |
| Urea (mmol/l) | Mean (SD) | 6.96 (2.81) | 7.25 (2.17) | 7.10 (2.47) |
| Creatinine (umol/l) | Mean (SD) | 76.64 (15.36) | 76.21 (20.04) | 76.43 (17.52) |
| Testosterone (nmol/l) | Mean (SD) | 11.10 (6.25) | 6.97 (5.48) | 9.04 (6.14) |
| Dehydroepiandrosterone sulphate (umol/l) | Mean (SD) | 3.53 (2.19) | 2.91 (1.69) | 3.22 (1.95) |
| Free thyroxine (pmol/l) | Mean (SD) | 14.86 (2.20) | 15.73 (1.58) | 15.30 (1.93) |
| Thyroid stimulating hormone (mIU/l) | Mean (SD) | 1.74 (0.66) | 1.72 (0.63) | 1.73 (0.64) |

Nn=Number non-missing; Q1=1st quartile; Q3=3rd quartile

### Compliance

Mean diary card compliance was >97.9% across both arms at all visits and >99.6% at day 35. Mean IMP compliance at each visit was >95.2% across both arms and reached >97.9% at day 35; overall IMP compliance was 84-101% (PCB) and 93-101% (AZD4017) (Table S3). Drug exposure data in plasma and skin did not indicate any discrepancies between reported compliance and actual exposure, suggesting participants had correctly adhered to the treatment regimen. Further, in the group receiving AZD4017 (n=12 with data available), there was a moderate correlation of Spearman’s rho=0.54 between levels in the skin, taken from the biopsy at day 28, and in plasma samples taken at day 35 (Figure S1).

### Efficacy

#### 11β-HSD1 activity in skin

Our primary outcome evaluated the ability of AZD4017 to inhibit 11β-HSD1 activity in skin. Median 11β-HSD1 activity (% conversion per 24hr) by radioassay at day 28 was similar across treatment groups [adjusted for baseline activity, baseline glycated haemoglobin (HbA1c), age and sex: median PCB=11.8, AZD4017=12.8; difference AZD-PCB (90% CI) 1.1 (−3.4, 5.5)] (Figure 2a, Table 2), similar to unadjusted estimates (Table S4) and comparable to baseline (Table 1). Proof-of-concept was not met because median activity was higher in the AZD4017 group than placebo at day 28; all CIs included 0. This conclusion was unaffected after removal of 2 baseline samples which failed QC prior to re-imputation (Table S5). Changes from baseline (Tables S6-S7) indicated that activity went down in both PCB and AZD4017 groups, but to a greater extent in PCB.

**Table 2:**
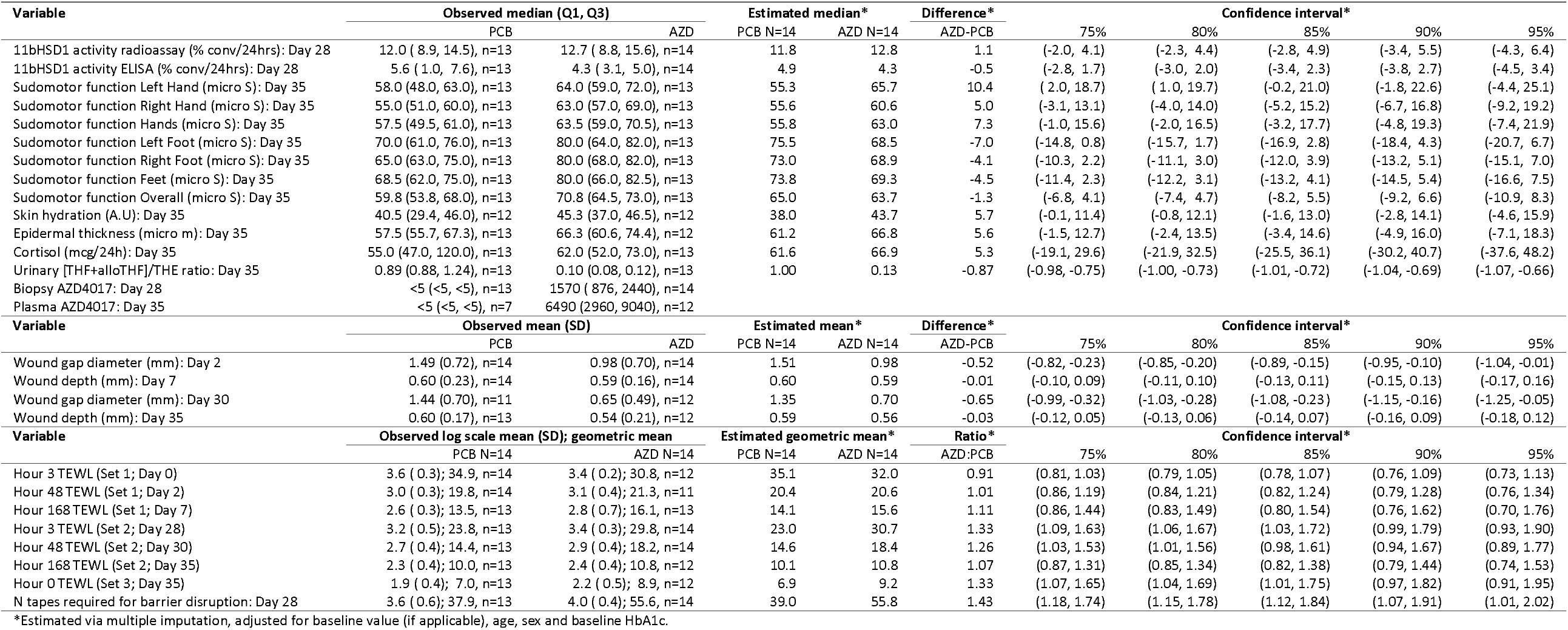
Primary and secondary efficacy outcomes; adjusted differences between treatment groups. Population: Full analysis set. Multiple imputation was used to address missing data. Due to issues with the data distributions, for all variables except TEWL, integrity, wound diameter & depth, median regression was used to estimate confidence intervals around differences between the groups. For TEWL, integrity, wound depth and diameter, linear regression was used. TEWL and integrity measurements were log-transformed prior to analysis; differences have been expressed as ratios of geometric means (AZD:PCB). For each variable, the comparison was adjusted for baseline value of the variable (not applicable to wound diameter & depth), age, sex and baseline HbA1c. All TEWL readings were adjusted using TEWL baseline at day 0. Between group differences were not calculated for biopsy and plasma AZD4017 because available PCB values did not vary, all were below the detection limit.

**Figure 2.**
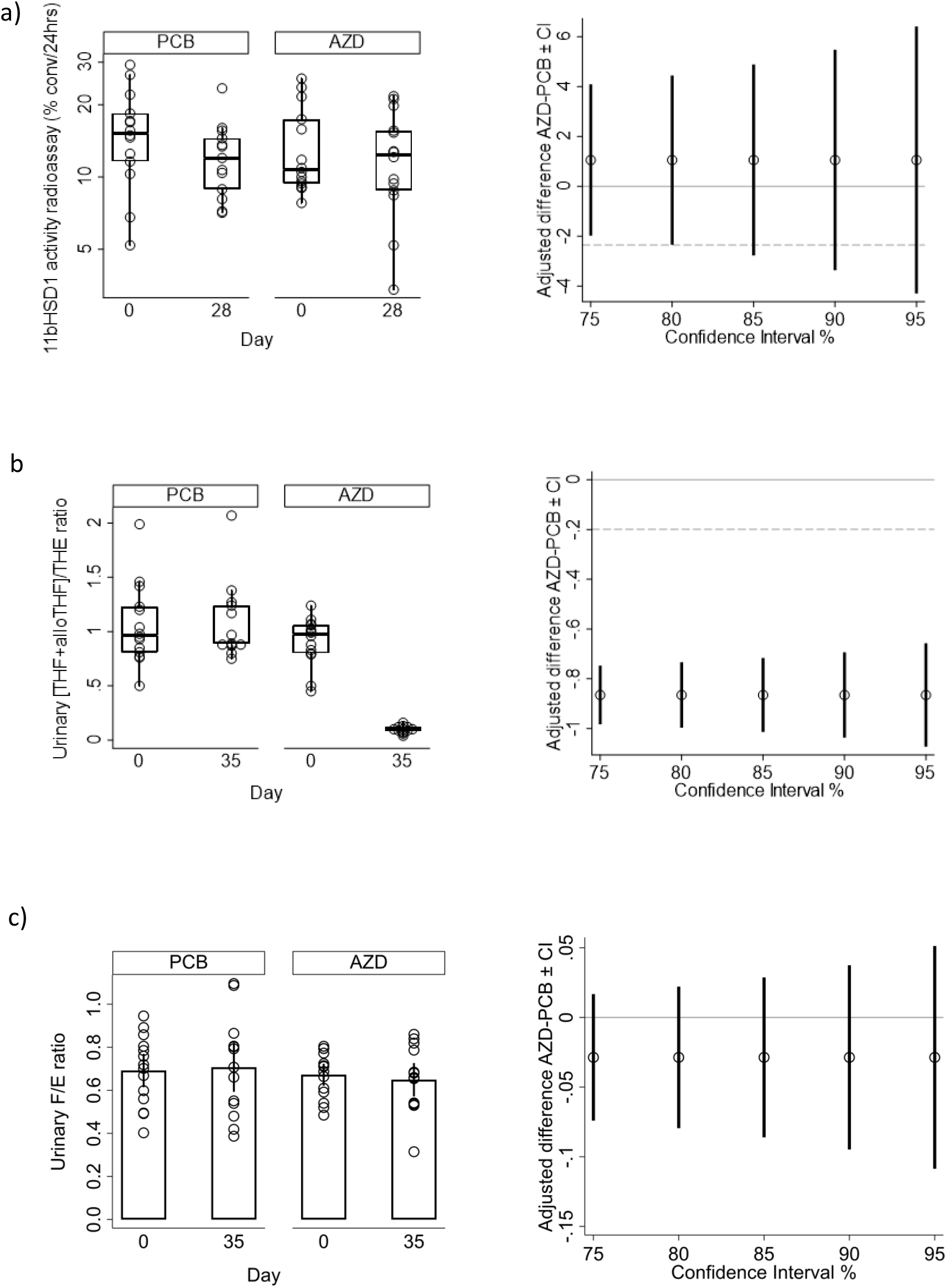
Efficacy outcome measures. Adjusted differences between placebo (PCB) and AZD4017 (AZD) groups in absolute values of primary and secondary efficacy outcomes in the full analysis set. Box plots of observed values (left panel) and adjusted differences between medians estimated in imputed data (right panel) are presented. Solid lines indicate no difference between groups (=0). Dotted lines indicate 20% improvement relative to the median PCB (not applicable to F/E ratio).

A sensitivity analysis was conducted using an alternative ELISA method, which demonstrated an acceptable correlation with the radioassay method at baseline (rho=0.70) but not day 28 (rho=0.19) (Figure S2, Table 3). In the primary analysis, median activity measured via ELISA was lower in the AZD4017 group, but all CIs included 0. In a sensitivity analysis which excluded an extremely high outlier in the placebo group (Table S5), the median was higher in the AZD4017 group, consistent with the radioassay method finding. Therefore, and mindful of the relatively low levels of ex vivo 11β-HSD1 activity over 24 hours, there was no proof-of-concept that AZD4017 was able to inhibit 11β-HSD1 activity in skin.

**Table 3:**
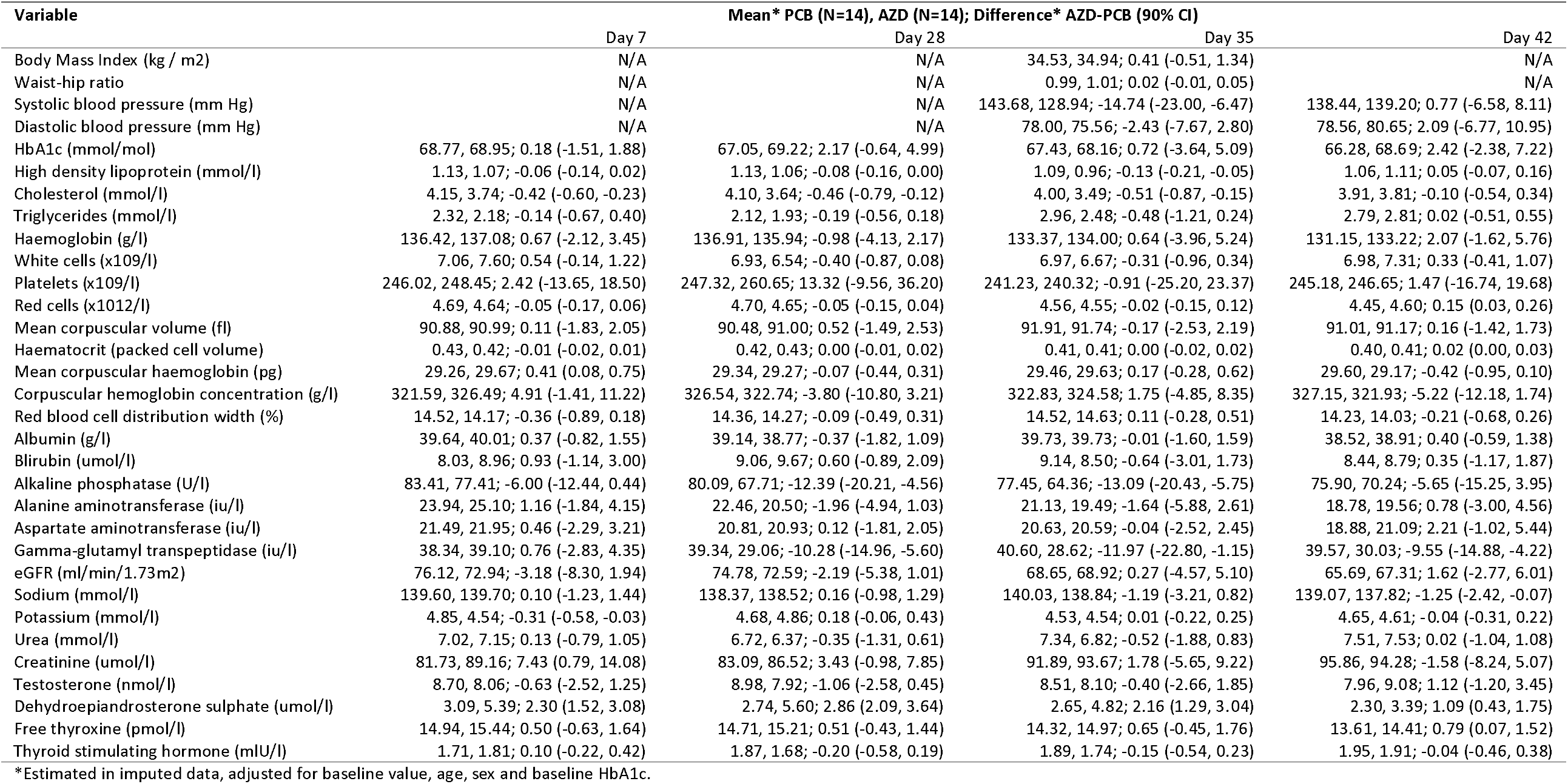
Longitudinal laboratory safety variables; adjusted differences between treatment groups. Population: Safety set. All point estimates and confidence intervals estimated via linear regression.

#### Systemic 11β-HSD1 activity

In contrast to the findings in the skin, systemic 11β-HSD1 activity, measured by urinary [THF+alloTHF]/THE ratio, differed to a substantive degree between groups at day 35. Values were lower in the AZD4017 group; adjusted median PCB=1.00, AZD4017=0.13; difference (90% CI) −0.87 (1.04, −0.69) (Figure 2b, Table 2). All supplementary CI up to 95% excluded 0. This suggests median systemic 11β-HSD1 activity may be ≥69% (median 87%) lower in subjects taking AZD4017. As anticipated, there was no evidence that urinary F/E ratio (a measure of systemic 11β-HSD2 which deactivates cortisol to cortisone; unplanned analysis) was affected by AZD4017 (Figure 2c, Table 2).

#### Wound healing

Separate biopsies were taken at baseline and day 28 from the outer forearm. In each case, based on maximal granulation tissue width (a marker of early wound healing), there was preliminary evidence that the mean wound gap 2 days following the biopsy differed between groups, at all levels of confidence. The 90% CI suggested that the mean gap was potentially 7% to 63% (mean 34%) narrower at day 2 and 12% to 85% (mean 48%) narrower at day 30 in the AZD4017 group, relative to placebo (Figure 3a-b, Table 2). Representative wound healing images for participants treated with placebo and ADZ4017 (Figure 3c) are presented.

**Figure 3.**
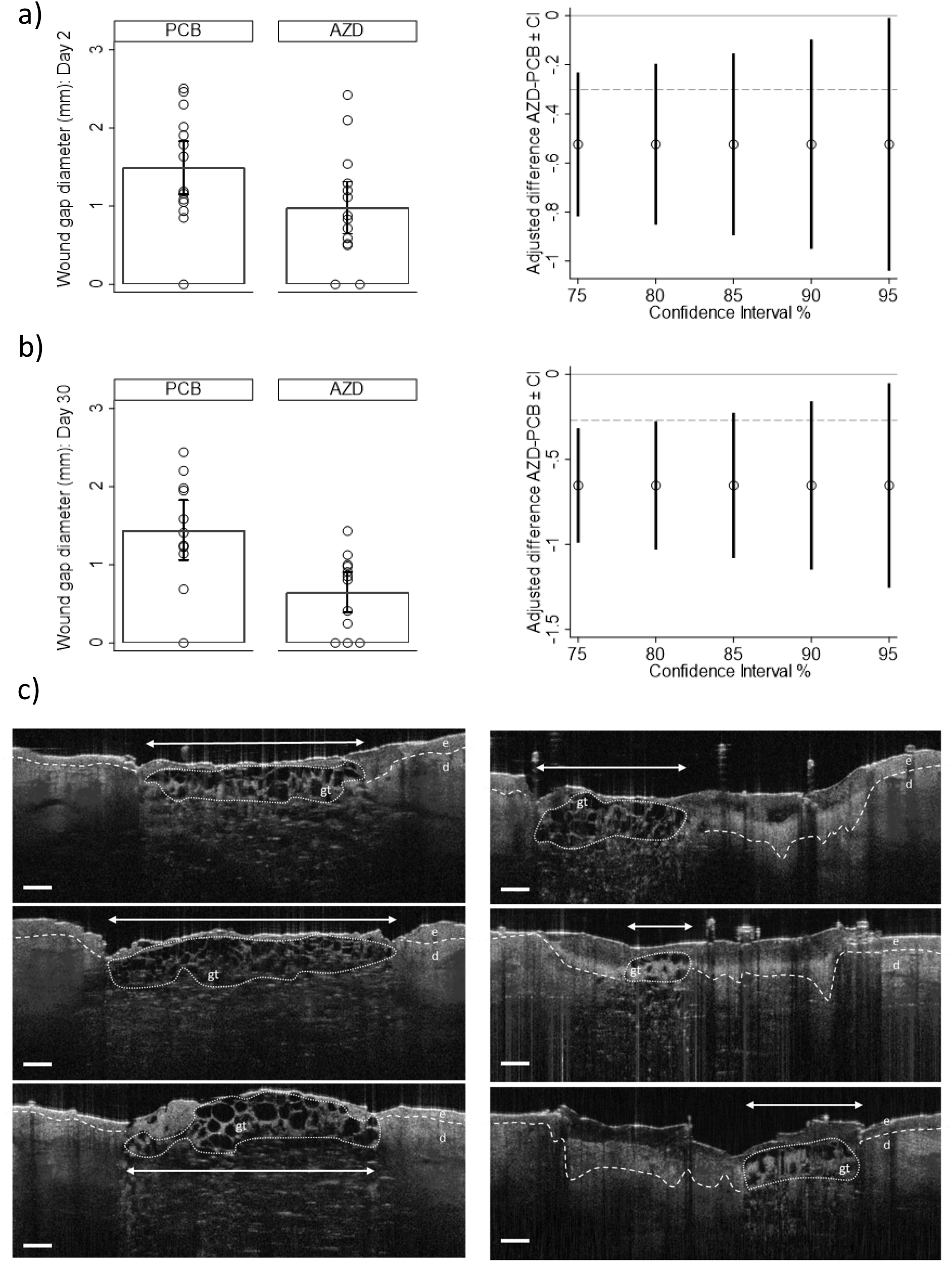
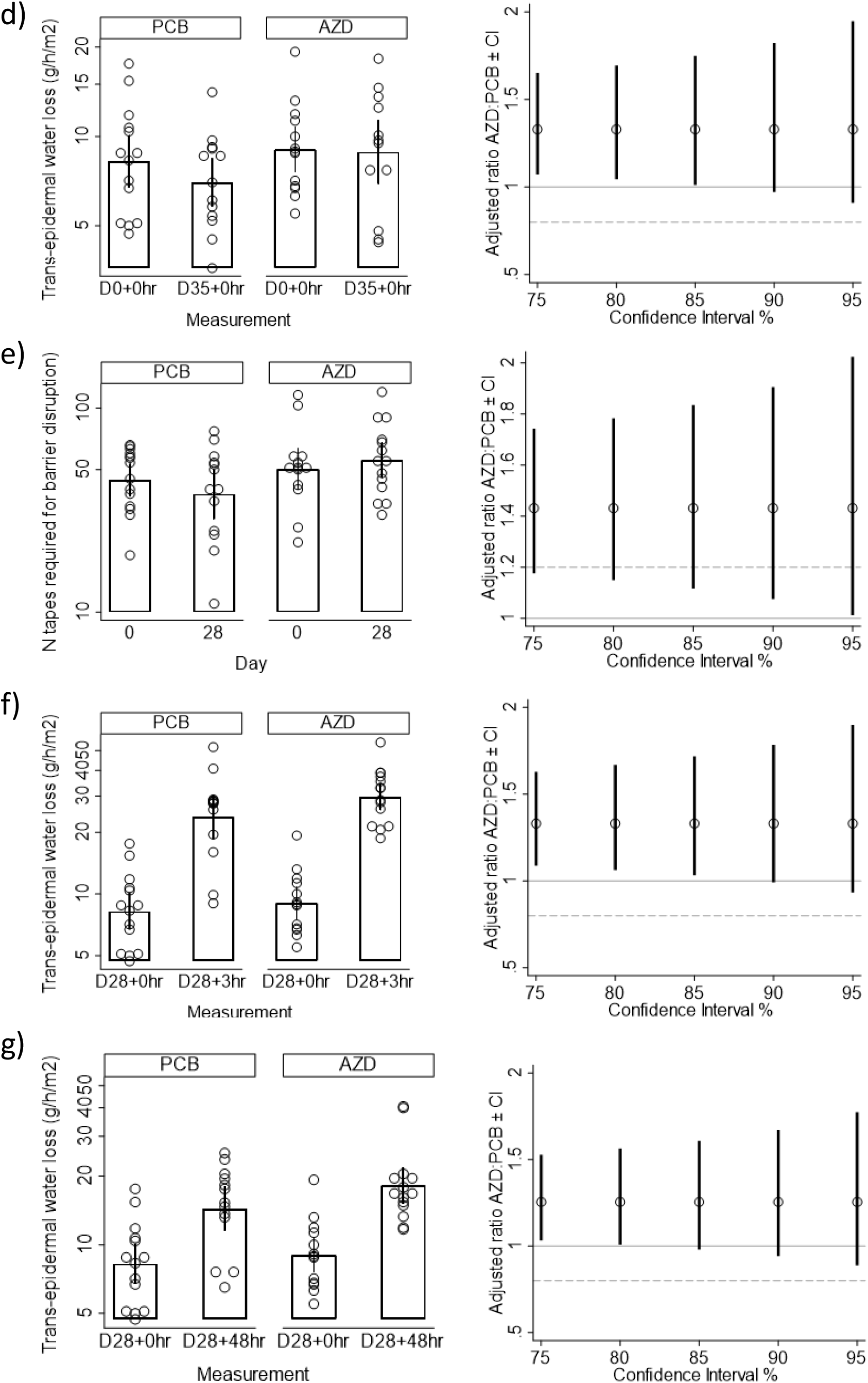
Skin outcome measures. (a-b, d-g) Adjusted differences between placebo (PCB) and AZD4017 (AZD) groups in absolute values of secondary efficacy outcomes in the full analysis set. Error bar plots of observed values (indicating 90% CIs; left panel) and adjusted differences between means estimated in imputed data (right panel) are presented. TEWL and integrity readings were log-transformed prior to analysis therefore geometric means and ratios have been provided for these variables. Solid lines indicate no difference between groups (=0 for WH, =1 for TEWL and integrity). Dotted lines indicate 20% improvement relative to the mean in PCB. (c)Representative wound healing OCT images at day 2 (2 days post-biopsy and 2 days of AZD4017 treatment). Maximal early granulation tissue width (arrow) was measured for participants treated with placebo (left panel) or AZD4017 (right panel). Wide dashes indicate dermal-epidermal junction. Epidermis = e. Dermis = d. Granulation tissue = gt Scale bar = 250µm.

Based on maximal clot depth (a marker of late wound healing), when re-measured 7 days post biopsy, there was no preliminary evidence of a difference between groups at either time-point (on days 7 and 35, Table 2).

#### Epidermal barrier function

Resting transepidermal water loss (TEWL), the gold-standard measure of epidermal barrier function, was on average 33% higher in the AZD4017 group on day 35 (Figure 3d, Table 2), suggesting a potentially deleterious effect on epidermal barrier function. Although CIs were wide, some excluded 0; at 85% confidence the results suggested between +1% and +75% higher TEWL in the AZD4017 group, but the 90% CI was −3% to +82%.

Sensitivity analysis re-estimated the 90% CI to be +3% to +88% (Table S5).

#### Epidermal integrity

The number of tapes required to achieve a barrier disruption TEWL of 40-50 g/h/m^2^ on day 28 was observed to be higher in the AZD4017 group (at all levels of confidence). The 90% CI indicated preliminary evidence of a difference of between +7% and +91% (mean +43%) relative to placebo (Figure 3e, Table 2). This was equivalent to an additional 16.8 tapes required to induce a comparable degree of barrier disruption in the AZD4017 group.

#### Epidermal barrier recovery

In the first set of post-disruption recovery measurements, collected between baseline and day 7, there was no proof-of-concept for a difference in TEWL; all CIs straddled 1 (Table 2).

In the second set, collected between day 28 and day 35, there was some preliminary evidence of potential differences in 3hr and 48hr recovery, although CIs were wide. For 3hr post-disruption, the 90% CI included 0 (−1% to +79%) but with 85% confidence, this may be between 3% and 72% higher (mean +33%) in the AZD4017 group following 28 days of treatment (Figure 3f, Table 2). Similarly, the primary 90% CI included 0 for 48hr post-disruption TEWL (−6% to 67%), but at 80% confidence there was preliminary evidence of higher TEWL in the AZD4017 group by +1% to 56% (mean +26%) relative to placebo (Figure 3g, Table 2). By 168hr post-disruption, although median TEWL was still higher in the AZD4017 group, all CI included 0 (no difference) (Table 2).

Sensitivity analyses did not differ from the main conclusions (table S5, S8).

#### Epidermal thickness

Although median epidermal thickness was greater in the AZD4017 group, consistent with proof-of-concept, all CIs included 0 (no difference) (Table 2). The 90% CI indicated that median skin thickness could be between 8% thinner to 26% thicker in the AZD4017 group relative to placebo.

#### Skin hydration

Although the medians suggested better hydration in the AZD4017 group, consistent with proof-of-concept, the 90% CI indicated median hydration could be anywhere between 7% lower and 37% higher in those receiving AZD4017; all CIs included 0 (Table 2).

#### Sudomotor function

Across all sites, sudomotor function was equivocal as an effect in a particular direction; all CI included 0 (Table 2). This was also true when the results were averaged across hands and feet separately. The 75% and 80% CIs excluded 0 in the left hand (but not the right) with 80% CI indicating between 2% and 36% greater function.

Sensitivity analyses indicated at 75% CI the possibility of between 1% and 13% lower sudomotor function in feet and in those taking AZD4017 relative to placebo.

### Correlations between compliance and efficacy outcomes

Several outcome measures displayed preliminary evidence of an association with compliance (absolute rho>=0.3) including negative associations with 11β-HSD1 activity by ELISA and [THF+alloTHF]/THE ratios and positive associations with TEWL and skin hydration (table S11). However, given the narrow range of compliance variability (and overall high compliance), these putative associations should be interpreted with caution.

### Additional sensitivity analyses of primary and secondary variables

Sensitivity analyses using available case (Table S9) and last observation carried forward (Table S10) as alternative methods of handling missing data did not yield results that differed substantially from the results of the main analyses, unless already highlighted above.

### Safety

#### Biopsy and ECG

Biopsy sites did not raise any clinical concerns and all were found to pass a physical inspection. No incidents of infection were observed. Participants did not report any pain or discomfort from the biopsies, which all healed relatively well.

No clinically meaningful ECG anomalies were found in any patients remaining in the trial at day 42 (placebo n=13; AZD4017 n=14).

#### Longitudinal laboratory safety data

The 90% CI for the (baseline value, age, sex and baseline HbA1c) adjusted difference between treatment groups indicated there was preliminary evidence that in the AZD4017 group relative to placebo (Table 3): dehydroepiandrosterone sulphate (DHEAS) was higher at days 7, 28, 35 & 42 (Figure 4a), cholesterol was lower at days 7, 28 and 35 (Figure 4b), high density lipoprotein (HDL) was lower at day 35 (Figure 4c) and systolic blood pressure (BP) was lower at day 28 (Figure 4d).

**Figure 4.**
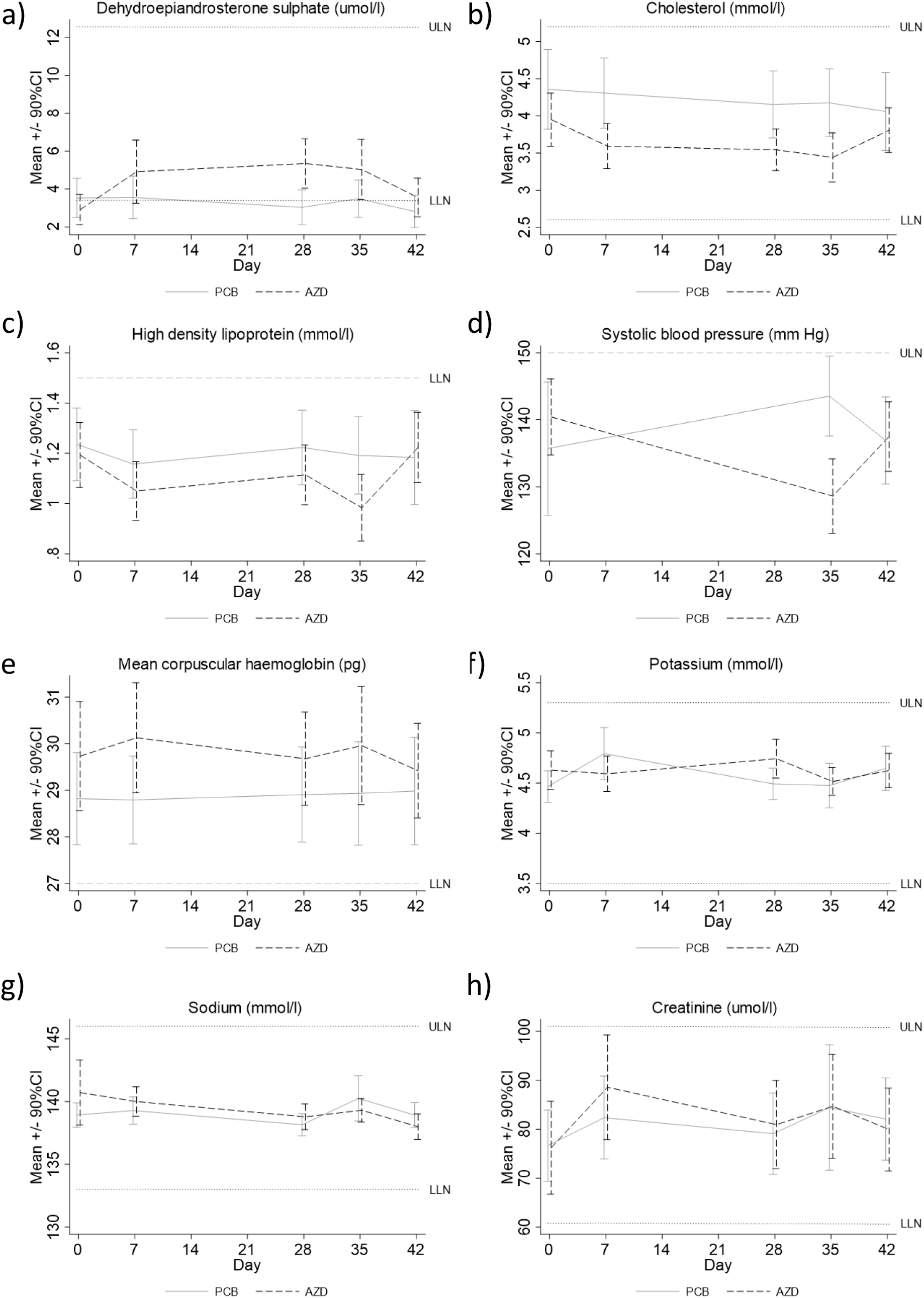
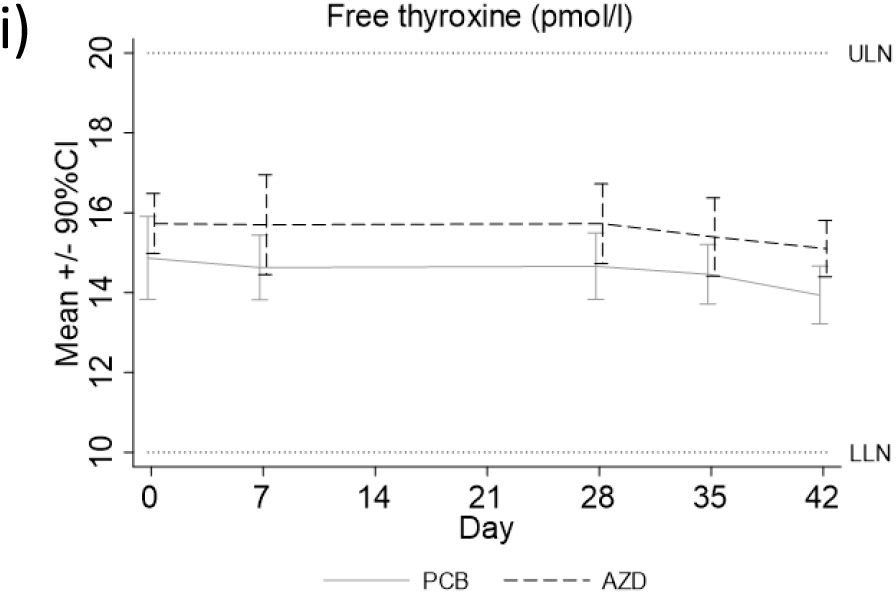
Safety. Displays of longitudinal laboratory safety data. Population: Safety set. ULN = upper limit of normal; LLN = lower limit of normal (where applicable).

Mean corpuscular Hb (Figure 4e) and potassium (Figure 4f) were lower at day 7, sodium was lower at day 42 (Figure 4g), creatinine was higher at day 7 (Figure 4h) and free thyroxine was higher at day 42 (Figure 4i). Most of these apparent differences were small in magnitude. For all except sodium, potassium and free thyroxine, the adjusted difference at day 42, 7 days after the end of treatment, was smaller than at day 35.

The remaining continuous safety variables did not show any preliminary differences between AZD4017 and placebo in this study, although this may have been due to the small sample size and relatively short treatment duration (Table 3). Unadjusted estimates (Table S12) were similar, although platelets were higher in AZD at each visit, values were already higher at baseline in this group. Unadjusted and adjusted changes from baseline are presented for all laboratory safety variables in Tables S13 and S14 respectively.

Although there were several values in each treatment group that were above or below accepted clinical normal limits (Table S15), there were relatively few laboratory findings that were sufficient cause for concern to warrant further investigation or intervention by the study team, and none resulted in withdrawal from IMP.

#### Adverse events

There were a total of 37 adverse events (AE) (placebo n=13; AZD4017 n=24); counting recurring instances of the same AE within a patient just once, there were 29 unique AE (placebo n=13; AZD4017 n=16) (Table 4). The majority of AE were gastrointestinal, nervous system or respiratory, and the majority were of mild severity (n=26), with only 3 of moderate severity and none of greater severity. None of the 29 unique AE were considered probably or definitely study drug related, most were deemed possibly related (n=20) or unlikely to be relate (n=8). AE were broadly balanced across the treatment arms, although all three of the moderately severe events occurred in the AZD4017 group.

**Table 4:** Adverse event summary. Population: Safety set

| Adverse events (AE) | PCB (n=14) | AZD (n=14) | All patients (n=28) |
| --- | --- | --- | --- |
| Total AE: n (n per PY) | 13 (8.3) | 24 (13.8) | 37 (11.1) |
| Unique AE: n (n per PY) | 13 (8.3) | 16 (9.2) | 29 (8.7) |
| Patients with >=1 AE: n (%) | 8 (57) | 10 (71) | 18 (64) |
| Discontinuation due to AE: n (%) | 0 (-) | 0 (-) | 0 (-) |
| **AE by SOC: n total (n unique\*), n patients (%)** |  |  |  |
| Blood | 0 (0), 0 (-) | 0 (0), 0 (-) | 0 (0), 0 (-) |
| Cardiac | 0 (0), 0 (-) | 0 (0), 0 (-) | 0 (0), 0 (-) |
| Congenital | 0 (0), 0 (-) | 0 (0), 0 (-) | 0 (0), 0 (-) |
| Ear | 0 (0), 0 (-) | 0 (0), 0 (-) | 0 (0), 0 (-) |
| Endocrine | 0 (0), 0 (-) | 0 (0), 0 (-) | 0 (0), 0 (-) |
| Eye | 0 (0), 0 (-) | 0 (0), 0 (-) | 0 (0), 0 (-) |
| Gastrointestinal | 9 (9), 6 (43) | 14 (8), 7 (50) | 23 (17), 13 (46) |
| General | 0 (0), 0 (-) | 0 (0), 0 (-) | 0 (0), 0 (-) |
| Hepatobiliary | 0 (0), 0 (-) | 0 (0), 0 (-) | 0 (0), 0 (-) |
| Immune | 0 (0), 0 (-) | 0 (0), 0 (-) | 0 (0), 0 (-) |
| Infections | 0 (0), 0 (-) | 1 (1), 1 (7) | 1 (1), 1 (4) |
| Injury | 0 (0), 0 (-) | 0 (0), 0 (-) | 0 (0), 0 (-) |
| Investigations | 0 (0), 0 (-) | 0 (0), 0 (-) | 0 (0), 0 (-) |
| Metabolism | 0 (0), 0 (-) | 0 (0), 0 (-) | 0 (0), 0 (-) |
| Musculoskeletal | 0 (0), 0 (-) | 1 (1), 1 (7) | 1 (1), 1 (4) |
| Neoplasms | 0 (0), 0 (-) | 0 (0), 0 (-) | 0 (0), 0 (-) |
| Nervous system | 1 (1), 1 (7) | 6 (4), 4 (29) | 7 (5), 5 (18) |
| Pregnancy | 0 (0), 0 (-) | 0 (0), 0 (-) | 0 (0), 0 (-) |
| Psychiatric | 0 (0), 0 (-) | 0 (0), 0 (-) | 0 (0), 0 (-) |
| Renal | 0 (0), 0 (-) | 0 (0), 0 (-) | 0 (0), 0 (-) |
| Reproductive system | 0 (0), 0 (-) | 0 (0), 0 (-) | 0 (0), 0 (-) |
| Respiratory | 2 (2), 1 (7) | 2 (2), 1 (7) | 4 (4), 2 (7) |
| Skin | 1 (1), 1 (7) | 0 (0), 0 (-) | 1 (1), 1 (4) |
| Social | 0 (0), 0 (-) | 0 (0), 0 (-) | 0 (0), 0 (-) |
| Surgical | 0 (0), 0 (-) | 0 (0), 0 (-) | 0 (0), 0 (-) |
| Vascular | 0 (0), 0 (-) | 0 (0), 0 (-) | 0 (0), 0 (-) |
| **AE severity: n\*** |  |  |  |
| Mild | 13 | 13 | 26 |
| Moderate | 0 | 3 | 3 |
| Severe | 0 | 0 | 0 |
| Life-threatening | 0 | 0 | 0 |
| Death | 0 | 0 | 0 |
| **AE by relation to study drug: n\*** |  |  |  |
| Not related | 0 | 1 | 1 |
| Unlikely | 4 | 4 | 8 |
| Possible | 9 | 11 | 20 |
| Probable | 0 | 0 | 0 |
| Definite | 0 | 0 | 0 |
| **SAE** |  |  |  |
| Total SAE: n | 0 | 1 | 1 |
| Patients with >=1 SAE: n (%) | 0 (-) | 1 (7) | 1 (4) |

\* Recurrent AEs counted once at maximum severity/relatedness reported. SOC=CTCAE system order class; PY=patient year

### Future study power estimation

Based on the power calculations from the current trial (table S16), we anticipate that n=100-150 per arm should suffice to detect a difference of 20% or more relative to placebo for all outcome measures.

## Discussion

Our double-blind, randomised, placebo-controlled, parallel clinical trial provides preliminary evidence that wounds were 12-85% (mean 48%) smaller and skin integrity was 7-91% (mean 43%) stronger in patients with type 2 diabetes mellitus following 11β-HSD1 inhibition. This represents a major advance in the development of 11β-HSD1 inhibitors as a novel therapy for diabetic ulcers.

Studies had previously demonstrated effective 11β-HSD1 inhibition in mouse skin (*19, 23*), but this had not been explored in humans. Our primary outcome measure failed to evidence 11β-HSD1 inhibition in skin. However, preliminary proof-of-concept was demonstrated though a reduction in [THF+alloTHF]/THE ratios (the gold-standard measure of systemic 11β-HSD1 activity) and elevated DHEAS consistent with other selective 11β-HSD1 inhibitor trials, including AZD4017 (*34, 36-39*). As anticipated, 11β-HSD2 activity was unaffected (*40, 41*). The lack of peripheral inhibition was not due to a lack of exposure *in situ* as skin biopsy AZD4017 correlated with plasma levels, albeit at lower concentrations. However, a loss of efficacy ex vivo (lower target affinity when removed from much higher *in situ* circulating AZD4017 levels) or insufficient assay sensitivity (mean baseline 11β-HSD1 activity was relatively low) may explain this finding.

Our secondary outcomes offer a series of novel insights into the effects of 11β-HSD1 inhibition on skin function. OCT is a validated method for non-invasive assessment of wound healing (*42*) and our study is the first application of this method in a randomised controlled trial. Our preliminary finding of improved wound healing at day 2 post-biopsy after less than two days’ AZD4017 treatment is very promising. More so, given that overall healing in this cohort was relatively normal and the diabetes relatively well-managed. Evidence indicative of improved healing by AZD4017 was obtained after just two days of treatment and the preliminary estimates of effect size were greater after 30 days treatment. The fact that all CI included considerable improvements relative to placebo, and this was finding was repeatable in the two separate biopsies, provides good justification for progressing to a larger trial. This novel human data is consistent with the known deleterious effects of GC on wound healing (*43*) and with pre-clinical evidence of improved healing by 11β-HSD1 inhibition in animal models of stress, obesity, GC excess and ageing from our group and others (*19, 23-25*). A recent study in *db/db* mice also reported increased 11β-HSD1 activity during wound healing compared to wild-type mice, although this remains to be explored in humans (*44*). By day 7 post-wounding, there was no preliminary evidence of an effect. However, this may have been limited by measurement of maximal clot depth, rather than volume, which should be addressed in future studies. Further, no granulation tissue was detectable at this time point, consistent with normal progression of healing.

To strengthen the case for potential skin benefits of AZD4017, we also found preliminary evidence that AZD4017 may increase epidermal integrity, an improvement comparable to that of young vs. aged skin (*45*). This is supported by studies demonstrating reduced skin integrity following human psychological stress (*46*) or exogenous GC treatment (*47*). In contrast, we previously found increased epidermal integrity in mice treated with oral corticosterone which was reversed by 11β-HSD1 inhibition (*23*), suggesting epidermal integrity regulation by endogenous GC may be opposed in humans and mouse, highlighting the importance of our findings.

Provisional evidence of a modest impairment in epidermal barrier function and recovery by AZD4017 was also observed in our study. GC are known to both impair and promote epidermal barrier function, the former predominantly with exogenous GC treatment (*47-49*) and the latter following endogenous GC excess (*50*). Our results indicate for the first time that endogenous cortisol also promotes epidermal barrier function via 11β-HSD1 in humans.

GC have been shown to impair healing through repression of epidermal growth factor signalling that inhibits keratinocyte re-epithelialisation (essential for wound repair) whilst concomitantly promoting keratinocyte differentiation to promote barrier function (*11*). Our differential findings of improved wound healing and impaired barrier function by AZD4017 are in agreement, although barrier impairment was more modest. However, future trials exploring possible benefits of 11β-HSD1 inhibition on wound healing should monitor the epidermal barrier, particularly in patients with a compromised function e.g. atopic dermatitis.

Peripheral neuropathy is a serious complication in diabetes. To safeguard against any possible detrimental effects on nerve function, sudomotor analysis was conducted (*51*). Nerve function was unaffected by AZD4017 overall and in the hands. At the lowest level of confidence, a modest impairment was noted in the feet and therefore, monitoring of nerve function should be included in future trials to rule out any adverse effects.

In this relatively short, preliminary study, there were no significant safety concerns. Overall, eGFR and other laboratory safety outcomes gave no evidence of a detrimental effect. Indeed conversely, we found evidence of improvements in some “metabolic” traits, notably high density lipoprotein, cholesterol and systolic blood pressure in the AZD4017 arm which were not present at baseline or after 7 days treatment washout (and despite ongoing treatment for these conditions). These improvements are consistent with results from other selective 11β-HSD1 inhibitor trials (*33-36*).

Importantly, cardiovascular disease markers are a strong predictor of ulcer occurrence, healing, complications and recurrence (*52, 53*). Through alleviating cardiovascular disease burden, oral 11β-HSD1 inhibitors could also reduce ulcer recurrence in high risk patients, but this will require further clinical investigation with long-term 11β-HSD1 inhibitor therapy. Concomitant medication was not formally captured in this trial and based on our findings, warrants monitoring in future studies.

The use of wounds induced by biopsy, rather than active diabetic ulcers, was a key limitation of our study design. However, this was necessary to ensure AZD4017 did not raise any safety concerns in acute wounds and patients with relatively well-managed diabetes before progression to patients with more severe disease. The small sample size and lack of inferential testing, whilst recommended for pilot trials, require all putative findings presented here to be confirmed in fully-powered trials. It is likely that these would have greater resources to support recruitment, which, although target sample size was achieved, was also a limitation.

Our findings make a strong case for a future study in patients with active ulcers where the primary outcome should be an established measure of ulcer healing. Given that approximately 5% of patients with type 2 diabetes have a diabetic foot ulcer, a future confirmatory trial will likely be multi-centre. Stratification of randomisation by key variables known to affect 11β-HSD1 function (e.g. age) and wound healing (e.g. infection / ulcer size at presentation) is also advisable. Between-group differences were strongest following 28 days of AZD4017 treatment. Therefore, a future study schedule could be simplified and coordinated with routine appointments to improve recruitment.

Although requiring confirmation, improvements in healing, skin integrity and metabolic risk factors of ulcer recurrence hold promise for 11β-HSD1 as a novel therapeutic target in wound repair. A fully-powered clinical trial in patients with active ulcers and type 2 diabetes mellitus is now warranted.

## Materials and Methods

The final approved trial Protocol v2.0 24th October 2018 is provided in Appendix 2.

### Study Design

This was a randomised, double-blind and parallel group, placebo-controlled, phase II pilot trial in patients with T2DM. Participants were recruited between 28^th^ March 2018 and 23^rd^ January 2019 (first to last randomised participant telephone invitations) with last participant last follow-up (telephone discharge) on 3^rd^ April 2019. A total of 28 participants (22 male, 6 female) were randomised and 27 completed the 35 day oral AZD4017 or placebo treatment. For study design details, please refer to section 5 of Appendix 2.

### Selection of sample size

Published guidance for pilot studies recommended a sample size of 12 participants per arm completing the protocol (*54*). To ensure this minimum number completed the protocol, the study aimed to randomise 15 per arm to allow for 20% drop-out. Please refer to section 13.9 in Appendix 2 for details.

### Consent, Ethical Approval and Oversight

Informed consent was obtained after the nature and possible consequences of the study had been explained. Full ethical approval was acquired prior to onset of recruitment from North West Greater Manchester Central Research Ethics Committee 17/NW/0283.

Data monitoring was carried out during the trial by the study management team and the sponsor. Independent oversight of the study including interim safety monitoring was conducted by an Independent Data Monitoring and Ethics Committee.

### Randomisation and blinding

The randomisation process is described in section 6.4.3 of Appendix 1. Blinding procedures included participants and the study team being blinded to the randomisation process, mitigation of accidental unblinding, semi-blind interim safety analyses and independent Data Monitoring and Ethics Committee oversight. Dosing compliance was monitored by self-reported completion of diary cards and by counting the number of tablets remaining at each study visit (see Materials and Methods). Further detail is provided in section 7 of Appendix 2.

### Treatments and withdrawal criteria

Participants received either oral AZD4017 (400mg) or matched oral placebo, twice daily for 35 days. For full treatment and compliance details please see section 8 of Appendix 2. AZD4017 withdrawal criteria are detailed in section 6.5 of Appendix 2.

### Study endpoints

The primary endpoint was 24hr 11β-HSD1 activity in skin (efficacy) at baseline and day 28. Secondary endpoints were systemic 11β-HSD1 activity at baseline and day 35 and AZD4017 quantification in skin at day 28 and plasma at day 35 (to support efficacy), safety variables at baseline and days 7, 28 and 35, blood pressure, blood safety variables, EGC and biopsy inspection at day 42, urinary cortisol to cortisone metabolite analysis at baseline and day 35 (to assess systemic GC levels), skin function variables at baseline and days 2, 7, 28, 30 and 35 and feasibility variables at throughout the study. Further details are provided in sections 4, 9 and 10 of Appendix 2.

### Statistical Methods

A standalone statistical analysis plan (SAP) was developed and finalised prior to breaking of the blind and processing of primary outcome samples.

In this pilot study, the analysis was descriptive throughout and hence no specific inferential hypotheses were formally tested. Analysis followed published recommendations which state that in pilot trials the focus should be on descriptive statistics and estimation, using a range of confidence intervals (CIs) other than 95% confidence, and interpreting these with regards to the minimum clinically important difference (MCID)(*55*). As MCIDs have not been established in this patient group for the outcomes under investigation, we have converted the CI limits into % difference relative to the mean or median in the placebo group as a guideline when reporting results. Analyses were conducted in Stata 16.1 (StataCorp 2019. College Station, TX). For further details please refer to section 13 of Appendix 2 and Appendix 3.

## Supporting information

CONSORT extension for Pilot and Feasibility Trials Abstracts Checklist

CONSORT extension for Pilot and Feasibility Trials Checklist

## Data Availability

The authors confirm that the data supporting the findings of this study are available within the article and its supplementary materials.

## Author contributions

Ramzi Ajjan: Conceptualization, Methodology, Investigation, Writing - Review & Editing, Resources, Supervision (joint), Elizabeth Hensor: Software, Formal analysis, Data Curation, Writing - Review & Editing, Visualization, Kave Shams: Investigation, Francesco Del Galdo: Investigation, Afroze Abbas: Investigation, Janet Woods: Investigation, Ann Morgan: Supervision (supporting), Writing - Review & Editing, Paul M Stewart: Conceptualization, Funding acquisition, Writing - Review & Editing, Rebecca Fairclough: Resources, Supervision (supporting), Lorraine Webber: Resources, Supervision (supporting), Lindsay Pegg: Resources, Supervision (supporting), Adrian Freeman: Writing - Review & Editing, Resources, Supervision (supporting), Angela E Taylor: Investigation, Wiebke Arlt: Methodology, Abd Tahrani: Conceptualization, Writing - Review & Editing, David Russell: Investigation, Supervision (supporting) and Ana Tiganescu: Conceptualization (lead), Funding acquisition (lead), Investigation (lead), Methodology (lead), Project administration (lead), Supervision (joint), Validation, Writing - Original Draft, Writing - Review & Editing.

## Data and materials availability

All data associated with this study are available in the main text or the supplementary materials.

## Trial registration

International Standard Randomised Controlled Trial Number 74621291 www.isrctn.com/ISRCTN74621291. ClinicalTrials.gov Identifier: NCT03313297 https://clinicaltrials.gov/ct2/show/NCT03313297.

This paper presents independent research supported by the National Institute for Health Research (NIHR) Leeds Biomedical Research Centre (BRC). The views expressed are those of the authors and not necessarily those of the NIHR or the Department of Health and Social Care.

## Acronym

GC-SHEALD: Glucocorticoids and Skin Healing in Diabetes

## Supplementary Material

Table S1. Full descriptive data for primary and secondary efficacy variables. Population: Full analysis set.

Table S2. Full descriptive data for laboratory safety variables. Population: Safety set.

Table S3. IMP (placebo or AZD4017) compliance. Population: Full analysis set.

Figure S1. Correlation between AZD4017 levels in the biopsy taken at day 28 and in plasma taken at day 35. Population: Full analysis set (AZD4017 group only).

Table S4. Primary and secondary efficacy outcomes; unadjusted differences in final values between treatment groups. Population: Full analysis set. Multiple imputation was used to address missing data. Due to issues with the data distributions, for all variables except TEWL, integrity, wound diameter & depth, median regression was used to estimate confidence intervals around differences between the groups. For TEWL, integrity, wound depth and diameter, linear regression was used. TEWL and integrity measurements were log-transformed prior to analysis; differences have been expressed as ratios of geometric means (AZD:PCB).

Table S5. Sensitivity analysis of primary and secondary efficacy outcomes re-imputed after QC fails and outliers removed. Population: Full analysis set. Sensitivity analysis. Data were re-imputed after deleting two baseline radioassays which failed QC and one outlying ELISA assay result at day 28, TEWL readings that were potentially unreliable due to high temperatures and TEWL and WH measures which were collected earlier than scheduled. For each variable, the comparison was adjusted for baseline value of the variable (where applicable; not applicable to wound diameter & depth), age, sex and baseline HbA1c. All TEWL readings were adjusted using TEWL baseline at day 0.

Figure S2. Correlation between the different methods of assessing 24hr 11bHSD1 activity (% conv/24hr). Population: Full analysis set.

Table S6: Unadjusted differences in changes from baseline in primary and secondary efficacy outcomes. Population: Full analysis set. Multiple imputation was used to address missing data. Due to issues with the data distributions, for all variables except TEWL & integrity, median regression was used to estimate confidence intervals around differences between the groups. For TEWL & integrity, linear regression was used. TEWL and integrity measurements were log-transformed prior to analysis. For these variables changes from baseline have been expressed as ratios of follow-up measurements to baseline values; between-group differences have been expressed as ratios of geometric means (AZD:PCB).

Table S7: Adjusted differences in changes from baseline in primary and secondary efficacy outcomes. Population: Full analysis set. Multiple imputation was used to address missing data. Due to issues with the data distributions, for all variables except TEWL & integrity, median regression was used to estimate confidence intervals around differences between the groups. For TEWL & integrity, linear regression was used. TEWL and integrity measurements were log-transformed prior to analysis. For these variables changes from baseline have been expressed as ratios of follow-up measurements to baseline values; between-group differences have been expressed as ratios of geometric means (AZD:PCB). For each variable, the comparison was adjusted for baseline value of the variable, age, sex and baseline HbA1c.

Table S8: Sensitivity analysis of trans-epidermal water loss (TEWL) adjusting for exact timing of post-disruption measurements. Population: Full analysis set. Analysis models included exact measurement times to account for variation in timings; results have been provided for a) data imputed including all observed values and b) data re-imputed after deleting TEWL readings that were potentially unreliable due to high temperatures. Linear mixed models were used to model post-baseline TEWL readings (within each set) as a function of time since disruption. Quadratic functions were added to allow for nonlinear change over time. Baseline values and changes over time were permitted to vary between patients. TEWL measurements were log-transformed prior to analysis; differences have been expressed as ratios of geometric means (AZD:PCB). For each variable, the comparison was adjusted for baseline (day 0, hour 0) TEWL, age, sex and baseline HbA1c.

Table S9. Sensitivity analysis of primary and secondary efficacy outcomes in available case data. Population: Full analysis set. Due to issues with the data distributions, for all variables except TEWL, integrity, wound diameter & depth, quantile (median) regression was used to obtain point estimates and confidence intervals for differences between groups. For TEWL, integrity, wound depth and diameter, linear regression was used. TEWL and integrity measurements were log-transformed prior to analysis; differences have been expressed as ratios of geometric means (AZD:PBO). For each variable, the comparison was adjusted for baseline value of the variable (where applicable; not applicable to wound diameter & depth), age, sex and baseline HbA1c. All TEWL readings were adjusted using TEWL baseline at day 0.

Table S10. Sensitivity analysis of primary and secondary efficacy outcomes using last observation carried forward. Population: Full analysis set. Due to issues with the data distributions, for all variables except TEWL, integrity, wound diameter & depth, quantile (median) regression was used to obtain point estimates and confidence intervals for differences between groups. For TEWL, integrity, wound depth and diameter, linear regression was used. TEWL and integrity measurements were log-transformed prior to analysis; differences have been expressed as ratios of geometric means (AZD:PBO). For each variable, the comparison was adjusted for baseline value of the variable (where applicable; not applicable to wound diameter & depth), age, sex and baseline HbA1c. All TEWL readings were adjusted using TEWL baseline at day 0.Note that day 0 TEWL measurements were missed in error for 1 patient therefore data could only be carried forward for 27 patients.

Table S11. Correlations between AZD4017 compliance and efficacy outcomes. Population: Full analysis set (AZD group only). Absolute rho values >/= 0.3 were considered preliminary evidence of association. Sensitivity analysis excluded baseline radioimmunoassay samples that failed QC, an outlying ELISA assay result at day 28, TEWL readings recorded on very hot days, and results that were recorded 1 day earlier than planned, prior to imputation. Note that not all of these issues occurred in samples from the AZD group but could have altered imputed values for any incomplete variables.

Table S12. Unadjusted differences in final values of longitudinal laboratory safety variables. Population: Safety set. Multiple imputation was used to address missing data. All point estimates and confidence intervals estimated via linear regression.

Table S13. Unadjusted differences in changes from baseline in longitudinal laboratory safety variables. Population: Safety set. Multiple imputation was used to address missing data. All point estimates and confidence intervals estimated via linear regression.

Table S14. Adjusted differences in changes from baseline in longitudinal laboratory safety variables. Population: Safety set. Multiple imputation was used to address missing data. All point estimates and confidence intervals estimated via linear regression.

Table S15. Longitudinal laboratory safety variables; absolute and relative frequencies of values below or above normal limits. Population: Safety set

Table S16. Sample sizes for future trials. Based on estimates from available case data in full analysis set. At alpha=0.05 (5% significance), 1-Beta=0.90 (90% power), accounting for 10% drop-out, sample sizes for a range of substantive between-group differences are presented below. For TEWL and integrity variables, means and SDs are presented on the log scale. Note that for all variables, sample size has been based on mean and SD (and Pearson’s r where applicable), as specified in the statistical analysis plan; issues with distributions for some variables may reduce the accuracy of these estimates. Sample sizes presented here should be considered preliminary.

