## Supplementary material for "A randomised controlled pilot trial of oral 11β-HSD1 inhibitor AZD4017 for wound healing in adults with type 2 diabetes mellitus": Table S1

### Table S1: Full descriptive data for primary and secondary efficacy variables

Population: Full analysis set

| **Variable** | **Summary** | **PCB** | **AZD** |
| --- | --- | --- | --- |
|  |  | **N=14** | **N=14** |
| **Baseline** |  |  |  |
| 11bHSD1 activity radioassay (% conv/24hrs): Day 0 | Mean (SD) | 15.84 (6.80) | 13.73 (6.02) |
|  | Median (Q1, Q3) | 15.25 (11.60, 18.40) | 10.70 (9.40, 17.40) |
|  | Min, Max | 5.20, 29.30 | 7.80, 25.70 |
|  | Nn | 14 | 14 |
| 11bHSD1 activity ELISA (% conv/24hrs): Day 0 | Mean (SD) | 11.04 (8.06) | 6.56 (3.38) |
|  | Median (Q1, Q3) | 6.80 (5.50, 15.60) | 6.45 (4.20, 8.50) |
|  | Min, Max | 3.00, 26.90 | 1.40, 13.60 |
|  | Nn | 14 | 14 |
| Sudomotor function Left Hand (micro S): Day 0 | Mean (SD) | 54.14 (15.04) | 56.43 (17.49) |
|  | Median (Q1, Q3) | 54.00 (47.00, 63.00) | 57.50 (39.00, 71.00) |
|  | Min, Max | 18.00, 75.00 | 30.00, 81.00 |
|  | Nn | 14 | 14 |
| Sudomotor function Right Hand (micro S): Day 0 | Mean (SD) | 51.43 (15.91) | 54.29 (16.99) |
|  | Median (Q1, Q3) | 54.00 (47.00, 60.00) | 56.50 (41.00, 67.00) |
|  | Min, Max | 16.00, 73.00 | 22.00, 80.00 |
|  | Nn | 14 | 14 |
| Sudomotor function Hands (micro S): Day 0 | Mean (SD) | 52.79 (15.20) | 55.36 (17.13) |
|  | Median (Q1, Q3) | 53.75 (48.00, 60.50) | 56.75 (40.00, 70.50) |
|  | Min, Max | 17.00, 74.00 | 26.00, 80.50 |
|  | Nn | 14 | 14 |
| Sudomotor function Left Foot (micro S): Day 0 | Mean (SD) | 63.29 (17.18) | 73.93 (15.62) |
|  | Median (Q1, Q3) | 65.50 (46.00, 78.00) | 79.00 (63.00, 84.00) |
|  | Min, Max | 38.00, 87.00 | 34.00, 92.00 |
|  | Nn | 14 | 14 |
| Sudomotor function Right Foot (micro S): Day 0 | Mean (SD) | 63.57 (18.00) | 71.36 (18.94) |
|  | Median (Q1, Q3) | 69.50 (48.00, 79.00) | 77.00 (69.00, 82.00) |
|  | Min, Max | 37.00, 87.00 | 14.00, 88.00 |
|  | Nn | 14 | 14 |
| Sudomotor function Feet (micro S): Day 0 | Mean (SD) | 63.43 (17.49) | 72.64 (17.08) |
|  | Median (Q1, Q3) | 67.50 (44.50, 76.50) | 78.00 (64.00, 83.50) |
|  | Min, Max | 37.50, 87.00 | 24.00, 90.00 |
|  | Nn | 14 | 14 |
| Sudomotor function Overall (micro S): Day 0 | Mean (SD) | 58.11 (14.71) | 64.00 (15.11) |
|  | Median (Q1, Q3) | 61.00 (46.75, 67.75) | 66.63 (63.25, 71.25) |
|  | Min, Max | 27.25, 80.00 | 25.00, 84.00 |
|  | Nn | 14 | 14 |
| Skin hydration (A.U): Day 0 | Mean (SD) | 40.88 (7.79) | 40.79 (9.19) |
|  | Median (Q1, Q3) | 40.47 (34.50, 46.18) | 40.35 (36.68, 45.51) |
|  | Min, Max | 27.17, 54.06 | 20.58, 58.68 |
|  | Nn | 14 | 14 |
| Epidermal thickness (micro m): Day 0 | Mean (SD) | 61.30 (10.53) | 65.96 (9.81) |
|  | Median (Q1, Q3) | 62.76 (54.51, 69.22) | 65.70 (60.74, 69.32) |
|  | Min, Max | 37.18, 77.57 | 44.08, 86.48 |
|  | Nn | 14 | 14 |
| Cortisol (mcg/24h): Day 0 | Mean (SD) | 81.50 (49.27) | 70.50 (22.24) |
|  | Median (Q1, Q3) | 68.50 (40.00, 101.00) | 75.50 (48.00, 84.00) |
|  | Min, Max | 25.00, 202.00 | 39.00, 116.00 |
|  | Nn | 14 | 14 |
| Urinary [THF+alloTHF]/THE ratio: Day 0 | Mean (SD) | 1.06 (0.38) | 0.91 (0.22) |
|  | Median (Q1, Q3) | 0.96 (0.81, 1.23) | 0.97 (0.80, 1.06) |
|  | Min, Max | 0.50, 1.99 | 0.45, 1.24 |
|  | Nn | 14 | 14 |
| Hour 0 TEWL (Set 1; Day 0) | Mean (SD) | 8.96 (3.94) | 9.61 (3.73) |
|  | Geometric mean | 8.24 | 9.05 |
|  | Median (Q1, Q3) | 8.55 (5.10, 10.70) | 8.90 (6.80, 11.30) |
|  | Min, Max | 4.70, 17.60 | 5.50, 19.30 |
|  | Nn | 14 | 13 |
| N tapes required for barrier disruption: Day 0 | Mean (SD) | 47.14 (15.21) | 55.54 (26.52) |
|  | Geometric mean | 44.49 | 50.48 |
|  | Median (Q1, Q3) | 49.50 (33.00, 60.00) | 51.00 (42.00, 58.00) |
|  | Min, Max | 19.00, 66.00 | 22.00, 116.00 |
|  | Nn | 14 | 13 |
| **Follow-up** |  |  |  |
| Sudomotor function Left Hand (micro S): Day 35 | Mean (SD) | 56.46 (11.12) | 62.85 (14.35) |
|  | Median (Q1, Q3) | 58.00 (48.00, 63.00) | 64.00 (59.00, 72.00) |
|  | Min, Max | 39.00, 73.00 | 28.00, 81.00 |
|  | Nn | 13 | 13 |
| Sudomotor function Right Hand (micro S): Day 35 | Mean (SD) | 54.08 (13.32) | 59.92 (14.79) |
|  | Median (Q1, Q3) | 55.00 (51.00, 60.00) | 63.00 (57.00, 69.00) |
|  | Min, Max | 29.00, 74.00 | 23.00, 77.00 |
|  | Nn | 13 | 13 |
| Sudomotor function Hands (micro S): Day 35 | Mean (SD) | 55.27 (11.97) | 61.38 (14.43) |
|  | Median (Q1, Q3) | 57.50 (49.50, 61.00) | 63.50 (59.00, 70.50) |
|  | Min, Max | 35.50, 71.50 | 25.50, 79.00 |
|  | Nn | 13 | 13 |
| Sudomotor function Left Foot (micro S): Day 35 | Mean (SD) | 68.69 (10.91) | 70.92 (18.61) |
|  | Median (Q1, Q3) | 70.00 (61.00, 76.00) | 80.00 (64.00, 82.00) |
|  | Min, Max | 49.00, 85.00 | 23.00, 88.00 |
|  | Nn | 13 | 13 |
| Sudomotor function Right Foot (micro S): Day 35 | Mean (SD) | 69.31 (9.87) | 70.31 (21.44) |
|  | Median (Q1, Q3) | 65.00 (63.00, 75.00) | 80.00 (68.00, 82.00) |
|  | Min, Max | 51.00, 85.00 | 10.00, 85.00 |
|  | Nn | 13 | 13 |
| Sudomotor function Feet (micro S): Day 35 | Mean (SD) | 69.00 (10.19) | 70.62 (19.92) |
|  | Median (Q1, Q3) | 68.50 (62.00, 75.00) | 80.00 (66.00, 82.50) |
|  | Min, Max | 50.00, 85.00 | 16.50, 84.50 |
|  | Nn | 13 | 13 |
| Sudomotor function Overall (micro S): Day 35 | Mean (SD) | 62.13 (10.01) | 66.00 (15.12) |
|  | Median (Q1, Q3) | 59.75 (53.75, 68.00) | 70.75 (64.50, 73.00) |
|  | Min, Max | 49.25, 77.50 | 21.00, 78.75 |
|  | Nn | 13 | 13 |
| Skin hydration (A.U): Day 35 | Mean (SD) | 38.19 (9.69) | 44.82 (10.10) |
|  | Median (Q1, Q3) | 40.52 (29.40, 46.04) | 45.28 (37.02, 46.53) |
|  | Min, Max | 23.74, 52.86 | 30.53, 69.72 |
|  | Nn | 12 | 12 |
| Epidermal thickness (micro m): Day 35 | Mean (SD) | 61.72 (8.35) | 66.07 (10.49) |
|  | Median (Q1, Q3) | 57.54 (55.69, 67.30) | 66.29 (60.57, 74.37) |
|  | Min, Max | 51.32, 77.40 | 49.13, 82.95 |
|  | Nn | 13 | 12 |
| Cortisol (mcg/24h): Day 35 | Mean (SD) | 81.62 (44.49) | 64.08 (22.04) |
|  | Median (Q1, Q3) | 55.00 (47.00, 120.00) | 62.00 (52.00, 73.00) |
|  | Min, Max | 29.00, 164.00 | 34.00, 124.00 |
|  | Nn | 13 | 13 |
| Urinary [THF+alloTHF]/THE ratio: Day 35 | Mean (SD) | 1.08 (0.36) | 0.10 (0.03) |
|  | Median (Q1, Q3) | 0.89 (0.88, 1.24) | 0.10 (0.08, 0.12) |
|  | Min, Max | 0.75, 2.07 | 0.04, 0.16 |
|  | Nn | 13 | 13 |
| Wound gap diameter (mm): Day 2 | Mean (SD) | 1.49 (0.72) | 0.98 (0.70) |
|  | Median (Q1, Q3) | 1.64 (1.05, 2.02) | 0.88 (0.52, 1.29) |
|  | Min, Max | 0.00, 2.51 | 0.00, 2.42 |
|  | Nn | 14 | 14 |
| Wound depth (mm): Day 7 | Mean (SD) | 0.60 (0.23) | 0.59 (0.16) |
|  | Median (Q1, Q3) | 0.66 (0.57, 0.77) | 0.59 (0.57, 0.68) |
|  | Min, Max | 0.00, 0.82 | 0.27, 0.86 |
|  | Nn | 14 | 14 |
| Wound gap diameter (mm): Day 30 | Mean (SD) | 1.44 (0.70) | 0.65 (0.49) |
|  | Median (Q1, Q3) | 1.41 (1.14, 1.98) | 0.81 (0.25, 0.98) |
|  | Min, Max | 0.00, 2.44 | 0.00, 1.43 |
|  | Nn | 11 | 12 |
| Wound depth (mm): Day 35 | Mean (SD) | 0.60 (0.17) | 0.54 (0.21) |
|  | Median (Q1, Q3) | 0.63 (0.55, 0.67) | 0.54 (0.47, 0.63) |
|  | Min, Max | 0.27, 0.90 | 0.19, 0.94 |
|  | Nn | 13 | 12 |
| Biopsy AZD4017: Day 28 | Mean (SD) | <5.00 (0.00) | 1685.07 (914.43) |
|  | Geometric mean | <5.00 | 1442.44 |
|  | Median (Q1, Q3) | <5.00 (<5.00, <5.00) | 1570.00 (876.00, 2440.00) |
|  | Min, Max | <5.00, <5.00 | 443.00, 3310.00 |
|  | Nn | 13 | 14 |
| Plasma AZD4017: Day 35 | Mean (SD) | <5.00 (0.00) | 6992.50 (5303.82) |
|  | Geometric mean | <5.00 | 5281.66 |
|  | Median (Q1, Q3) | <5.00 (<5.00, <5.00) | 6490.00 (2960.00, 9040.00) |
|  | Min, Max | <5.00, <5.00 | 1180.00, 19400.00 |
|  | Nn | 7 | 12 |
| Hour 3 TEWL (Set 1; Day 0) | Mean (SD) | 36.03 (9.66) | 31.68 (8.00) |
|  | Geometric mean | 34.94 | 30.84 |
|  | Median (Q1, Q3) | 32.50 (29.30, 43.20) | 31.50 (26.10, 35.50) |
|  | Min, Max | 26.30, 53.80 | 22.60, 50.70 |
|  | Nn | 14 | 12 |
| Hour 48 TEWL (Set 1; Day 2) | Mean (SD) | 20.53 (5.64) | 22.98 (10.04) |
|  | Geometric mean | 19.79 | 21.29 |
|  | Median (Q1, Q3) | 19.90 (16.10, 26.00) | 21.40 (15.40, 25.80) |
|  | Min, Max | 12.40, 29.80 | 11.40, 47.80 |
|  | Nn | 14 | 11 |
| Hour 168 TEWL (Set 1; Day 7) | Mean (SD) | 13.95 (3.52) | 21.22 (18.23) |
|  | Geometric mean | 13.55 | 16.06 |
|  | Median (Q1, Q3) | 14.90 (11.10, 15.90) | 14.50 (9.40, 24.70) |
|  | Min, Max | 9.00, 21.10 | 6.90, 60.60 |
|  | Nn | 13 | 13 |
| Hour 3 TEWL (Set 2; Day 28) | Mean (SD) | 26.35 (11.63) | 31.17 (9.81) |
|  | Geometric mean | 23.77 | 29.83 |
|  | Median (Q1, Q3) | 27.80 (19.50, 28.60) | 32.80 (21.40, 37.40) |
|  | Min, Max | 9.00, 52.10 | 18.70, 54.90 |
|  | Nn | 13 | 14 |
| Hour 48 TEWL (Set 2; Day 30) | Mean (SD) | 15.65 (5.99) | 19.64 (9.12) |
|  | Geometric mean | 14.43 | 18.21 |
|  | Median (Q1, Q3) | 15.30 (13.20, 19.50) | 16.80 (14.90, 19.60) |
|  | Min, Max | 6.50, 25.30 | 11.70, 40.50 |
|  | Nn | 13 | 14 |
| Hour 168 TEWL (Set 2; Day 35) | Mean (SD) | 10.72 (4.01) | 11.72 (4.83) |
|  | Geometric mean | 10.02 | 10.84 |
|  | Median (Q1, Q3) | 10.50 (8.20, 12.40) | 10.40 (8.80, 15.10) |
|  | Min, Max | 4.70, 18.20 | 4.90, 20.40 |
|  | Nn | 13 | 12 |
| Hour 0 TEWL (Set 3; Day 35) | Mean (SD) | 7.45 (2.82) | 9.78 (4.35) |
|  | Geometric mean | 6.99 | 8.87 |
|  | Median (Q1, Q3) | 7.00 (5.40, 9.10) | 9.70 (7.70, 12.50) |
|  | Min, Max | 3.60, 14.10 | 4.40, 18.30 |
|  | Nn | 13 | 12 |
| N tapes required for barrier disruption: Day 28 | Mean (SD) | 42.85 (19.79) | 60.14 (25.72) |
|  | Geometric mean | 37.88 | 55.55 |
|  | Median (Q1, Q3) | 40.00 (25.00, 54.00) | 55.00 (41.00, 70.00) |
|  | Min, Max | 11.00, 77.00 | 30.00, 120.00 |
|  | Nn | 13 | 14 |
| Nn=Number non-missing; Q1=1st quartile; Q3=3rd quartile | | | |
