## Supplementary material for "A randomised controlled pilot trial of oral 11β-HSD1 inhibitor AZD4017 for wound healing in adults with type 2 diabetes mellitus": Table S2

### Table S2: Full descriptive data for laboratory safety variables

Population: Safety set

| **Variable** | **Summary** | **Day 0** | | **Day 7** | | **Day 28** | | **Day 35** | | **Day 42** | |
| --- | --- | --- | --- | --- | --- | --- | --- | --- | --- | --- | --- |
|  |  | **PCB**  **N=14** | **AZD**  **N=14** | **PCB**  **N=14** | **AZD**  **N=14** | **PCB**  **N=14** | **AZD**  **N=14** | **PCB**  **N=14** | **AZD**  **N=14** | **PCB**  **N=14** | **AZD**  **N=14** |
| Body Mass Index (kg / m2) | Mean (SD) | 33.67 (13.47) | 35.05 (5.68) |  |  |  |  | 34.07 (14.56) | 35.71 (6.43) |  |  |
|  | Median | 31.02 | 34.54 |  |  |  |  | 31.35 | 35.06 |  |  |
|  | (Q1, Q3) | (26.52, 33.57) | (30.65, 39.08) |  |  |  |  | (26.51, 32.93) | (31.38, 38.61) |  |  |
|  | Min, Max | 22.59, 75.64 | 27.04, 46.54 |  |  |  |  | 22.85, 77.85 | 27.35, 51.80 |  |  |
|  | Nn | 14 | 14 |  |  |  |  | 13 | 13 |  |  |
| Waist-hip ratio | Mean (SD) | 0.98 (0.08) | 1.03 (0.08) |  |  |  |  | 0.98 (0.07) | 1.02 (0.07) |  |  |
|  | Median | 0.98 | 1.03 |  |  |  |  | 0.98 | 1.01 |  |  |
|  | (Q1, Q3) | (0.92, 1.05) | (0.95, 1.10) |  |  |  |  | (0.91, 1.04) | (0.97, 1.07) |  |  |
|  | Min, Max | 0.85, 1.13 | 0.92, 1.17 |  |  |  |  | 0.89, 1.09 | 0.92, 1.16 |  |  |
|  | Nn | 14 | 14 |  |  |  |  | 13 | 13 |  |  |
| Systolic blood pressure (mm Hg) | Mean (SD) | 135.71 (21.01) | 140.43 (12.02) |  |  |  |  | 143.54 (12.07) | 128.62 (11.23) | 136.92 (13.12) | 137.50 (10.97) |
|  | Median | 136.00 | 140.00 |  |  |  |  | 148.00 | 126.00 | 137.00 | 140.00 |
|  | (Q1, Q3) | (120.00, 145.00) | (131.00, 150.00) |  |  |  |  | (140.00, 152.00) | (123.00, 134.00) | (128.00, 146.00) | (129.00, 146.00) |
|  | Min, Max | 101.00, 174.00 | 120.00, 162.00 |  |  |  |  | 121.00, 159.00 | 106.00, 154.00 | 118.00, 158.00 | 113.00, 150.00 |
|  | Nn | 14 | 14 |  |  |  |  | 13 | 13 | 13 | 14 |
| Diastolic blood pressure (mm Hg) | Mean (SD) | 83.86 (8.91) | 72.64 (9.96) |  |  |  |  | 79.62 (7.11) | 73.69 (7.88) | 80.00 (10.04) | 79.29 (11.36) |
|  | Median | 83.00 | 74.00 |  |  |  |  | 80.00 | 73.00 | 80.00 | 77.00 |
|  | (Q1, Q3) | (77.00, 90.00) | (63.00, 84.00) |  |  |  |  | (75.00, 84.00) | (70.00, 78.00) | (77.00, 86.00) | (71.00, 89.00) |
|  | Min, Max | 68.00, 99.00 | 60.00, 86.00 |  |  |  |  | 67.00, 92.00 | 61.00, 91.00 | 59.00, 94.00 | 60.00, 103.00 |
|  | Nn | 14 | 14 |  |  |  |  | 13 | 13 | 13 | 14 |
| HbA1c (mmol/mol) | Mean (SD) | 72.29 (19.43) | 66.00 (14.91) | 73.77 (18.09) | 64.31 (14.60) | 68.92 (17.09) | 66.00 (15.66) | 70.00 (20.10) | 63.67 (16.27) | 68.46 (17.76) | 65.43 (16.31) |
|  | Median | 72.00 | 64.00 | 71.00 | 63.00 | 67.00 | 69.00 | 71.00 | 68.00 | 69.00 | 68.00 |
|  | (Q1, Q3) | (54.00, 90.00) | (59.00, 82.00) | (56.00, 90.00) | (57.00, 71.00) | (55.00, 85.00) | (54.00, 81.00) | (49.00, 86.00) | (52.00, 77.00) | (55.00, 81.00) | (53.00, 78.00) |
|  | Min, Max | 46.00, 100.00 | 42.00, 86.00 | 49.00, 98.00 | 41.00, 85.00 | 46.00, 98.00 | 42.00, 87.00 | 45.00, 108.00 | 42.00, 86.00 | 44.00, 109.00 | 42.00, 90.00 |
|  | Nn | 14 | 14 | 13 | 13 | 13 | 14 | 11 | 12 | 13 | 14 |
| High density lipoprotein (mmol/l) | Mean (SD) | 1.24 (0.31) | 1.19 (0.27) | 1.16 (0.29) | 1.05 (0.25) | 1.22 (0.30) | 1.11 (0.25) | 1.19 (0.30) | 0.98 (0.26) | 1.18 (0.36) | 1.22 (0.28) |
|  | Median | 1.20 | 1.20 | 1.10 | 1.10 | 1.10 | 1.10 | 1.10 | 0.90 | 1.20 | 1.30 |
|  | (Q1, Q3) | (1.00, 1.30) | (0.90, 1.40) | (0.90, 1.30) | (0.80, 1.20) | (1.10, 1.30) | (0.80, 1.30) | (1.10, 1.20) | (0.80, 1.10) | (1.00, 1.20) | (0.90, 1.40) |
|  | Min, Max | 0.90, 2.00 | 0.70, 1.60 | 0.80, 1.80 | 0.70, 1.40 | 0.90, 1.90 | 0.70, 1.40 | 0.90, 1.90 | 0.60, 1.40 | 0.50, 1.80 | 0.80, 1.70 |
|  | Nn | 14 | 14 | 14 | 14 | 13 | 14 | 12 | 12 | 12 | 13 |
| Cholesterol (mmol/l) | Mean (SD) | 4.36 (1.13) | 3.95 (0.75) | 4.31 (1.00) | 3.59 (0.64) | 4.15 (0.91) | 3.54 (0.59) | 4.17 (0.88) | 3.44 (0.64) | 4.06 (1.01) | 3.81 (0.61) |
|  | Median | 4.40 | 3.90 | 4.30 | 3.40 | 4.10 | 3.40 | 4.10 | 3.40 | 4.20 | 3.80 |
|  | (Q1, Q3) | (3.60, 4.60) | (3.40, 4.40) | (3.40, 4.50) | (3.20, 4.20) | (3.70, 4.50) | (3.20, 3.70) | (3.80, 4.60) | (3.00, 3.70) | (3.70, 4.40) | (3.40, 4.20) |
|  | Min, Max | 3.10, 7.50 | 2.60, 5.50 | 3.10, 6.70 | 2.60, 4.90 | 2.70, 6.30 | 2.60, 4.90 | 2.80, 6.20 | 2.70, 4.80 | 2.00, 6.00 | 2.80, 5.10 |
|  | Nn | 14 | 14 | 14 | 14 | 13 | 14 | 12 | 12 | 12 | 13 |
| Triglycerides (mmol/l) | Mean (SD) | 1.68 (0.74) | 2.02 (1.02) | 2.11 (0.70) | 2.39 (1.58) | 1.80 (0.86) | 1.71 (0.75) | 2.23 (1.02) | 2.38 (2.22) | 1.63 (0.78) | 1.89 (1.31) |
|  | Median | 1.60 | 1.50 | 2.00 | 2.00 | 1.50 | 1.60 | 2.00 | 1.80 | 1.70 | 1.60 |
|  | (Q1, Q3) | (1.10, 1.90) | (1.40, 2.50) | (1.70, 2.70) | (1.70, 2.30) | (1.20, 2.00) | (1.00, 2.00) | (1.60, 2.80) | (1.40, 2.10) | (1.00, 1.90) | (1.20, 2.00) |
|  | Min, Max | 1.10, 3.90 | 1.10, 4.70 | 0.70, 3.30 | 1.10, 7.10 | 1.00, 3.70 | 1.00, 3.40 | 0.80, 3.90 | 0.90, 8.90 | 0.70, 3.20 | 0.80, 6.00 |
|  | Nn | 14 | 14 | 14 | 14 | 13 | 14 | 12 | 12 | 12 | 13 |
| Haemoglobin (g/l) | Mean (SD) | 139.21 (14.12) | 139.43 (11.35) | 136.07 (13.04) | 137.43 (10.80) | 137.46 (10.11) | 138.36 (10.58) | 134.42 (12.91) | 136.31 (11.50) | 135.31 (13.19) | 139.86 (13.42) |
|  | Median | 138.00 | 139.00 | 136.00 | 135.00 | 138.00 | 136.00 | 133.00 | 134.00 | 139.00 | 140.00 |
|  | (Q1, Q3) | (130.00, 146.00) | (130.00, 145.00) | (126.00, 145.00) | (131.00, 144.00) | (132.00, 145.00) | (134.00, 144.00) | (126.00, 144.00) | (129.00, 140.00) | (130.00, 145.00) | (131.00, 145.00) |
|  | Min, Max | 112.00, 168.00 | 121.00, 162.00 | 115.00, 161.00 | 122.00, 163.00 | 113.00, 153.00 | 124.00, 164.00 | 110.00, 155.00 | 122.00, 162.00 | 106.00, 151.00 | 117.00, 162.00 |
|  | Nn | 14 | 14 | 14 | 14 | 13 | 14 | 12 | 13 | 13 | 14 |
| White cells (x109/l) | Mean (SD) | 6.46 (2.55) | 7.34 (2.04) | 6.71 (1.83) | 7.97 (2.12) | 6.54 (2.11) | 6.72 (1.57) | 6.69 (1.96) | 7.04 (1.94) | 6.51 (1.97) | 7.66 (2.18) |
|  | Median | 6.62 | 7.36 | 6.19 | 8.40 | 6.44 | 6.63 | 7.01 | 7.09 | 6.42 | 7.69 |
|  | (Q1, Q3) | (4.59, 8.33) | (6.09, 8.74) | (5.41, 8.00) | (6.47, 9.54) | (5.81, 7.48) | (6.05, 7.43) | (5.06, 7.78) | (5.85, 8.59) | (5.73, 6.82) | (7.00, 8.68) |
|  | Min, Max | 2.13, 11.62 | 3.21, 11.27 | 4.11, 10.35 | 3.83, 10.91 | 2.33, 10.21 | 3.30, 9.34 | 3.42, 10.49 | 3.58, 9.90 | 3.19, 10.24 | 3.65, 12.00 |
|  | Nn | 14 | 14 | 14 | 14 | 13 | 14 | 12 | 13 | 13 | 14 |
| Platelets (x109/l) | Mean (SD) | 218.71 (61.85) | 273.14 (81.37) | 219.36 (64.89) | 275.29 (81.60) | 222.69 (59.57) | 288.57 (99.85) | 215.92 (66.59) | 272.69 (85.96) | 221.77 (60.01) | 281.43 (92.00) |
|  | Median | 204.00 | 264.00 | 214.00 | 264.00 | 190.00 | 286.00 | 204.00 | 260.00 | 203.00 | 270.00 |
|  | (Q1, Q3) | (171.00, 259.00) | (232.00, 335.00) | (171.00, 264.00) | (223.00, 317.00) | (180.00, 262.00) | (221.00, 331.00) | (171.00, 212.00) | (245.00, 285.00) | (170.00, 254.00) | (205.00, 349.00) |
|  | Min, Max | 149.00, 352.00 | 117.00, 402.00 | 146.00, 381.00 | 130.00, 428.00 | 146.00, 325.00 | 124.00, 476.00 | 149.00, 365.00 | 136.00, 442.00 | 155.00, 357.00 | 133.00, 446.00 |
|  | Nn | 14 | 14 | 14 | 14 | 13 | 14 | 12 | 13 | 13 | 14 |
| Red cells (x1012/l) | Mean (SD) | 4.84 (0.49) | 4.71 (0.48) | 4.75 (0.54) | 4.59 (0.45) | 4.77 (0.39) | 4.68 (0.47) | 4.66 (0.47) | 4.57 (0.44) | 4.68 (0.49) | 4.77 (0.53) |
|  | Median | 4.80 | 4.65 | 4.79 | 4.54 | 4.70 | 4.65 | 4.65 | 4.58 | 4.55 | 4.76 |
|  | (Q1, Q3) | (4.46, 5.17) | (4.56, 5.05) | (4.27, 5.11) | (4.39, 4.87) | (4.52, 4.89) | (4.44, 4.78) | (4.25, 5.01) | (4.38, 4.81) | (4.41, 4.98) | (4.59, 4.92) |
|  | Min, Max | 4.13, 5.78 | 3.59, 5.68 | 3.93, 5.66 | 3.57, 5.53 | 4.24, 5.62 | 3.74, 5.60 | 4.01, 5.42 | 3.57, 5.43 | 3.96, 5.53 | 3.62, 5.97 |
|  | Nn | 14 | 14 | 14 | 14 | 13 | 14 | 12 | 13 | 13 | 14 |
| Mean corpuscular volume (fl) | Mean (SD) | 88.43 (4.64) | 91.21 (6.96) | 89.71 (4.39) | 92.14 (6.77) | 88.15 (5.41) | 91.21 (6.82) | 90.00 (6.30) | 92.23 (6.73) | 88.08 (4.57) | 90.50 (6.10) |
|  | Median | 88.00 | 91.00 | 90.00 | 93.00 | 87.00 | 91.00 | 91.00 | 94.00 | 88.00 | 91.00 |
|  | (Q1, Q3) | (87.00, 91.00) | (86.00, 95.00) | (89.00, 92.00) | (88.00, 95.00) | (86.00, 92.00) | (84.00, 97.00) | (86.00, 93.00) | (89.00, 95.00) | (85.00, 92.00) | (84.00, 95.00) |
|  | Min, Max | 79.00, 97.00 | 80.00, 107.00 | 80.00, 98.00 | 81.00, 105.00 | 79.00, 98.00 | 82.00, 105.00 | 76.00, 99.00 | 80.00, 104.00 | 80.00, 96.00 | 81.00, 101.00 |
|  | Nn | 14 | 14 | 14 | 14 | 13 | 14 | 12 | 13 | 13 | 14 |
| Haematocrit (packed cell volume) | Mean (SD) | 0.43 (0.04) | 0.43 (0.03) | 0.43 (0.04) | 0.42 (0.03) | 0.42 (0.03) | 0.43 (0.03) | 0.42 (0.04) | 0.42 (0.04) | 0.41 (0.03) | 0.43 (0.04) |
|  | Median | 0.42 | 0.43 | 0.42 | 0.41 | 0.41 | 0.42 | 0.42 | 0.41 | 0.40 | 0.43 |
|  | (Q1, Q3) | (0.40, 0.46) | (0.41, 0.45) | (0.38, 0.46) | (0.40, 0.43) | (0.40, 0.43) | (0.40, 0.45) | (0.39, 0.45) | (0.40, 0.43) | (0.39, 0.45) | (0.40, 0.45) |
|  | Min, Max | 0.37, 0.50 | 0.37, 0.48 | 0.37, 0.50 | 0.37, 0.49 | 0.37, 0.48 | 0.38, 0.50 | 0.35, 0.48 | 0.37, 0.49 | 0.36, 0.47 | 0.36, 0.50 |
|  | Nn | 14 | 14 | 14 | 14 | 13 | 14 | 12 | 13 | 13 | 14 |
| Mean corpuscular haemoglobin (pg) | Mean (SD) | 28.82 (2.09) | 29.74 (2.47) | 28.79 (1.99) | 30.13 (2.50) | 28.91 (2.07) | 29.68 (2.12) | 28.93 (2.14) | 29.96 (2.56) | 28.98 (2.34) | 29.42 (2.15) |
|  | Median | 28.70 | 30.00 | 28.80 | 30.10 | 29.60 | 30.00 | 29.10 | 30.10 | 29.10 | 29.70 |
|  | (Q1, Q3) | (27.70, 29.90) | (27.30, 31.50) | (27.60, 29.90) | (27.70, 32.00) | (27.70, 29.90) | (28.20, 31.10) | (28.00, 30.10) | (27.70, 31.30) | (28.10, 30.70) | (27.70, 30.90) |
|  | Min, Max | 23.70, 31.80 | 25.60, 34.50 | 24.60, 32.30 | 26.40, 35.00 | 24.00, 32.00 | 26.10, 33.40 | 23.70, 32.40 | 26.40, 34.50 | 23.60, 32.80 | 26.20, 33.10 |
|  | Nn | 14 | 14 | 14 | 14 | 13 | 14 | 12 | 13 | 13 | 14 |
| Corpuscular hemoglobin concentration (g/l) | Mean (SD) | 325.57 (10.43) | 326.64 (8.29) | 321.07 (11.87) | 326.93 (9.03) | 328.62 (13.61) | 325.79 (7.76) | 322.08 (11.20) | 325.08 (8.65) | 328.85 (15.56) | 324.86 (6.69) |
|  | Median | 328.00 | 328.00 | 321.00 | 327.00 | 333.00 | 326.00 | 324.00 | 329.00 | 333.00 | 326.00 |
|  | (Q1, Q3) | (322.00, 334.00) | (320.00, 332.00) | (311.00, 327.00) | (316.00, 334.00) | (319.00, 337.00) | (321.00, 328.00) | (311.00, 330.00) | (320.00, 330.00) | (320.00, 338.00) | (318.00, 330.00) |
|  | Min, Max | 302.00, 337.00 | 315.00, 345.00 | 307.00, 343.00 | 314.00, 344.00 | 306.00, 347.00 | 314.00, 344.00 | 303.00, 337.00 | 306.00, 334.00 | 294.00, 350.00 | 315.00, 336.00 |
|  | Nn | 14 | 14 | 14 | 14 | 13 | 14 | 12 | 13 | 13 | 14 |
| Red blood cell distribution width (%) | Mean (SD) | 13.88 (0.98) | 14.12 (1.10) | 14.43 (1.29) | 14.27 (1.12) | 14.02 (0.87) | 14.08 (0.82) | 14.23 (1.01) | 14.44 (0.88) | 13.80 (0.98) | 13.74 (1.04) |
|  | Median | 13.90 | 13.90 | 14.50 | 13.90 | 13.90 | 14.00 | 14.30 | 14.20 | 13.90 | 13.60 |
|  | (Q1, Q3) | (13.10, 14.50) | (13.20, 15.30) | (13.90, 14.80) | (13.70, 14.70) | (13.60, 14.30) | (13.80, 14.60) | (13.50, 14.40) | (14.00, 14.90) | (13.10, 14.10) | (13.40, 14.40) |
|  | Min, Max | 12.40, 16.00 | 12.70, 15.80 | 12.70, 17.70 | 12.90, 16.80 | 12.50, 16.00 | 12.00, 15.30 | 13.20, 16.60 | 12.70, 15.90 | 12.50, 16.30 | 11.70, 15.70 |
|  | Nn | 14 | 14 | 14 | 14 | 13 | 14 | 12 | 13 | 13 | 14 |
| Albumin (g/l) | Mean (SD) | 39.43 (2.56) | 39.50 (2.44) | 39.50 (2.53) | 40.14 (3.23) | 38.85 (2.48) | 38.71 (2.89) | 39.67 (2.27) | 40.00 (3.00) | 38.15 (1.95) | 38.69 (2.18) |
|  | Median | 39.00 | 40.00 | 40.00 | 40.00 | 39.00 | 39.00 | 40.00 | 39.00 | 38.00 | 39.00 |
|  | (Q1, Q3) | (38.00, 41.00) | (38.00, 41.00) | (38.00, 41.00) | (38.00, 42.00) | (37.00, 41.00) | (37.00, 40.00) | (38.00, 41.00) | (38.00, 41.00) | (37.00, 40.00) | (36.00, 40.00) |
|  | Min, Max | 35.00, 44.00 | 36.00, 44.00 | 35.00, 44.00 | 36.00, 49.00 | 34.00, 42.00 | 35.00, 46.00 | 36.00, 44.00 | 37.00, 46.00 | 35.00, 42.00 | 36.00, 42.00 |
|  | Nn | 14 | 14 | 14 | 14 | 13 | 14 | 12 | 13 | 13 | 13 |
| Blirubin (umol/l) | Mean (SD) | 8.93 (2.97) | 8.14 (2.82) | 8.36 (2.17) | 8.64 (5.53) | 9.69 (3.35) | 9.86 (4.49) | 8.92 (5.55) | 8.54 (3.80) | 8.31 (2.14) | 8.38 (2.66) |
|  | Median | 9.00 | 8.00 | 9.00 | 7.00 | 10.00 | 9.00 | 7.00 | 8.00 | 8.00 | 8.00 |
|  | (Q1, Q3) | (6.00, 12.00) | (6.00, 9.00) | (7.00, 10.00) | (6.00, 8.00) | (7.00, 11.00) | (7.00, 12.00) | (6.00, 9.00) | (7.00, 10.00) | (7.00, 11.00) | (7.00, 10.00) |
|  | Min, Max | 5.00, 14.00 | 4.00, 14.00 | 4.00, 12.00 | 4.00, 24.00 | 5.00, 16.00 | 5.00, 22.00 | 4.00, 24.00 | 4.00, 19.00 | 5.00, 11.00 | 5.00, 13.00 |
|  | Nn | 14 | 14 | 14 | 14 | 13 | 14 | 12 | 13 | 13 | 13 |
| Alkaline phosphatase (U/l) | Mean (SD) | 80.43 (23.76) | 85.07 (26.08) | 80.86 (27.04) | 80.14 (26.38) | 77.31 (21.55) | 68.93 (23.03) | 79.58 (21.21) | 66.15 (21.10) | 76.85 (25.66) | 78.62 (30.06) |
|  | Median | 77.00 | 88.00 | 72.00 | 79.00 | 72.00 | 70.00 | 76.00 | 69.00 | 69.00 | 79.00 |
|  | (Q1, Q3) | (65.00, 95.00) | (64.00, 106.00) | (66.00, 96.00) | (57.00, 102.00) | (64.00, 86.00) | (55.00, 80.00) | (71.00, 86.00) | (55.00, 75.00) | (65.00, 77.00) | (61.00, 91.00) |
|  | Min, Max | 50.00, 145.00 | 37.00, 123.00 | 49.00, 147.00 | 38.00, 122.00 | 51.00, 129.00 | 29.00, 114.00 | 47.00, 130.00 | 27.00, 100.00 | 50.00, 145.00 | 33.00, 152.00 |
|  | Nn | 14 | 14 | 14 | 14 | 13 | 14 | 12 | 13 | 13 | 13 |
| Alanine aminotransferase (iu/l) | Mean (SD) | 24.64 (7.10) | 27.43 (10.91) | 22.93 (6.83) | 26.14 (9.72) | 22.69 (6.59) | 23.07 (9.39) | 21.67 (6.36) | 21.15 (10.36) | 21.15 (6.40) | 24.31 (10.86) |
|  | Median | 25.00 | 25.00 | 23.00 | 25.00 | 20.00 | 22.00 | 21.00 | 18.00 | 19.00 | 22.00 |
|  | (Q1, Q3) | (19.00, 31.00) | (20.00, 33.00) | (19.00, 30.00) | (21.00, 27.00) | (19.00, 28.00) | (16.00, 30.00) | (18.00, 25.00) | (15.00, 23.00) | (18.00, 24.00) | (15.00, 29.00) |
|  | Min, Max | 15.00, 36.00 | 15.00, 51.00 | 12.00, 34.00 | 14.00, 53.00 | 12.00, 33.00 | 11.00, 41.00 | 14.00, 33.00 | 10.00, 46.00 | 12.00, 34.00 | 13.00, 45.00 |
|  | Nn | 14 | 14 | 14 | 14 | 13 | 14 | 12 | 13 | 13 | 13 |
| Aspartate aminotransferase (iu/l) | Mean (SD) | 21.00 (4.95) | 22.64 (5.50) | 21.07 (5.93) | 22.36 (3.99) | 20.77 (4.17) | 21.36 (2.68) | 20.92 (3.85) | 20.67 (2.93) | 19.00 (5.70) | 22.18 (3.76) |
|  | Median | 21.00 | 22.00 | 20.00 | 22.00 | 21.00 | 21.00 | 20.00 | 21.00 | 20.00 | 22.00 |
|  | (Q1, Q3) | (19.00, 22.00) | (19.00, 26.00) | (17.00, 24.00) | (19.00, 24.00) | (19.00, 23.00) | (19.00, 23.00) | (19.00, 21.00) | (19.00, 23.00) | (16.00, 20.00) | (21.00, 23.00) |
|  | Min, Max | 14.00, 32.00 | 14.00, 36.00 | 14.00, 36.00 | 18.00, 30.00 | 14.00, 28.00 | 18.00, 27.00 | 15.00, 30.00 | 16.00, 25.00 | 12.00, 34.00 | 15.00, 30.00 |
|  | Nn | 14 | 14 | 14 | 14 | 13 | 14 | 12 | 12 | 12 | 11 |
| Gamma-glutamyl transpeptidase (iu/l) | Mean (SD) | 41.86 (33.87) | 39.29 (29.87) | 39.71 (32.51) | 37.71 (30.94) | 42.46 (31.59) | 29.86 (21.31) | 43.67 (36.88) | 23.92 (15.94) | 41.23 (33.38) | 29.23 (20.66) |
|  | Median | 37.00 | 29.00 | 37.00 | 22.00 | 34.00 | 21.00 | 41.00 | 19.00 | 40.00 | 24.00 |
|  | (Q1, Q3) | (21.00, 45.00) | (21.00, 43.00) | (19.00, 46.00) | (20.00, 46.00) | (22.00, 58.00) | (17.00, 33.00) | (19.00, 47.00) | (15.00, 24.00) | (22.00, 42.00) | (18.00, 35.00) |
|  | Min, Max | 13.00, 145.00 | 16.00, 127.00 | 12.00, 143.00 | 13.00, 129.00 | 16.00, 133.00 | 14.00, 94.00 | 15.00, 147.00 | 10.00, 71.00 | 15.00, 143.00 | 6.00, 89.00 |
|  | Nn | 14 | 14 | 14 | 14 | 13 | 14 | 12 | 13 | 13 | 13 |
| eGFR (ml/min/1.73m2) | Mean (SD) | 81.86 (9.69) | 79.21 (13.59) | 77.50 (12.37) | 71.50 (15.24) | 79.77 (11.13) | 75.14 (14.09) | 76.83 (15.91) | 73.31 (16.69) | 77.46 (11.60) | 76.21 (14.73) |
|  | Median | 87.00 | 85.00 | 78.00 | 70.00 | 83.00 | 72.00 | 78.00 | 69.00 | 78.00 | 77.00 |
|  | (Q1, Q3) | (76.00, 90.00) | (70.00, 90.00) | (67.00, 90.00) | (55.00, 88.00) | (71.00, 90.00) | (67.00, 90.00) | (73.00, 90.00) | (62.00, 90.00) | (67.00, 90.00) | (69.00, 90.00) |
|  | Min, Max | 61.00, 90.00 | 54.00, 90.00 | 55.00, 90.00 | 47.00, 90.00 | 58.00, 90.00 | 51.00, 90.00 | 39.00, 90.00 | 47.00, 90.00 | 59.00, 90.00 | 42.00, 90.00 |
|  | Nn | 14 | 14 | 14 | 14 | 13 | 14 | 12 | 13 | 13 | 14 |
| Sodium (mmol/l) | Mean (SD) | 138.93 (2.06) | 140.71 (5.47) | 139.29 (2.30) | 140.00 (2.48) | 138.15 (1.82) | 138.79 (2.15) | 140.25 (3.47) | 139.31 (1.89) | 138.92 (2.02) | 138.00 (2.15) |
|  | Median | 139.00 | 140.00 | 139.00 | 140.00 | 138.00 | 139.00 | 140.00 | 139.00 | 139.00 | 138.00 |
|  | (Q1, Q3) | (137.00, 140.00) | (137.00, 142.00) | (138.00, 142.00) | (139.00, 141.00) | (137.00, 139.00) | (137.00, 140.00) | (138.00, 142.00) | (138.00, 140.00) | (138.00, 140.00) | (137.00, 139.00) |
|  | Min, Max | 136.00, 143.00 | 136.00, 158.00 | 135.00, 143.00 | 136.00, 144.00 | 135.00, 141.00 | 134.00, 142.00 | 136.00, 148.00 | 136.00, 143.00 | 135.00, 142.00 | 134.00, 142.00 |
|  | Nn | 14 | 14 | 14 | 14 | 13 | 14 | 12 | 13 | 13 | 14 |
| Potassium (mmol/l) | Mean (SD) | 4.46 (0.33) | 4.63 (0.41) | 4.79 (0.55) | 4.59 (0.37) | 4.49 (0.31) | 4.74 (0.41) | 4.48 (0.43) | 4.52 (0.28) | 4.65 (0.45) | 4.63 (0.33) |
|  | Median | 4.40 | 4.60 | 4.70 | 4.60 | 4.60 | 4.70 | 4.50 | 4.50 | 4.70 | 4.70 |
|  | (Q1, Q3) | (4.20, 4.70) | (4.40, 4.80) | (4.50, 5.00) | (4.30, 4.90) | (4.40, 4.70) | (4.40, 5.10) | (4.30, 4.80) | (4.30, 4.60) | (4.20, 4.90) | (4.60, 4.70) |
|  | Min, Max | 3.90, 5.10 | 3.90, 5.50 | 3.70, 5.70 | 4.10, 5.30 | 3.80, 4.90 | 4.20, 5.40 | 3.50, 5.00 | 4.20, 5.10 | 3.90, 5.50 | 4.10, 5.10 |
|  | Nn | 14 | 14 | 14 | 14 | 13 | 14 | 12 | 13 | 13 | 12 |
| Urea (mmol/l) | Mean (SD) | 6.96 (2.81) | 7.25 (2.17) | 6.91 (2.72) | 7.24 (1.89) | 6.79 (3.85) | 6.56 (1.80) | 7.67 (5.13) | 7.60 (2.72) | 7.07 (2.98) | 7.57 (2.72) |
|  | Median | 6.70 | 6.40 | 6.40 | 6.60 | 5.80 | 6.60 | 6.30 | 8.50 | 6.80 | 7.30 |
|  | (Q1, Q3) | (5.20, 7.30) | (5.90, 8.70) | (5.10, 8.90) | (5.60, 9.20) | (4.90, 6.80) | (5.30, 7.90) | (5.80, 7.40) | (5.60, 9.30) | (5.50, 7.30) | (6.10, 8.30) |
|  | Min, Max | 3.90, 14.90 | 3.90, 11.00 | 2.60, 12.30 | 4.90, 10.60 | 3.10, 18.60 | 3.80, 9.50 | 2.70, 23.10 | 3.70, 13.30 | 3.00, 14.70 | 3.90, 15.40 |
|  | Nn | 14 | 14 | 14 | 14 | 13 | 14 | 12 | 13 | 13 | 14 |
| Creatinine (umol/l) | Mean (SD) | 76.64 (15.36) | 76.21 (20.04) | 82.36 (17.90) | 88.57 (22.57) | 79.08 (16.81) | 80.93 (19.07) | 84.42 (24.67) | 84.69 (21.53) | 82.08 (16.96) | 79.93 (17.93) |
|  | Median | 77.00 | 76.00 | 82.00 | 84.00 | 81.00 | 75.00 | 81.00 | 74.00 | 82.00 | 75.00 |
|  | (Q1, Q3) | (66.00, 88.00) | (60.00, 94.00) | (70.00, 97.00) | (71.00, 102.00) | (69.00, 93.00) | (67.00, 93.00) | (70.00, 87.00) | (72.00, 101.00) | (66.00, 96.00) | (70.00, 95.00) |
|  | Min, Max | 49.00, 101.00 | 51.00, 115.00 | 55.00, 114.00 | 61.00, 129.00 | 55.00, 106.00 | 54.00, 120.00 | 55.00, 148.00 | 59.00, 125.00 | 59.00, 110.00 | 56.00, 111.00 |
|  | Nn | 14 | 14 | 14 | 14 | 13 | 14 | 12 | 13 | 13 | 14 |
| Testosterone (nmol/l) | Mean (SD) | 11.10 (6.25) | 6.97 (5.48) | 10.34 (7.47) | 6.43 (4.94) | 11.72 (6.75) | 6.46 (5.01) | 10.17 (7.94) | 6.03 (5.08) | 10.12 (6.47) | 7.66 (6.43) |
|  | Median | 10.10 | 8.00 | 10.70 | 7.00 | 11.50 | 6.70 | 9.30 | 5.00 | 9.70 | 8.10 |
|  | (Q1, Q3) | (8.00, 16.80) | (0.80, 11.40) | (6.20, 13.70) | (1.40, 9.90) | (8.50, 14.70) | (1.00, 10.30) | (8.00, 10.60) | (1.00, 10.00) | (6.90, 13.80) | (0.70, 12.30) |
|  | Min, Max | 0.60, 22.00 | 0.40, 15.00 | 0.50, 27.50 | 0.70, 14.90 | 0.60, 27.20 | 0.80, 14.20 | 0.50, 31.50 | 0.60, 13.50 | 0.40, 25.00 | 0.60, 18.30 |
|  | Nn | 14 | 14 | 14 | 14 | 12 | 14 | 12 | 13 | 13 | 14 |
| Dehydroepiandrosterone sulphate (umol/l) | Mean (SD) | 3.53 (2.19) | 2.91 (1.69) | 3.56 (2.36) | 4.91 (3.53) | 3.03 (1.86) | 5.35 (2.76) | 3.49 (1.90) | 5.03 (3.22) | 2.82 (1.70) | 3.55 (2.16) |
|  | Median | 2.90 | 2.50 | 3.60 | 3.50 | 3.20 | 5.30 | 3.80 | 4.60 | 2.70 | 3.40 |
|  | (Q1, Q3) | (1.00, 5.00) | (1.90, 3.40) | (1.00, 5.30) | (2.90, 5.50) | (1.20, 4.60) | (3.40, 6.70) | (2.30, 4.90) | (2.90, 5.40) | (1.10, 3.40) | (2.00, 4.30) |
|  | Min, Max | 1.00, 7.30 | 1.00, 7.90 | 1.00, 8.00 | 1.00, 14.70 | 1.00, 6.20 | 1.50, 11.60 | 1.00, 6.40 | 1.60, 12.60 | 1.00, 6.30 | 1.10, 9.70 |
|  | Nn | 14 | 14 | 14 | 14 | 13 | 14 | 12 | 13 | 13 | 14 |
| Free thyroxine (pmol/l) | Mean (SD) | 14.86 (2.20) | 15.73 (1.58) | 14.63 (1.72) | 15.70 (2.53) | 14.66 (1.61) | 15.72 (1.92) | 14.46 (1.43) | 15.39 (1.98) | 13.94 (1.47) | 15.10 (1.49) |
|  | Median | 15.20 | 15.20 | 14.70 | 15.50 | 14.60 | 16.20 | 14.60 | 14.80 | 14.00 | 15.10 |
|  | (Q1, Q3) | (13.10, 16.40) | (14.80, 16.50) | (13.20, 15.30) | (13.90, 16.80) | (13.40, 15.60) | (14.30, 17.30) | (13.50, 14.90) | (14.40, 17.60) | (12.60, 15.00) | (13.80, 16.10) |
|  | Min, Max | 11.20, 18.80 | 14.00, 20.00 | 12.10, 18.50 | 12.20, 21.50 | 12.60, 17.30 | 13.10, 19.00 | 12.70, 17.90 | 12.30, 18.40 | 12.10, 16.60 | 13.00, 18.80 |
|  | Nn | 14 | 14 | 14 | 13 | 12 | 12 | 12 | 13 | 13 | 14 |
| Thyroid stimulating hormone (mlU/l) | Mean (SD) | 1.74 (0.66) | 1.72 (0.63) | 1.68 (0.56) | 1.88 (0.76) | 1.91 (0.84) | 1.76 (0.81) | 1.76 (0.75) | 1.72 (0.64) | 1.79 (0.71) | 1.76 (0.93) |
|  | Median | 1.40 | 1.60 | 1.60 | 1.70 | 1.70 | 1.60 | 1.60 | 1.60 | 1.70 | 1.60 |
|  | (Q1, Q3) | (1.30, 2.40) | (1.20, 2.10) | (1.30, 2.00) | (1.30, 2.00) | (1.20, 2.40) | (1.30, 2.10) | (1.20, 2.00) | (1.50, 1.90) | (1.30, 2.10) | (1.40, 2.10) |
|  | Min, Max | 0.78, 2.80 | 0.94, 3.30 | 0.95, 2.90 | 0.97, 3.70 | 1.10, 3.50 | 0.68, 3.70 | 0.98, 3.40 | 0.62, 3.10 | 0.81, 3.00 | 0.51, 3.90 |
|  | Nn | 14 | 14 | 14 | 13 | 12 | 12 | 12 | 13 | 13 | 14 |
