## Supplementary material for "A randomised controlled pilot trial of oral 11β-HSD1 inhibitor AZD4017 for wound healing in adults with type 2 diabetes mellitus": Table S3

### Table S3: IMP (placebo or AZD4017) compliance

Population: Full analysis set

| **Definition** | **Summary** | **Day 2** | | **Day 7** | | **Day 28** | | **Day 30** | | **Day 35** | |
| --- | --- | --- | --- | --- | --- | --- | --- | --- | --- | --- | --- |
|  |  | **PCB**  **N=14** | **AZD**  **N=14** | **PCB**  **N=14** | **AZD**  **N=14** | **PCB**  **N=14** | **AZD**  **N=14** | **PCB**  **N=14** | **AZD**  **N=14** | **PCB**  **N=14** | **AZD**  **N=14** |
| Percent | Mean (SD) | >99 ( 1) | >99 ( 1) | 99 ( 1) | 99 ( 2) | 99 ( 3) | 98 ( 2) | 97 ( 6) | 99 ( 2) | 98 ( 5) | 98 ( 2) |
|  | Median (Q1, Q3) | 100 (100, 100) | 100 (100, 100) | 100 ( 99, 100) | 100 ( 98, 100) | 99 ( 98, 100) | 99 ( 97, 100) | 99 ( 98, 100) | 99 ( 98, 100) | 100 ( 98, 100) | 99 ( 97, 100) |
|  | Min, Max | 97, 101 | 98, 100 | 96, 101 | 94, 101 | 91, 101 | 94, 100 | 81, 101 | 94, 101 | 84, 101 | 93, 101 |
|  | Nn | 13 | 10 | 13 | 14 | 12 | 13 | 13 | 11 | 12 | 14 |
| Cumulative | Mean (SD) | 95 ( 17) | 96 ( 12) | 97 ( 7) | 96 ( 10) | 98 ( 3) | 98 ( 2) | 97 ( 6) | 98 ( 2) | 98 ( 5) | 98 ( 2) |
| percent | Median (Q1, Q3) | 100 (100, 100) | 100 (100, 100) | 100 ( 96, 100) | 100 ( 89, 100) | 99 ( 97, 100) | 98 ( 96, 100) | 99 ( 98, 100) | 98 ( 98, 100) | 100 ( 98, 100) | 99 ( 97, 100) |
|  | Min, Max | 50, 125 | 63, 100 | 79, 107 | 71, 107 | 89, 102 | 93, 100 | 78, 102 | 93, 101 | 84, 101 | 93, 101 |
|  | Nn | 13 | 10 | 13 | 14 | 12 | 13 | 13 | 11 | 12 | 14 |
