## Supplementary material for "A randomised controlled pilot trial of oral 11β-HSD1 inhibitor AZD4017 for wound healing in adults with type 2 diabetes mellitus": Table S4

### Table S4: Primary and secondary efficacy outcomes; unadjusted differences in final values between treatment groups

Population: Full analysis set

Multiple imputation was used to address missing data. Due to issues with the data distributions, for all variables except TEWL, integrity, wound diameter & depth, median regression was used to estimate confidence intervals around differences between the groups. For TEWL, integrity, wound depth and diameter, linear regression was used. TEWL and integrity measurements were log-transformed prior to analysis; differences have been expressed as ratios of geometric means (AZD:PCB).

| **Variable** | **Median*** | | **Difference** | **Confidence interval** | | | | |
| --- | --- | --- | --- | --- | --- | --- | --- | --- |
|  | PCB n=14 | AZD n=14 | AZD-PCB | 75% | 80% | 85% | 90% | 95% |
| 11bHSD1 activity radioassay (% conv/24hrs): Day 28 | 12.18 | 12.70 | 0.52 | (-2.85, 3.90) | (-3.25, 4.30) | (-3.73, 4.78) | (-4.37, 5.42) | (-5.38, 6.43) |
| 11bHSD1 activity ELISA (% conv/24hrs): Day 28 | 5.84 | 4.30 | -1.54 | (-3.58, 0.49) | (-3.82, 0.73) | (-4.12, 1.03) | (-4.50, 1.41) | (-5.12, 2.03) |
| Sudomotor function Left Hand (micro S): Day 35 | 58.00 | 63.15 | 5.15 | (-2.52, 12.82) | (-3.42, 13.72) | (-4.52, 14.82) | (-5.98, 16.28) | (-8.28, 18.58) |
| Sudomotor function Right Hand (micro S): Day 35 | 54.80 | 62.60 | 7.80 | (0.35, 15.25) | (-0.53, 16.13) | (-1.60, 17.20) | (-3.02, 18.62) | (-5.26, 20.86) |
| Sudomotor function Hands (micro S): Day 35 | 57.02 | 62.80 | 5.78 | (-2.14, 13.69) | (-3.08, 14.63) | (-4.22, 15.77) | (-5.73, 17.28) | (-8.11, 19.66) |
| Sudomotor function Left Foot (micro S): Day 35 | 69.40 | 76.20 | 6.80 | (-1.72, 15.32) | (-2.73, 16.33) | (-3.95, 17.55) | (-5.57, 19.17) | (-8.13, 21.73) |
| Sudomotor function Right Foot (micro S): Day 35 | 70.10 | 79.05 | 8.95 | (0.86, 17.04) | (-0.10, 18.00) | (-1.27, 19.17) | (-2.82, 20.72) | (-5.27, 23.17) |
| Sudomotor function Feet (micro S): Day 35 | 69.95 | 79.53 | 9.57 | (1.75, 17.40) | (0.83, 18.32) | (-0.29, 19.44) | (-1.78, 20.93) | (-4.12, 23.27) |
| Sudomotor function Overall (micro S): Day 35 | 62.48 | 70.58 | 8.10 | (1.07, 15.13) | (0.24, 15.96) | (-0.77, 16.97) | (-2.11, 18.31) | (-4.22, 20.42) |
| Skin hydration (A.U): Day 35 | 40.17 | 45.39 | 5.22 | (-0.64, 11.07) | (-1.33, 11.76) | (-2.17, 12.60) | (-3.29, 13.72) | (-5.05, 15.48) |
| Epidermal thickness (micro m): Day 35 | 60.32 | 66.94 | 6.62 | (-1.16, 14.41) | (-2.09, 15.34) | (-3.23, 16.47) | (-4.74, 17.99) | (-7.16, 20.40) |
| Cortisol (mcg/24h): Day 35 | 63.25 | 62.40 | -0.85 | (-28.55, 26.85) | (-31.96, 30.26) | (-36.19, 34.49) | (-41.91, 40.21) | (-51.28, 49.58) |
| Urinary [THF+alloTHF]/THE ratio: Day 35 | 0.91 | 0.10 | -0.81 | (-0.90, -0.72) | (-0.91, -0.70) | (-0.92, -0.69) | (-0.94, -0.67) | (-0.97, -0.64) |
| **Variable** | **Mean*** | | **Difference** | **Confidence interval** | | | | |
|  | PCB n=14 | AZD n=14 | AZD-PCB | 75% | 80% | 85% | 90% | 95% |
| Wound gap diameter (mm): Day 2 | 1.49 | 0.98 | -0.51 | (-0.83, -0.20) | (-0.87, -0.16) | (-0.91, -0.11) | (-0.97, -0.05) | (-1.07, 0.04) |
| Wound depth (mm): Day 7 | 0.60 | 0.59 | -0.01 | (-0.10, 0.08) | (-0.11, 0.09) | (-0.12, 0.10) | (-0.14, 0.12) | (-0.17, 0.15) |
| Wound gap diameter (mm): Day 30 | 1.38 | 0.67 | -0.71 | (-1.02, -0.39) | (-1.06, -0.35) | (-1.10, -0.31) | (-1.16, -0.25) | (-1.26, -0.15) |
| Wound depth (mm): Day 35 | 0.61 | 0.54 | -0.06 | (-0.15, 0.02) | (-0.16, 0.04) | (-0.17, 0.05) | (-0.19, 0.06) | (-0.22, 0.09) |
| **Variable** | **Geometric mean*** | | **Ratio** | **Confidence interval** | | | | |
|  | PCB n=14 | AZD n=14 | AZD:PCB | 75% | 80% | 85% | 90% | 95% |
| Hour 3 TEWL (Set 1; Day 0) | 34.94 | 32.00 | 0.92 | (0.82, 1.03) | (0.80, 1.04) | (0.79, 1.06) | (0.77, 1.08) | (0.75, 1.12) |
| Hour 48 TEWL (Set 1; Day 2) | 19.79 | 21.37 | 1.08 | (0.92, 1.27) | (0.90, 1.29) | (0.88, 1.32) | (0.86, 1.36) | (0.82, 1.43) |
| Hour 168 TEWL (Set 1; Day 7) | 13.52 | 16.36 | 1.21 | (0.94, 1.56) | (0.91, 1.61) | (0.88, 1.67) | (0.84, 1.75) | (0.77, 1.89) |
| Hour 3 TEWL (Set 2; Day 28) | 23.52 | 29.83 | 1.27 | (1.05, 1.53) | (1.03, 1.57) | (1.00, 1.61) | (0.96, 1.67) | (0.91, 1.76) |
| Hour 48 TEWL (Set 2; Day 30) | 14.65 | 18.21 | 1.24 | (1.03, 1.49) | (1.01, 1.53) | (0.99, 1.57) | (0.95, 1.62) | (0.90, 1.72) |
| Hour 168 TEWL (Set 2; Day 35) | 9.98 | 10.88 | 1.09 | (0.90, 1.31) | (0.89, 1.34) | (0.86, 1.38) | (0.83, 1.43) | (0.79, 1.51) |
| Hour 0 TEWL (Set 3; Day 35) | 7.09 | 9.00 | 1.27 | (1.05, 1.54) | (1.02, 1.58) | (0.99, 1.62) | (0.96, 1.68) | (0.90, 1.78) |
| N tapes required for barrier disruption: Day 28 | 38.58 | 55.55 | 1.44 | (1.16, 1.79) | (1.13, 1.84) | (1.09, 1.90) | (1.05, 1.98) | (0.98, 2.11) |
| *Estimated in imputed data | | | | | | | | |
