## Supplementary material for "A randomised controlled pilot trial of oral 11β-HSD1 inhibitor AZD4017 for wound healing in adults with type 2 diabetes mellitus": Table S6

#### Table S6: Unadjusted differences in changes from baseline in primary and secondary efficacy outcomes

Population: Full analysis set

Multiple imputation was used to address missing data. Due to issues with the data distributions, for all variables except TEWL, integrity, wound diameter & depth, median regression was used to estimate confidence intervals around differences between the groups. For TEWL, integrity, wound depth and diameter, linear regression was used. TEWL and integrity measurements were log-transformed prior to analysis. For these variables changes from baseline have been expressed as ratios of follow-up measurements to baseline values; between-group differences have been expressed as ratios of geometric means (AZD:PCB).

| **Variable** | **Median*** | | **Difference** | **Confidence interval** | | | | |
| --- | --- | --- | --- | --- | --- | --- | --- | --- |
|  | PCB n=14 | AZD n=14 | AZD-PCB | 75% | 80% | 85% | 90% | 95% |
| 11bHSD1 activity radioassay (% conv/24hrs): Day 28 | -3.10 | -0.50 | 2.60 | (-1.63, 6.83) | (-2.13, 7.33) | (-2.74, 7.94) | (-3.54, 8.74) | (-4.81, 10.01) |
| 11bHSD1 activity ELISA (% conv/24hrs): Day 28 | -3.11 | -1.80 | 1.31 | (-2.68, 5.29) | (-3.15, 5.76) | (-3.73, 6.34) | (-4.48, 7.09) | (-5.68, 8.29) |
| Sudomotor function Left Hand (micro S): Day 35 | -0.30 | 1.70 | 2.00 | (-7.20, 11.20) | (-8.28, 12.28) | (-9.60, 13.60) | (-11.35, 15.35) | (-14.11, 18.11) |
| Sudomotor function Right Hand (micro S): Day 35 | 3.65 | 5.05 | 1.40 | (-6.70, 9.50) | (-7.66, 10.46) | (-8.83, 11.63) | (-10.37, 13.17) | (-12.81, 15.61) |
| Sudomotor function Hands (micro S): Day 35 | 0.45 | 4.03 | 3.58 | (-4.65, 11.80) | (-5.62, 12.77) | (-6.81, 13.96) | (-8.38, 15.53) | (-10.86, 18.01) |
| Sudomotor function Left Foot (micro S): Day 35 | 5.75 | -4.00 | -9.75 | (-17.14, -2.36) | (-18.01, -1.49) | (-19.07, -0.43) | (-20.48, 0.98) | (-22.70, 3.20) |
| Sudomotor function Right Foot (micro S): Day 35 | 6.80 | -1.80 | -8.60 | (-14.55, -2.65) | (-15.26, -1.94) | (-16.11, -1.09) | (-17.24, 0.04) | (-19.03, 1.83) |
| Sudomotor function Feet (micro S): Day 35 | 5.90 | -3.45 | -9.35 | (-15.79, -2.91) | (-16.55, -2.15) | (-17.48, -1.22) | (-18.70, 0.00) | (-20.63, 1.93) |
| Sudomotor function Overall (micro S): Day 35 | 4.71 | 0.93 | -3.79 | (-8.68, 1.10) | (-9.26, 1.68) | (-9.97, 2.40) | (-10.92, 3.34) | (-12.42, 4.84) |
| Skin hydration (A.U): Day 35 | -2.88 | 3.06 | 5.94 | (0.67, 11.21) | (0.05, 11.83) | (-0.71, 12.59) | (-1.71, 13.59) | (-3.29, 15.17) |
| Epidermal thickness (micro m): Day 35 | -0.43 | -1.92 | -1.49 | (-11.12, 8.15) | (-12.25, 9.28) | (-13.64, 10.66) | (-15.46, 12.49) | (-18.35, 15.38) |
| Cortisol (mcg/24h): Day 35 | -2.70 | -1.00 | 1.70 | (-31.61, 35.01) | (-35.54, 38.94) | (-40.33, 43.73) | (-46.66, 50.06) | (-56.65, 60.05) |
| Urinary [THF+alloTHF]/THE ratio: Day 35 | 0.01 | -0.84 | -0.85 | (-0.94, -0.75) | (-0.95, -0.74) | (-0.96, -0.73) | (-0.98, -0.71) | (-1.01, -0.68) |
| **Variable** | **Ratio FU:BL*** | | **Ratio** | **Confidence interval** | | | | |
|  | PCB n=14 | AZD n=14 | AZD:PCB | 75% | 80% | 85% | 90% | 95% |
| Hour 3 TEWL (Set 1; Day 0) | 4.24 | 3.49 | 0.82 | (0.69, 0.98) | (0.68, 1.00) | (0.66, 1.02) | (0.64, 1.06) | (0.61, 1.12) |
| Hour 48 TEWL (Set 1; Day 2) | 2.40 | 2.33 | 0.97 | (0.80, 1.18) | (0.78, 1.21) | (0.75, 1.25) | (0.73, 1.29) | (0.68, 1.37) |
| Hour 168 TEWL (Set 1; Day 7) | 1.64 | 1.78 | 1.09 | (0.85, 1.39) | (0.82, 1.43) | (0.80, 1.48) | (0.76, 1.56) | (0.70, 1.68) |
| Hour 3 TEWL (Set 2; Day 28) | 2.85 | 3.25 | 1.14 | (0.89, 1.45) | (0.87, 1.49) | (0.84, 1.55) | (0.80, 1.62) | (0.74, 1.74) |
| Hour 48 TEWL (Set 2; Day 30) | 1.78 | 1.98 | 1.12 | (0.89, 1.40) | (0.87, 1.44) | (0.84, 1.48) | (0.80, 1.55) | (0.75, 1.66) |
| Hour 168 TEWL (Set 2; Day 35) | 1.21 | 1.19 | 0.98 | (0.78, 1.23) | (0.76, 1.26) | (0.73, 1.31) | (0.70, 1.37) | (0.66, 1.46) |
| Hour 0 TEWL (Set 3; Day 35) | 0.86 | 0.98 | 1.14 | (0.86, 1.51) | (0.83, 1.56) | (0.80, 1.62) | (0.76, 1.71) | (0.70, 1.86) |
| N tapes required for barrier disruption: Day 28 | 0.87 | 1.09 | 1.26 | (1.01, 1.58) | (0.98, 1.63) | (0.95, 1.68) | (0.91, 1.75) | (0.85, 1.88) |
| *Estimated in imputed data. BL=baseline; FU=follow-up | | | | | | | | |
