## Supplementary material for "A randomised controlled pilot trial of oral 11β-HSD1 inhibitor AZD4017 for wound healing in adults with type 2 diabetes mellitus": Table S7

#### Table S7: Adjusted differences in changes from baseline in primary and secondary efficacy outcomes

Population: Full analysis set

Multiple imputation was used to address missing data. Due to issues with the data distributions, for all variables except TEWL, integrity, wound diameter & depth, median regression was used to estimate confidence intervals around differences between the groups. For TEWL & integrity, linear regression was used. TEWL and integrity measurements were log-transformed prior to analysis. For these variables changes from baseline have been expressed as ratios of follow-up measurements to baseline values; between-group differences have been expressed as ratios of geometric means (AZD:PCB). For each variable, the comparison was adjusted for baseline value of the variable (not applicable to wound diameter & depth), age, sex and baseline HbA1c.

| **Variable** | **Median*** | | **Difference** | **Confidence interval** | | | | |
| --- | --- | --- | --- | --- | --- | --- | --- | --- |
|  | PCB n=14 | AZD n=14 | AZD-PCB | 75% | 80% | 85% | 90% | 95% |
| 11bHSD1 activity radioassay (% conv/24hrs): Day 28 | -2.99 | -1.94 | 1.05 | (-1.98, 4.09) | (-2.34, 4.45) | (-2.78, 4.89) | (-3.37, 5.47) | (-4.30, 6.41) |
| 11bHSD1 activity ELISA (% conv/24hrs): Day 28 | -3.93 | -4.47 | -0.54 | (-2.78, 1.70) | (-3.05, 1.97) | (-3.38, 2.30) | (-3.82, 2.74) | (-4.53, 3.45) |
| Sudomotor function Left Hand (micro S): Day 35 | -0.01 | 10.38 | 10.39 | (2.03, 18.74) | (1.04, 19.73) | (-0.18, 20.95) | (-1.80, 22.57) | (-4.38, 25.15) |
| Sudomotor function Right Hand (micro S): Day 35 | 2.77 | 7.77 | 5.01 | (-3.07, 13.08) | (-4.03, 14.04) | (-5.20, 15.21) | (-6.75, 16.76) | (-9.21, 19.23) |
| Sudomotor function Hands (micro S): Day 35 | 1.71 | 8.96 | 7.25 | (-1.05, 15.56) | (-2.04, 16.54) | (-3.24, 17.75) | (-4.84, 19.35) | (-7.39, 21.89) |
| Sudomotor function Left Foot (micro S): Day 35 | 6.91 | -0.10 | -7.01 | (-14.81, 0.79) | (-15.73, 1.71) | (-16.86, 2.84) | (-18.36, 4.34) | (-20.73, 6.71) |
| Sudomotor function Right Foot (micro S): Day 35 | 5.49 | 1.43 | -4.07 | (-10.35, 2.21) | (-11.09, 2.96) | (-12.00, 3.87) | (-13.21, 5.07) | (-15.12, 6.99) |
| Sudomotor function Feet (micro S): Day 35 | 5.77 | 1.24 | -4.52 | (-11.36, 2.32) | (-12.18, 3.13) | (-13.17, 4.12) | (-14.48, 5.43) | (-16.56, 7.51) |
| Sudomotor function Overall (micro S): Day 35 | 3.99 | 2.66 | -1.33 | (-6.76, 4.10) | (-7.41, 4.75) | (-8.20, 5.54) | (-9.25, 6.59) | (-10.92, 8.25) |
| Skin hydration (A.U): Day 35 | -2.85 | 2.82 | 5.67 | (-0.08, 11.43) | (-0.77, 12.12) | (-1.62, 12.97) | (-2.75, 14.10) | (-4.57, 15.92) |
| Epidermal thickness (micro m): Day 35 | -2.38 | 3.20 | 5.58 | (-1.54, 12.69) | (-2.40, 13.55) | (-3.45, 14.60) | (-4.85, 16.01) | (-7.11, 18.26) |
| Cortisol (mcg/24h): Day 35 | -14.42 | -9.12 | 5.29 | (-19.05, 29.64) | (-21.94, 32.53) | (-25.47, 36.06) | (-30.15, 40.74) | (-37.58, 48.16) |
| Urinary [THF+alloTHF]/THE ratio: Day 35 | 0.01 | -0.85 | -0.87 | (-0.98, -0.75) | (-1.00, -0.73) | (-1.01, -0.72) | (-1.04, -0.69) | (-1.07, -0.66) |
| **Variable** | **Ratio FU:BL*** | | **Ratio** | **Confidence interval** | | | | |
|  | PCB n=14 | AZD n=14 | AZD:PCB | 75% | 80% | 85% | 90% | 95% |
| Hour 3 TEWL (Set 1; Day 0) | 4.03 | 3.68 | 0.91 | (0.81, 1.03) | (0.79, 1.05) | (0.78, 1.07) | (0.76, 1.09) | (0.73, 1.13) |
| Hour 48 TEWL (Set 1; Day 2) | 2.35 | 2.37 | 1.01 | (0.86, 1.19) | (0.84, 1.21) | (0.82, 1.24) | (0.79, 1.28) | (0.76, 1.34) |
| Hour 168 TEWL (Set 1; Day 7) | 1.62 | 1.80 | 1.11 | (0.86, 1.44) | (0.83, 1.49) | (0.80, 1.54) | (0.76, 1.62) | (0.70, 1.76) |
| Hour 3 TEWL (Set 2; Day 28) | 2.65 | 3.53 | 1.33 | (1.09, 1.63) | (1.06, 1.67) | (1.03, 1.72) | (0.99, 1.79) | (0.93, 1.90) |
| Hour 48 TEWL (Set 2; Day 30) | 1.68 | 2.11 | 1.26 | (1.03, 1.53) | (1.01, 1.56) | (0.98, 1.61) | (0.94, 1.67) | (0.89, 1.77) |
| Hour 168 TEWL (Set 2; Day 35) | 1.16 | 1.24 | 1.07 | (0.87, 1.31) | (0.85, 1.34) | (0.82, 1.38) | (0.79, 1.44) | (0.74, 1.53) |
| Hour 0 TEWL (Set 3; Day 35) | 0.80 | 1.06 | 1.33 | (1.07, 1.65) | (1.04, 1.69) | (1.01, 1.75) | (0.97, 1.82) | (0.91, 1.95) |
| N tapes required for barrier disruption: Day 28 | 0.82 | 1.17 | 1.43 | (1.18, 1.74) | (1.15, 1.78) | (1.12, 1.84) | (1.07, 1.91) | (1.01, 2.02) |
| *Estimated in imputed data, adjusted for baseline value, age, sex and baseline HbA1c. BL=baseline; FU=follow-up | | | | | | | | |
