## Supplementary material for "A randomised controlled pilot trial of oral 11β-HSD1 inhibitor AZD4017 for wound healing in adults with type 2 diabetes mellitus": Table S8

### Table S8: Sensitivity analysis of trans-epidermal water loss (TEWL) adjusting for exact timing of post-disruption measurements

Population: Full analysis set

Analysis models included exact measurement times to account for variation in timings; results have been provided for a) data imputed including all observed values and b) data re-imputed after deleting TEWL readings that were potentially unreliable due to high temperatures.

Linear mixed models were used to model post-baseline TEWL readings (within each set) as a function of time since disruption. Quadratic functions were added to allow for nonlinear change over time. Baseline values and changes over time were permitted to vary between patients. TEWL measurements were log-transformed prior to analysis; differences have been expressed as ratios of geometric means (AZD:PCB). For each variable, the comparison was adjusted for baseline (day 0, hour 0) TEWL, age, sex and baseline HbA1c.

| **Dataset** | **Disruption day** | **Post-disruption hour** | **Ratio AZD:PCB** | **Confidence interval** | | | | |
| --- | --- | --- | --- | --- | --- | --- | --- | --- |
|  |  |  |  | 75% | 80% | 85% | 90% | 95% |
| Imputed | 0 | 3 | 0.89 | (0.78 , 1.00) | (0.77 , 1.02) | (0.76 , 1.03) | (0.74 , 1.06) | (0.72 , 1.09) |
| Imputed | 0 | 48 | 1.00 | (0.89 , 1.14) | (0.88 , 1.15) | (0.86 , 1.17) | (0.84 , 1.20) | (0.81 , 1.24) |
| Imputed | 0 | 168 | 1.17 | (0.94 , 1.45) | (0.91 , 1.49) | (0.89 , 1.53) | (0.85 , 1.59) | (0.80 , 1.69) |
| Imputed | 28 | 3 | 1.28 | (1.08 , 1.52) | (1.06 , 1.55) | (1.04 , 1.59) | (1.01 , 1.64) | (0.96 , 1.72) |
| Imputed | 28 | 48 | 1.25 | (1.07 , 1.47) | (1.05 , 1.50) | (1.03 , 1.53) | (1.00 , 1.57) | (0.96 , 1.64) |
| Imputed | 28 | 168 | 1.12 | (0.93 , 1.34) | (0.92 , 1.36) | (0.89 , 1.40) | (0.86 , 1.44) | (0.82 , 1.52) |
| Re-imputed | 0 | 3 | 0.88 | (0.78 , 1.00) | (0.77 , 1.01) | (0.76 , 1.03) | (0.74 , 1.05) | (0.72 , 1.08) |
| Re-imputed | 0 | 48 | 0.90 | (0.80 , 1.01) | (0.79 , 1.03) | (0.78 , 1.04) | (0.76 , 1.07) | (0.74 , 1.10) |
| Re-imputed | 0 | 168 | 0.90 | (0.77 , 1.04) | (0.76 , 1.06) | (0.74 , 1.08) | (0.72 , 1.11) | (0.69 , 1.16) |
| Re-imputed | 28 | 3 | 1.28 | (1.08 , 1.51) | (1.06 , 1.54) | (1.03 , 1.58) | (1.00 , 1.63) | (0.95 , 1.71) |
| Re-imputed | 28 | 48 | 1.25 | (1.07 , 1.47) | (1.05 , 1.50) | (1.02 , 1.53) | (1.00 , 1.58) | (0.95 , 1.65) |
| Re-imputed | 28 | 168 | 1.11 | (0.93 , 1.33) | (0.91 , 1.36) | (0.89 , 1.39) | (0.86 , 1.44) | (0.82 , 1.51) |
