## Supplementary material for "A randomised controlled pilot trial of oral 11β-HSD1 inhibitor AZD4017 for wound healing in adults with type 2 diabetes mellitus": Table S10

### Table S10: Sensitivity analysis of primary and secondary efficacy outcomes using last observation carried forward

Population: Full analysis set

Due to issues with the data distributions, for all variables except TEWL, integrity, wound diameter & depth, quantile (median) regression was used to obtain point estimates and confidence intervals for differences between groups. For TEWL, integrity, wound depth and diameter, linear regression was used. TEWL and integrity measurements were log-transformed prior to analysis; differences have been expressed as ratios of geometric means (AZD:PBO). For each variable, the comparison was adjusted for baseline value of the variable (where applicable; not applicable to wound diameter & depth), age, sex and baseline HbA1c. All TEWL readings were adjusted using TEWL baseline at day 0.Note that day 0 TEWL measurements were missed in error for 1 patient therefore data could only be carried forward for 27 patients.

| **Variable** | **Estimated median*** | | **Difference*** | **Confidence interval*** | | | | |
| --- | --- | --- | --- | --- | --- | --- | --- | --- |
|  | PCB | AZD | AZD-PCB | 75% | 80% | 85% | 90% | 95% |
| 11bHSD1 activity radioassay (% conv/24hrs): Day 28 | 13.41 (n=14) | 12.69 (n=14) | -0.72 | (-3.71, 2.27) | (-3.83, 2.39) | (-4.59, 3.15) | (-5.13, 3.68) | (-5.73, 4.29) |
| 11bHSD1 activity ELISA (% conv/24hrs): Day 28 | 5.76 (n=14) | 4.36 (n=14) | -1.39 | (-3.65, 0.87) | (-3.76, 0.97) | (-3.86, 1.07) | (-4.39, 1.60) | (-4.79, 2.00) |
| Sudomotor function Left Hand (micro S): Day 35 | 56.92 (n=14) | 59.57 (n=14) | 2.65 | (-5.65, 10.96) | (-5.99, 11.29) | (-6.38, 11.68) | (-8.80, 14.10) | (-10.68, 15.98) |
| Sudomotor function Right Hand (micro S): Day 35 | 54.60 (n=14) | 59.45 (n=14) | 4.85 | (-1.92, 11.62) | (-3.00, 12.71) | (-3.35, 13.06) | (-6.44, 16.14) | (-8.12, 17.83) |
| Sudomotor function Hands (micro S): Day 35 | 55.06 (n=14) | 57.94 (n=14) | 2.88 | (-3.84, 9.60) | (-4.12, 9.89) | (-4.64, 10.40) | (-8.55, 14.31) | (-9.79, 15.55) |
| Sudomotor function Left Foot (micro S): Day 35 | 75.20 (n=14) | 69.34 (n=14) | -5.86 | (-10.67, -1.06) | (-10.87, -0.86) | (-11.22, -0.51) | (-11.69, -0.04) | (-19.25, 7.53) |
| Sudomotor function Right Foot (micro S): Day 35 | 73.12 (n=14) | 69.90 (n=14) | -3.23 | (-6.96, 0.51) | (-7.41, 0.95) | (-8.42, 1.97) | (-10.27, 3.82) | (-13.35, 6.89) |
| Sudomotor function Feet (micro S): Day 35 | 74.01 (n=14) | 69.70 (n=14) | -4.31 | (-8.44, -0.18) | (-8.61, -0.01) | (-9.63, 1.01) | (-11.71, 3.10) | (-15.60, 6.98) |
| Sudomotor function Overall (micro S): Day 35 | 64.24 (n=14) | 63.79 (n=14) | -0.45 | (-4.23, 3.32) | (-5.09, 4.19) | (-7.76, 6.86) | (-8.73, 7.83) | (-9.40, 8.50) |
| Skin hydration (A.U): Day 35 | 38.34 (n=14) | 43.42 (n=14) | 5.08 | (1.95, 8.21) | (0.66, 9.50) | (0.46, 9.70) | (-1.79, 11.94) | (-2.86, 13.02) |
| Epidermal thickness (micro m): Day 35 | 63.31 (n=14) | 65.43 (n=14) | 2.11 | (-4.24, 8.46) | (-4.64, 8.87) | (-5.63, 9.86) | (-6.09, 10.32) | (-6.88, 11.11) |
| Cortisol (mcg/24h): Day 35 | 64.34 (n=14) | 68.42 (n=14) | 4.08 | (-18.81, 26.97) | (-23.30, 31.47) | (-26.68, 34.84) | (-28.32, 36.48) | (-37.79, 45.95) |
| Urinary [THF+alloTHF]/THE ratio: Day 35 | 0.99 (n=14) | 0.17 (n=14) | -0.82 | (-0.96, -0.68) | (-0.96, -0.68) | (-1.01, -0.63) | (-1.02, -0.62) | (-1.04, -0.60) |
|  | **Estimated mean*** | | **Difference*** | **Confidence interval*** | | | | |
|  | PCB | AZD | AZD-PCB | 75% | 80% | 85% | 90% | 95% |
| Wound gap diameter (mm): Day 2 | 1.51 (n=14) | 0.98 (n=14) | -0.52 | (-0.82, -0.23) | (-0.85, -0.20) | (-0.89, -0.15) | (-0.95, -0.10) | (-1.04, -0.01) |
| Wound depth (mm): Day 7 | 0.60 (n=14) | 0.59 (n=14) | -0.01 | (-0.10, 0.09) | (-0.11, 0.10) | (-0.13, 0.11) | (-0.14, 0.13) | (-0.17, 0.16) |
| Wound gap diameter (mm): Day 30 | 1.32 (n=14) | 0.66 (n=14) | -0.66 | (-0.94, -0.37) | (-0.98, -0.34) | (-1.02, -0.30) | (-1.07, -0.24) | (-1.16, -0.16) |
| Wound depth (mm): Day 35 | 0.59 (n=14) | 0.56 (n=14) | -0.03 | (-0.11, 0.05) | (-0.12, 0.06) | (-0.13, 0.07) | (-0.15, 0.09) | (-0.17, 0.11) |
|  | **Estimated geometric mean*** | | **Ratio*** | **Confidence interval*** | | | | |
|  | PCB | AZD | AZD:PCB | 75% | 80% | 85% | 90% | 95% |
| Hour 3 TEWL (Set 1; Day 0) | 34.85 (n=14) | 29.97 (n=13) | 0.86 | (0.75, 0.98) | (0.74, 1.00) | (0.73, 1.02) | (0.71, 1.04) | (0.68, 1.08) |
| Hour 48 TEWL (Set 1; Day 2) | 20.37 (n=14) | 21.11 (n=13) | 1.04 | (0.89, 1.21) | (0.87, 1.23) | (0.85, 1.26) | (0.83, 1.30) | (0.79, 1.36) |
| Hour 168 TEWL (Set 1; Day 7) | 14.59 (n=14) | 15.22 (n=13) | 1.04 | (0.81, 1.35) | (0.78, 1.39) | (0.75, 1.44) | (0.72, 1.52) | (0.66, 1.64) |
| Hour 3 TEWL (Set 2; Day 28) | 21.70 (n=14) | 29.62 (n=13) | 1.36 | (1.08, 1.73) | (1.05, 1.78) | (1.01, 1.84) | (0.97, 1.93) | (0.90, 2.07) |
| Hour 48 TEWL (Set 2; Day 30) | 13.87 (n=14) | 18.00 (n=13) | 1.30 | (1.05, 1.60) | (1.03, 1.64) | (1.00, 1.69) | (0.96, 1.76) | (0.90, 1.88) |
| Hour 168 TEWL (Set 2; Day 35) | 9.92 (n=14) | 11.87 (n=13) | 1.20 | (0.95, 1.51) | (0.92, 1.56) | (0.89, 1.61) | (0.85, 1.68) | (0.79, 1.81) |
| Hour 0 TEWL (Set 3; Day 35) | 6.88 (n=14) | 9.67 (n=13) | 1.41 | (1.12, 1.77) | (1.09, 1.81) | (1.05, 1.88) | (1.01, 1.96) | (0.94, 2.10) |
| N tapes required for barrier disruption: Day 28 | 39.36 (n=14) | 55.58 (n=13) | 1.41 | (1.15, 1.73) | (1.13, 1.77) | (1.09, 1.82) | (1.05, 1.90) | (0.99, 2.02) |
| *Adjusted for baseline value if applicable, age, sex and baseline HbA1c. | | | | | | | | |
