## Supplementary material for "A randomised controlled pilot trial of oral 11β-HSD1 inhibitor AZD4017 for wound healing in adults with type 2 diabetes mellitus": Table S11

### Table S11: Correlations between AZD4017 compliance and efficacy outcomes

Population: Full analysis set (AZD group only)

Absolute rho values >/= 0.3 were considered preliminary evidence of association. Sensitivity analysis excluded baseline radioimmunoassay samples that failed QC, an outlying ELISA assay result at day 28, TEWL readings recorded on very hot days, and results that were recorded 1 day earlier than planned, prior to imputation. Note that not all of these issues occurred in samples from the AZD group but could have altered imputed values for any incomplete variables.

|  | **All values** | | **Excl. outliers/QC failures** | |
| --- | --- | --- | --- | --- |
| **Variable** | **Imputed** | **Available case** | **Imputed** | **Available case** |
| 11bHSD1 activity radioassay (% conv/24hrs): Day 28 | -0.04 (14) | -0.04 (14) | -0.04 (14) | -0.04 (14) |
| 11bHSD1 activity ELISA (% conv/24hrs): Day 28 | -0.74 (14) | -0.74 (14) | -0.74 (14) | -0.74 (14) |
| Sudomotor function Left Hand (micro S): Day 35 | -0.05 (14) | -0.08 (13) | -0.03 (14) | -0.08 (13) |
| Sudomotor function Right Hand (micro S): Day 35 | -0.16 (14) | -0.23 (13) | -0.16 (14) | -0.23 (13) |
| Sudomotor function Hands (micro S): Day 35 | -0.03 (14) | -0.07 (13) | -0.03 (14) | -0.07 (13) |
| Sudomotor function Left Foot (micro S): Day 35 | -0.21 (14) | -0.20 (13) | -0.19 (14) | -0.20 (13) |
| Sudomotor function Right Foot (micro S): Day 35 | 0.06 (14) | 0.12 (13) | 0.09 (14) | 0.12 (13) |
| Sudomotor function Feet (micro S): Day 35 | -0.18 (14) | -0.17 (13) | -0.15 (14) | -0.17 (13) |
| Sudomotor function Overall (micro S): Day 35 | 0.03 (14) | 0.05 (13) | 0.05 (14) | 0.05 (13) |
| Skin hydration (A.U): Day 35 | 0.55 (14) | 0.77 (12) | 0.62 (14) | 0.77 (12) |
| Epidermal thickness (micro m): Day 35 | -0.11 (14) | -0.15 (12) | -0.11 (14) | -0.15 (12) |
| Cortisol (mcg/24h): Day 35 | -0.30 (14) | -0.31 (13) | -0.31 (14) | -0.31 (13) |
| Urinary [THF+alloTHF]/THE ratio: Day 35 | -0.28 (14) | -0.34 (13) | -0.26 (14) | -0.34 (13) |
| Wound gap diameter (mm): Day 2 | 0.35 (14) | 0.35 (14) | 0.35 (14) | 0.35 (14) |
| Wound depth (mm): Day 7 | 0.23 (14) | 0.23 (14) | 0.27 (14) | 0.31 (13) |
| Wound gap diameter (mm): Day 30 | -0.06 (14) | -0.01 (12) | -0.07 (14) | -0.01 (12) |
| Wound depth (mm): Day 35 | 0.09 (14) | 0.10 (12) | 0.09 (14) | 0.10 (12) |
| Hour 0 TEWL (Set 1; Day 0) | -0.42 (14) | -0.43 (13) | -0.43 (14) | -0.43 (13) |
| Hour 3 TEWL (Set 1; Day 0) | -0.35 (14) | -0.39 (12) | -0.34 (14) | -0.39 (12) |
| Hour 48 TEWL (Set 1; Day 2) | -0.20 (14) | -0.13 (11) | -0.25 (14) | -0.17 (8) |
| Hour 168 TEWL (Set 1; Day 7) | -0.07 (14) | -0.01 (13) | 0.18 (14) | 0.38 (9) |
| Hour 0 TEWL (Set 2; Day 28) | -0.33 (14) | -0.33 (14) | -0.33 (14) | -0.33 (14) |
| Hour 3 TEWL (Set 2; Day 28) | -0.37 (14) | -0.37 (14) | -0.37 (14) | -0.37 (14) |
| Hour 48 TEWL (Set 2; Day 30) | 0.29 (14) | 0.29 (14) | 0.29 (14) | 0.29 (14) |
| Hour 168 TEWL (Set 2; Day 35) | 0.37 (14) | 0.38 (12) | 0.34 (14) | 0.38 (12) |
| Hour 0 TEWL (Set 3; Day 35) | 0.30 (14) | 0.39 (12) | 0.31 (14) | 0.39 (12) |
| N tapes required for barrier disruption: Day 28 | -0.15 (14) | -0.15 (14) | -0.15 (14) | -0.15 (14) |
| All values presented as Spearman's rho (number of patients included) | | | | |
