## Supplementary material for "A randomised controlled pilot trial of oral 11β-HSD1 inhibitor AZD4017 for wound healing in adults with type 2 diabetes mellitus": Table S12

#### Table S12: Unadjusted differences in final values of longitudinal laboratory safety variables

Population: Safety set

Multiple imputation was used to address missing data. All point estimates and confidence intervals estimated via linear regression.

| **Variable** | **Mean PCB (N=14), AZD (N=14); Difference AZD-PCB (90% CI)** | | | |
| --- | --- | --- | --- | --- |
|  | Day 7 | Day 28 | Day 35 | Day 42 |
| Body Mass Index (kg / m2) | N/A | N/A | 33.90, 35.55; 1.65 (-5.35, 8.65) | N/A |
| Waist-hip ratio | N/A | N/A | 0.98, 1.02; 0.04 (-0.01, 0.09) | N/A |
| Systolic blood pressure (mm Hg) | N/A | N/A | 143.66, 129.01; -14.64 (-22.83, -6.45) | 136.56, 137.50; 0.94 (-7.02, 8.89) |
| Diastolic blood pressure (mm Hg) | N/A | N/A | 79.95, 73.64; -6.31 (-11.24, -1.39) | 80.11, 79.29; -0.83 (-8.02, 6.37) |
| HbA1c (mmol/mol) | 71.88, 65.81; -6.07 (-17.14, 5.00) | 70.14, 66.00; -4.14 (-14.73, 6.45) | 70.64, 64.79; -5.85 (-17.14, 5.44) | 69.37, 65.43; -3.94 (-14.93, 7.05) |
| High density lipoprotein (mmol/l) | 1.16, 1.05; -0.11 (-0.28, 0.07) | 1.22, 1.11; -0.11 (-0.28, 0.07) | 1.18, 1.01; -0.17 (-0.34, 0.00) | 1.19, 1.21; 0.02 (-0.18, 0.22) |
| Cholesterol (mmol/l) | 4.31, 3.59; -0.71 (-1.26, -0.17) | 4.21, 3.54; -0.67 (-1.17, -0.16) | 4.11, 3.44; -0.68 (-1.18, -0.17) | 4.06, 3.84; -0.22 (-0.77, 0.33) |
| Triglycerides (mmol/l) | 2.11, 2.39; 0.28 (-0.51, 1.07) | 1.81, 1.71; -0.11 (-0.63, 0.42) | 2.22, 2.37; 0.15 (-0.91, 1.21) | 1.61, 1.94; 0.33 (-0.39, 1.06) |
| Haemoglobin (g/l) | 136.07, 137.43; 1.36 (-6.38, 9.09) | 138.46, 138.36; -0.10 (-6.97, 6.77) | 134.84, 136.30; 1.46 (-6.54, 9.46) | 136.56, 139.86; 3.29 (-5.51, 12.09) |
| White cells (x109/l) | 6.71, 7.96; 1.26 (-0.02, 2.54) | 6.52, 6.72; 0.20 (-0.97, 1.38) | 6.68, 6.93; 0.25 (-1.00, 1.50) | 6.55, 7.66; 1.12 (-0.23, 2.46) |
| Platelets (x109/l) | 219.36, 275.29; 55.93 (8.28, 103.58) | 219.37, 288.57; 69.20 (15.99, 122.41) | 213.32, 265.82; 52.50 (2.98, 102.03) | 220.63, 281.43; 60.80 (10.77, 110.82) |
| Red cells (x1012/l) | 4.74, 4.59; -0.16 (-0.48, 0.16) | 4.81, 4.68; -0.13 (-0.41, 0.16) | 4.67, 4.57; -0.10 (-0.39, 0.19) | 4.73, 4.77; 0.04 (-0.29, 0.38) |
| Mean corpuscular volume (fl) | 89.71, 92.14; 2.43 (-1.26, 6.12) | 88.30, 91.21; 2.92 (-1.05, 6.88) | 90.23, 92.20; 1.96 (-2.13, 6.06) | 88.19, 90.50; 2.31 (-1.16, 5.78) |
| Haematocrit (packed cell volume) | 0.43, 0.42; -0.01 (-0.03, 0.02) | 0.42, 0.43; 0.00 (-0.02, 0.03) | 0.42, 0.42; 0.00 (-0.03, 0.03) | 0.42, 0.43; 0.02 (-0.01, 0.04) |
| Mean corpuscular haemoglobin (pg) | 28.79, 30.13; 1.34 (-0.12, 2.80) | 28.96, 29.68; 0.72 (-0.61, 2.05) | 28.97, 29.99; 1.02 (-0.44, 2.48) | 29.01, 29.42; 0.41 (-1.02, 1.85) |
| Corpuscular hemoglobin concentration (g/l) | 321.07, 326.93; 5.86 (-0.96, 12.68) | 328.44, 325.79; -2.66 (-9.90, 4.59) | 321.93, 325.20; 3.27 (-3.47, 10.01) | 328.81, 324.86; -3.95 (-11.54, 3.64) |
| Red blood cell distribution width (%) | 14.43, 14.27; -0.16 (-0.94, 0.62) | 13.98, 14.08; 0.10 (-0.45, 0.65) | 14.21, 14.42; 0.21 (-0.38, 0.81) | 13.76, 13.74; -0.02 (-0.68, 0.64) |
| Albumin (g/l) | 39.50, 40.14; 0.64 (-1.23, 2.52) | 38.94, 38.71; -0.22 (-1.95, 1.51) | 39.88, 40.04; 0.16 (-1.65, 1.98) | 38.29, 38.84; 0.56 (-0.80, 1.91) |
| Blirubin (umol/l) | 8.36, 8.64; 0.29 (-2.43, 3.00) | 9.81, 9.86; 0.05 (-2.50, 2.60) | 8.80, 8.36; -0.44 (-3.41, 2.53) | 8.29, 8.40; 0.11 (-1.54, 1.75) |
| Alkaline phosphatase (U/l) | 80.86, 80.14; -0.71 (-17.98, 16.56) | 78.01, 68.93; -9.08 (-23.40, 5.24) | 77.67, 68.18; -9.50 (-23.41, 4.41) | 78.30, 76.94; -1.36 (-19.31, 16.60) |
| Alanine aminotransferase (iu/l) | 22.93, 26.14; 3.21 (-2.22, 8.64) | 23.19, 23.07; -0.12 (-5.46, 5.22) | 22.81, 22.19; -0.61 (-6.76, 5.53) | 21.74, 23.96; 2.23 (-3.55, 8.00) |
| Aspartate aminotransferase (iu/l) | 21.07, 22.36; 1.29 (-1.98, 4.55) | 20.63, 21.36; 0.73 (-1.56, 3.01) | 20.76, 20.92; 0.16 (-2.17, 2.49) | 19.15, 21.80; 2.65 (-0.73, 6.03) |
| Gamma-glutamyl transpeptidase (iu/l) | 39.71, 37.71; -2.00 (-22.51, 18.51) | 42.23, 29.86; -12.37 (-29.42, 4.68) | 43.29, 28.10; -15.19 (-35.42, 5.05) | 40.96, 28.82; -12.14 (-29.49, 5.21) |
| eGFR (ml/min/1.73m2) | 77.50, 71.50; -6.00 (-14.97, 2.97) | 80.49, 75.14; -5.34 (-13.52, 2.83) | 76.61, 73.60; -3.02 (-13.42, 7.39) | 78.28, 76.21; -2.06 (-10.63, 6.51) |
| Sodium (mmol/l) | 139.29, 140.00; 0.71 (-0.83, 2.26) | 138.12, 138.79; 0.66 (-0.62, 1.95) | 140.05, 139.28; -0.78 (-2.63, 1.08) | 138.86, 138.00; -0.86 (-2.24, 0.52) |
| Potassium (mmol/l) | 4.79, 4.59; -0.20 (-0.50, 0.10) | 4.50, 4.74; 0.24 (0.00, 0.49) | 4.47, 4.52; 0.05 (-0.18, 0.28) | 4.63, 4.63; -0.00 (-0.26, 0.26) |
| Urea (mmol/l) | 6.91, 7.24; 0.33 (-1.18, 1.84) | 6.71, 6.56; -0.15 (-2.04, 1.74) | 7.68, 7.49; -0.20 (-2.70, 2.31) | 7.04, 7.57; 0.53 (-1.33, 2.38) |
| Creatinine (umol/l) | 82.36, 88.57; 6.21 (-6.95, 19.38) | 77.83, 80.93; 3.10 (-8.58, 14.77) | 84.69, 84.94; 0.25 (-14.58, 15.08) | 81.18, 79.93; -1.25 (-12.61, 10.11) |
| Testosterone (nmol/l) | 10.34, 6.43; -3.91 (-8.01, 0.18) | 11.08, 6.46; -4.62 (-8.48, -0.75) | 10.26, 6.46; -3.80 (-7.92, 0.31) | 10.25, 7.66; -2.59 (-6.72, 1.53) |
| Dehydroepiandrosterone sulphate (umol/l) | 3.56, 4.91; 1.36 (-0.58, 3.30) | 3.25, 5.35; 2.10 (0.53, 3.66) | 3.51, 4.91; 1.39 (-0.31, 3.10) | 3.00, 3.55; 0.55 (-0.75, 1.85) |
| Free thyroxine (pmol/l) | 14.63, 15.75; 1.12 (-0.28, 2.52) | 14.64, 15.74; 1.10 (-0.05, 2.26) | 14.39, 15.48; 1.09 (-0.05, 2.23) | 14.00, 15.10; 1.10 (0.15, 2.06) |
| Thyroid stimulating hormone (mlU/l) | 1.68, 1.84; 0.16 (-0.27, 0.59) | 1.89, 1.71; -0.18 (-0.71, 0.35) | 1.77, 1.68; -0.09 (-0.54, 0.35) | 1.79, 1.76; -0.03 (-0.57, 0.51) |
