## Supplementary material for "A randomised controlled pilot trial of oral 11β-HSD1 inhibitor AZD4017 for wound healing in adults with type 2 diabetes mellitus": Table S13

#### Table S13: Unadjusted differences in changes from baseline in longitudinal laboratory safety variables; (unadjusted; imputed)

Population: Safety set

Multiple imputation was used to address missing data. All point estimates and confidence intervals estimated via linear regression.

| **Variable** | **Mean* PCB (N-=14), AZD (N=14); Difference* AZD-PCB (90% CI)** | | | |
| --- | --- | --- | --- | --- |
|  | Day 7 | Day 28 | Day 35 | Day 42 |
| Body Mass Index (kg / m2) | N/A | N/A | 0.23, 0.51; 0.28 (-0.64, 1.19) | N/A |
| Waist-hip ratio | N/A | N/A | -0.00, -0.00; -0.00 (-0.03, 0.03) | N/A |
| Systolic blood pressure (mm Hg) | N/A | N/A | 7.94, -11.41; -19.36 (-31.07, -7.64) | 0.85, -2.93; -3.78 (-13.95, 6.39) |
| Diastolic blood pressure (mm Hg) | N/A | N/A | -3.90, 1.00; 4.90 (-1.16, 10.96) | -3.75, 6.64; 10.39 (2.27, 18.51) |
| HbA1c (mmol/mol) | -0.40, -0.19; 0.22 (-1.26, 1.69) | -2.15, 0.00; 2.15 (-0.89, 5.18) | -1.64, -1.21; 0.44 (-3.99, 4.86) | -2.92, -0.57; 2.35 (-2.59, 7.28) |
| High density lipoprotein (mmol/l) | -0.08, -0.14; -0.06 (-0.15, 0.02) | -0.01, -0.08; -0.07 (-0.14, 0.01) | -0.06, -0.19; -0.13 (-0.21, -0.05) | -0.04, 0.02; 0.06 (-0.04, 0.17) |
| Cholesterol (mmol/l) | -0.05, -0.36; -0.31 (-0.51, -0.10) | -0.15, -0.41; -0.26 (-0.66, 0.14) | -0.24, -0.51; -0.27 (-0.70, 0.17) | -0.29, -0.11; 0.19 (-0.34, 0.71) |
| Triglycerides (mmol/l) | 0.44, 0.37; -0.06 (-0.54, 0.41) | 0.13, -0.31; -0.45 (-0.85, -0.05) | 0.54, 0.35; -0.19 (-0.89, 0.50) | -0.07, -0.08; -0.01 (-0.50, 0.48) |
| Haemoglobin (g/l) | -3.14, -2.00; 1.14 (-2.06, 4.34) | -0.76, -1.07; -0.31 (-4.32, 3.69) | -4.38, -3.13; 1.24 (-3.57, 6.06) | -2.65, 0.43; 3.08 (-0.70, 6.86) |
| White cells (x109/l) | 0.25, 0.63; 0.37 (-0.33, 1.08) | 0.06, -0.62; -0.68 (-1.27, -0.09) | 0.22, -0.41; -0.63 (-1.40, 0.13) | 0.09, 0.33; 0.23 (-0.58, 1.05) |
| Platelets (x109/l) | 0.64, 2.14; 1.50 (-12.40, 15.40) | 0.66, 15.43; 14.77 (-6.85, 36.40) | -5.40, -7.32; -1.93 (-23.02, 19.17) | 1.92, 8.29; 6.37 (-10.84, 23.58) |
| Red cells (x1012/l) | -0.10, -0.13; -0.03 (-0.13, 0.07) | -0.03, -0.03; 0.00 (-0.11, 0.11) | -0.17, -0.14; 0.03 (-0.11, 0.16) | -0.11, 0.06; 0.17 (0.06, 0.29) |
| Mean corpuscular volume (fl) | 1.29, 0.93; -0.36 (-2.12, 1.41) | -0.13, 0.00; 0.13 (-1.68, 1.94) | 1.80, 0.98; -0.82 (-2.99, 1.35) | -0.24, -0.71; -0.48 (-2.01, 1.06) |
| Haematocrit (packed cell volume) | -0.00, -0.01; -0.01 (-0.02, 0.01) | -0.01, -0.00; 0.01 (-0.01, 0.02) | -0.01, -0.01; 0.00 (-0.01, 0.02) | -0.01, 0.01; 0.02 (0.00, 0.03) |
| Mean corpuscular haemoglobin (pg) | -0.03, 0.39; 0.42 (0.07, 0.78) | 0.14, -0.06; -0.19 (-0.59, 0.20) | 0.15, 0.25; 0.10 (-0.38, 0.59) | 0.19, -0.31; -0.50 (-0.99, -0.01) |
| Corpuscular hemoglobin concentration (g/l) | -4.50, 0.29; 4.79 (-1.61, 11.19) | 2.87, -0.86; -3.73 (-10.81, 3.35) | -3.64, -1.44; 2.20 (-5.76, 10.16) | 3.24, -1.79; -5.03 (-11.66, 1.61) |
| Red blood cell distribution width (%) | 0.55, 0.15; -0.40 (-0.89, 0.09) | 0.10, -0.04; -0.14 (-0.62, 0.34) | 0.33, 0.30; -0.03 (-0.46, 0.40) | -0.12, -0.38; -0.26 (-0.73, 0.21) |
| Albumin (g/l) | 0.07, 0.64; 0.57 (-0.61, 1.75) | -0.49, -0.79; -0.29 (-1.70, 1.12) | 0.45, 0.54; 0.09 (-1.45, 1.63) | -1.14, -0.66; 0.49 (-0.62, 1.59) |
| Blirubin (umol/l) | -0.57, 0.50; 1.07 (-0.75, 2.90) | 0.88, 1.71; 0.83 (-0.69, 2.35) | -0.13, 0.21; 0.35 (-2.19, 2.88) | -0.64, 0.26; 0.89 (-0.85, 2.64) |
| Alkaline phosphatase (U/l) | 0.43, -4.93; -5.36 (-11.92, 1.20) | -2.42, -16.14; -13.72 (-22.00, -5.44) | -2.75, -16.89; -14.14 (-21.88, -6.39) | -2.13, -8.13; -6.00 (-16.68, 4.68) |
| Alanine aminotransferase (iu/l) | -1.71, -1.29; 0.43 (-2.53, 3.38) | -1.45, -4.36; -2.90 (-6.02, 0.21) | -1.84, -5.24; -3.40 (-8.09, 1.29) | -2.91, -3.47; -0.56 (-4.51, 3.39) |
| Aspartate aminotransferase (iu/l) | 0.07, -0.29; -0.36 (-3.22, 2.51) | -0.37, -1.29; -0.92 (-3.59, 1.75) | -0.24, -1.73; -1.48 (-4.90, 1.94) | -1.85, -0.85; 1.01 (-2.75, 4.76) |
| Gamma-glutamyl transpeptidase (iu/l) | -2.14, -1.57; 0.57 (-2.73, 3.87) | 0.37, -9.43; -9.80 (-17.09, -2.51) | 1.43, -11.18; -12.61 (-25.48, 0.26) | -0.90, -10.47; -9.57 (-17.50, -1.64) |
| eGFR (ml/min/1.73m2) | -4.36, -7.71; -3.36 (-7.87, 1.15) | -1.37, -4.07; -2.70 (-5.83, 0.43) | -5.24, -5.62; -0.38 (-5.36, 4.61) | -3.58, -3.00; 0.58 (-3.91, 5.08) |
| Sodium (mmol/l) | 0.36, -0.71; -1.07 (-3.83, 1.69) | -0.81, -1.93; -1.12 (-3.94, 1.70) | 1.13, -1.44; -2.56 (-5.46, 0.33) | -0.07, -2.71; -2.65 (-5.44, 0.15) |
| Potassium (mmol/l) | 0.33, -0.04; -0.36 (-0.62, -0.11) | 0.03, 0.11; 0.08 (-0.16, 0.32) | 0.01, -0.11; -0.11 (-0.37, 0.14) | 0.17, 0.01; -0.16 (-0.43, 0.10) |
| Urea (mmol/l) | -0.04, -0.01; 0.04 (-0.87, 0.94) | -0.25, -0.69; -0.45 (-1.34, 0.45) | 0.72, 0.24; -0.49 (-1.84, 0.87) | 0.09, 0.32; 0.23 (-0.82, 1.29) |
| Creatinine (umol/l) | 5.71, 12.36; 6.64 (0.17, 13.12) | 1.19, 4.71; 3.53 (-0.64, 7.69) | 8.05, 8.72; 0.68 (-6.83, 8.18) | 4.54, 3.71; -0.82 (-7.40, 5.75) |
| Testosterone (nmol/l) | -0.76, -0.54; 0.21 (-1.52, 1.94) | -0.02, -0.51; -0.49 (-1.83, 0.86) | -0.84, -0.51; 0.33 (-1.61, 2.26) | -0.85, 0.69; 1.53 (-0.41, 3.48) |
| Dehydroepiandrosterone sulphate (umol/l) | 0.03, 2.00; 1.97 (1.04, 2.91) | -0.28, 2.44; 2.71 (1.98, 3.44) | -0.02, 1.99; 2.01 (1.14, 2.88) | -0.53, 0.64; 1.16 (0.51, 1.82) |
| Free thyroxine (pmol/l) | -0.24, 0.02; 0.26 (-0.89, 1.40) | -0.22, 0.01; 0.24 (-0.76, 1.23) | -0.47, -0.24; 0.23 (-0.94, 1.39) | -0.87, -0.63; 0.24 (-0.67, 1.15) |
| Thyroid stimulating hormone (mlU/l) | -0.06, 0.12; 0.18 (-0.17, 0.52) | 0.15, -0.01; -0.16 (-0.51, 0.18) | 0.03, -0.05; -0.08 (-0.48, 0.33) | 0.05, 0.04; -0.01 (-0.40, 0.37) |
| *Estimated in imputed data. | | | | |
