## Supplementary material for "A randomised controlled pilot trial of oral 11β-HSD1 inhibitor AZD4017 for wound healing in adults with type 2 diabetes mellitus": Table S14

#### Table S14: Adjusted differences in changes from baseline in longitudinal laboratory safety variables

Population: Safety set

Multiple imputation was used to address missing data. All point estimates and confidence intervals estimated via linear regression.

| **Variable** | **Mean* PCB, AZD; Difference AZD-PCB (90% CI) (total N=28)** | | | |
| --- | --- | --- | --- | --- |
|  | Day 7 | Day 28 | Day 35 | Day 42 |
| Body Mass Index (kg / m2) | N/A | N/A | 0.17, 0.58; 0.41 (-0.51, 1.34) | N/A |
| Waist-hip ratio | N/A | N/A | -0.01, 0.01; 0.02 (-0.01, 0.05) | N/A |
| Systolic blood pressure (mm Hg) | N/A | N/A | 5.61, -9.13; -14.74 (-23.00, -6.47) | 89.67, 90.44; 0.77 (-6.58, 8.11) |
| Diastolic blood pressure (mm Hg) | N/A | N/A | -0.25, -2.69; -2.43 (-7.67, 2.80) | 58.31, 60.40; 2.09 (-6.77, 10.95) |
| HbA1c (mmol/mol) | -0.38, -0.19; 0.18 (-1.51, 1.88) | -2.10, 0.08; 2.17 (-0.64, 4.99) | -1.71, -0.99; 0.72 (-3.64, 5.09) | -2.87, -0.45; 2.42 (-2.38, 7.22) |
| High density lipoprotein (mmol/l) | -0.08, -0.14; -0.06 (-0.14, 0.02) | 0.18, 0.10; -0.08 (-0.16, 0.00) | 0.10, -0.03; -0.13 (-0.21, -0.05) | 0.06, 0.11; 0.05 (-0.07, 0.16) |
| Cholesterol (mmol/l) | 0.00, -0.41; -0.42 (-0.60, -0.23) | 1.55, 1.10; -0.46 (-0.79, -0.12) | 0.97, 0.46; -0.51 (-0.87, -0.15) | 1.28, 1.18; -0.10 (-0.54, 0.34) |
| Triglycerides (mmol/l) | 0.47, 0.33; -0.14 (-0.67, 0.40) | 0.62, 0.43; -0.19 (-0.56, 0.18) | -0.03, -0.52; -0.48 (-1.21, 0.24) | 0.05, 0.07; 0.02 (-0.51, 0.55) |
| Haemoglobin (g/l) | -2.90, -2.24; 0.67 (-2.12, 3.45) | 45.08, 44.10; -0.98 (-4.13, 2.17) | 15.89, 16.53; 0.64 (-3.96, 5.24) | -3.55, -1.48; 2.07 (-1.62, 5.76) |
| White cells (x109/l) | 0.17, 0.71; 0.54 (-0.14, 1.22) | 1.76, 1.36; -0.40 (-0.87, 0.08) | 1.51, 1.21; -0.31 (-0.96, 0.34) | 1.07, 1.40; 0.33 (-0.41, 1.07) |
| Platelets (x109/l) | 0.10, 2.52; 2.42 (-13.65, 18.50) | 7.72, 21.04; 13.32 (-9.56, 36.20) | 9.47, 8.55; -0.91 (-25.20, 23.37) | 2.42, 3.89; 1.47 (-16.74, 19.68) |
| Red cells (x1012/l) | -0.09, -0.14; -0.05 (-0.17, 0.06) | 0.93, 0.88; -0.05 (-0.15, 0.04) | 0.42, 0.40; -0.02 (-0.15, 0.12) | -0.29, -0.15; 0.15 (0.03, 0.26) |
| Mean corpuscular volume (fl) | 1.06, 1.17; 0.11 (-1.83, 2.05) | 6.03, 6.55; 0.52 (-1.49, 2.53) | 9.58, 9.41; -0.17 (-2.53, 2.19) | 12.90, 13.06; 0.16 (-1.42, 1.73) |
| Haematocrit (packed cell volume) | -0.00, -0.01; -0.01 (-0.02, 0.01) | 0.12, 0.12; 0.00 (-0.01, 0.02) | 0.03, 0.03; 0.00 (-0.02, 0.02) | -0.02, -0.01; 0.02 (0.00, 0.03) |
| Mean corpuscular haemoglobin (pg) | -0.02, 0.39; 0.41 (0.08, 0.75) | 3.58, 3.51; -0.07 (-0.44, 0.31) | 1.34, 1.51; 0.17 (-0.28, 0.62) | 2.42, 2.00; -0.42 (-0.95, 0.10) |
| Corpuscular hemoglobin concentration (g/l) | -4.52, 0.39; 4.91 (-1.41, 11.22) | 150.85, 147.06; -3.80 (-10.80, 3.21) | 121.63, 123.38; 1.75 (-4.85, 8.35) | 62.90, 57.67; -5.22 (-12.18, 1.74) |
| Red blood cell distribution width (%) | 0.52, 0.17; -0.36 (-0.89, 0.18) | 6.04, 5.95; -0.09 (-0.49, 0.31) | 2.59, 2.70; 0.11 (-0.28, 0.51) | 2.84, 2.64; -0.21 (-0.68, 0.26) |
| Albumin (g/l) | 0.18, 0.54; 0.37 (-0.82, 1.55) | 10.87, 10.50; -0.37 (-1.82, 1.09) | 9.36, 9.35; -0.01 (-1.60, 1.59) | 11.80, 12.19; 0.40 (-0.59, 1.38) |
| Blirubin (umol/l) | -0.50, 0.43; 0.93 (-1.14, 3.00) | 1.36, 1.97; 0.60 (-0.89, 2.09) | 4.14, 3.50; -0.64 (-3.01, 1.73) | 2.16, 2.50; 0.35 (-1.17, 1.87) |
| Alkaline phosphatase (U/l) | 0.66, -5.34; -6.00 (-12.44, 0.44) | 18.33, 5.94; -12.39 (-20.21, -4.56) | 12.99, -0.10; -13.09 (-20.43, -5.75) | 8.43, 2.78; -5.65 (-15.25, 3.95) |
| Alanine aminotransferase (iu/l) | -2.09, -0.94; 1.16 (-1.84, 4.15) | 4.23, 2.27; -1.96 (-4.94, 1.03) | 5.83, 4.19; -1.64 (-5.88, 2.61) | 1.56, 2.34; 0.78 (-3.00, 4.56) |
| Aspartate aminotransferase (iu/l) | -0.33, 0.13; 0.46 (-2.29, 3.21) | 11.62, 11.74; 0.12 (-1.81, 2.05) | 6.93, 6.90; -0.04 (-2.52, 2.45) | 6.53, 8.74; 2.21 (-1.02, 5.44) |
| Gamma-glutamyl transpeptidase (iu/l) | -2.23, -1.47; 0.76 (-2.83, 4.35) | 10.09, -0.19; -10.28 (-14.96, -5.60) | 7.96, -4.02; -11.97 (-22.80, -1.15) | 6.46, -3.09; -9.55 (-14.88, -4.22) |
| eGFR (ml/min/1.73m2) | -4.42, -7.60; -3.18 (-8.30, 1.94) | 2.79, 0.60; -2.19 (-5.38, 1.01) | -14.06, -13.79; 0.27 (-4.57, 5.10) | 4.24, 5.86; 1.62 (-2.77, 6.01) |
| Sodium (mmol/l) | -0.23, -0.12; 0.10 (-1.23, 1.44) | 138.48, 138.64; 0.16 (-0.98, 1.29) | 1.57, 0.37; -1.19 (-3.21, 0.82) | 135.61, 134.37; -1.25 (-2.42, -0.07) |
| Potassium (mmol/l) | 0.30, -0.00; -0.31 (-0.58, -0.03) | 2.17, 2.35; 0.18 (-0.06, 0.43) | 1.61, 1.63; 0.01 (-0.22, 0.25) | 1.85, 1.80; -0.04 (-0.31, 0.22) |
| Urea (mmol/l) | -0.08, 0.04; 0.13 (-0.79, 1.05) | -0.33, -0.68; -0.35 (-1.31, 0.61) | -1.47, -1.99; -0.52 (-1.88, 0.83) | 0.16, 0.18; 0.02 (-1.04, 1.08) |
| Creatinine (umol/l) | 5.30, 12.74; 7.43 (0.79, 14.08) | -1.29, 2.14; 3.43 (-0.98, 7.85) | -8.65, -6.87; 1.78 (-5.65, 9.22) | 9.36, 7.78; -1.58 (-8.24, 5.07) |
| Testosterone (nmol/l) | -0.34, -0.97; -0.63 (-2.52, 1.25) | 0.86, -0.20; -1.06 (-2.58, 0.45) | 0.38, -0.02; -0.40 (-2.66, 1.85) | 0.44, 1.57; 1.12 (-1.20, 3.45) |
| Dehydroepiandrosterone sulphate (umol/l) | -0.13, 2.17; 2.30 (1.52, 3.08) | -0.50, 2.36; 2.86 (2.09, 3.64) | -0.15, 2.02; 2.16 (1.29, 3.04) | 0.32, 1.41; 1.09 (0.43, 1.75) |
| Free thyroxine (pmol/l) | -0.36, 0.14; 0.50 (-0.63, 1.64) | 5.32, 5.82; 0.51 (-0.43, 1.44) | 4.33, 4.99; 0.65 (-0.45, 1.76) | 3.77, 4.56; 0.79 (0.07, 1.52) |
| Thyroid stimulating hormone (mlU/l) | -0.02, 0.08; 0.10 (-0.22, 0.42) | 0.12, -0.07; -0.20 (-0.58, 0.19) | 0.52, 0.36; -0.15 (-0.54, 0.23) | 0.09, 0.05; -0.04 (-0.46, 0.38) |
| *Estimated in imputed data, adjusted for baseline value, age, sex and baseline HbA1c. | | | | |
