## Supplementary material for "A randomised controlled pilot trial of oral 11β-HSD1 inhibitor AZD4017 for wound healing in adults with type 2 diabetes mellitus": Table S15

### Table S15: Longitudinal laboratory safety variables; absolute and relative frequencies of values below or above normal limits

Population: Safety set

| **Variable** | **LLN** | **ULN** | **Summary** | **Day 0** | | **s1** | **Day 7** | | **s2** | **Day 28** | | **s3** | **Day 35** | | **s4** | **Day 42** | |
| --- | --- | --- | --- | --- | --- | --- | --- | --- | --- | --- | --- | --- | --- | --- | --- | --- | --- |
|  |  |  |  | **PCB** | **AZD** |  | **PCB** | **AZD** |  | **PCB** | **AZD** |  | **PCB** | **AZD** |  | **PCB** | **AZD** |
| BMI | N/A | N/A | Total, n | 14 | 14 |  |  |  |  |  |  |  | 13 | 13 |  |  |  |
| WHR | N/A | N/A | Total, n | 14 | 14 |  |  |  |  |  |  |  | 13 | 13 |  |  |  |
| BPSystolic | N/A | 150 | Total, n | 14 | 14 |  |  |  |  |  |  |  | 13 | 13 |  | 13 | 14 |
|  |  |  | Above normal, n | 3 | 2 |  |  |  |  |  |  |  | 4 | 1 |  | 3 |  |
|  |  |  | Above normal, % | 21 | 14 |  |  |  |  |  |  |  | 31 | 8 |  | 23 |  |
| BPDiastolic | N/A | 90 | Total, n | 14 | 14 |  |  |  |  |  |  |  | 13 | 13 |  | 13 | 14 |
|  |  |  | Above normal, n | 2 |  |  |  |  |  |  |  |  | 1 | 1 |  | 2 | 3 |
|  |  |  | Above normal, % | 14 |  |  |  |  |  |  |  |  | 8 | 8 |  | 15 | 21 |
| HbA1c | N/A | 41 | Total, n | 14 | 14 |  | 13 | 13 |  | 13 | 14 |  | 11 | 12 |  | 13 | 14 |
|  |  |  | Above normal, n | 14 | 14 |  | 13 | 12 |  | 13 | 14 |  | 11 | 12 |  | 13 | 14 |
|  |  |  | Above normal, % | 100 | 100 |  | 100 | 92 |  | 100 | 100 |  | 100 | 100 |  | 100 | 100 |
| HDL | 1.5 | N/A | Total, n | 14 | 14 |  | 14 | 14 |  | 13 | 14 |  | 12 | 12 |  | 12 | 13 |
|  |  |  | Below normal, n | 12 | 12 |  | 12 | 14 |  | 11 | 14 |  | 10 | 12 |  | 10 | 10 |
|  |  |  | Below normal, % | 86 | 86 |  | 86 | 100 |  | 85 | 100 |  | 83 | 100 |  | 83 | 77 |
| Cholesterol | 2.6 | 5.2 | Total, n | 14 | 14 |  | 14 | 14 |  | 13 | 14 |  | 12 | 12 |  | 12 | 13 |
|  |  |  | Below normal, n |  |  |  |  |  |  |  |  |  |  |  |  | 1 |  |
|  |  |  | Below normal, % |  |  |  |  |  |  |  |  |  |  |  |  | 8 |  |
|  |  |  | Above normal, n | 2 | 1 |  | 2 |  |  | 1 |  |  | 1 |  |  | 1 |  |
|  |  |  | Above normal, % | 14 | 7 |  | 14 |  |  | 8 |  |  | 8 |  |  | 8 |  |
| Triglycerides | N/A | N/A | Total, n | 14 | 14 |  | 14 | 14 |  | 13 | 14 |  | 12 | 12 |  | 12 | 13 |
| Haemoglobin | 114 | 160 | Total, n | 14 | 14 |  | 14 | 14 |  | 13 | 14 |  | 12 | 13 |  | 13 | 14 |
|  |  |  | Below normal, n | 1 |  |  |  |  |  | 1 |  |  | 1 |  |  | 1 |  |
|  |  |  | Below normal, % | 7 |  |  |  |  |  | 8 |  |  | 8 |  |  | 8 |  |
|  |  |  | Above normal, n | 1 | 1 |  | 1 | 1 |  |  | 1 |  |  | 1 |  |  | 1 |
|  |  |  | Above normal, % | 7 | 7 |  | 7 | 7 |  |  | 7 |  |  | 8 |  |  | 7 |
| WBC | 4 | 11 | Total, n | 14 | 14 |  | 14 | 14 |  | 13 | 14 |  | 12 | 13 |  | 13 | 14 |
|  |  |  | Below normal, n | 1 | 1 |  |  | 1 |  | 1 | 1 |  | 1 | 1 |  | 1 | 1 |
|  |  |  | Below normal, % | 7 | 7 |  |  | 7 |  | 8 | 7 |  | 8 | 8 |  | 8 | 7 |
|  |  |  | Above normal, n | 1 | 1 |  |  |  |  |  |  |  |  |  |  |  | 1 |
|  |  |  | Above normal, % | 7 | 7 |  |  |  |  |  |  |  |  |  |  |  | 7 |
| Platelets | 150 | 400 | Total, n | 14 | 14 |  | 14 | 14 |  | 13 | 14 |  | 12 | 13 |  | 13 | 14 |
|  |  |  | Below normal, n | 1 | 1 |  | 2 | 1 |  | 1 | 1 |  | 1 | 1 |  |  | 1 |
|  |  |  | Below normal, % | 7 | 7 |  | 14 | 7 |  | 8 | 7 |  | 8 | 8 |  |  | 7 |
|  |  |  | Above normal, n |  | 1 |  |  | 2 |  |  | 2 |  |  | 2 |  |  | 2 |
|  |  |  | Above normal, % |  | 7 |  |  | 14 |  |  | 14 |  |  | 15 |  |  | 14 |
| RBC | 3.8 | 5.8 | Total, n | 14 | 14 |  | 14 | 14 |  | 13 | 14 |  | 12 | 13 |  | 13 | 14 |
|  |  |  | Below normal, n |  | 1 |  |  | 1 |  |  | 1 |  |  | 1 |  |  | 1 |
|  |  |  | Below normal, % |  | 7 |  |  | 7 |  |  | 7 |  |  | 8 |  |  | 7 |
|  |  |  | Above normal, n |  |  |  |  |  |  |  |  |  |  |  |  |  | 1 |
|  |  |  | Above normal, % |  |  |  |  |  |  |  |  |  |  |  |  |  | 7 |
| MCV | 78 | 100 | Total, n | 14 | 14 |  | 14 | 14 |  | 13 | 14 |  | 12 | 13 |  | 13 | 14 |
|  |  |  | Below normal, n |  |  |  |  |  |  |  |  |  | 1 |  |  |  |  |
|  |  |  | Below normal, % |  |  |  |  |  |  |  |  |  | 8 |  |  |  |  |
|  |  |  | Above normal, n |  | 1 |  |  | 2 |  |  | 1 |  |  | 1 |  |  | 1 |
|  |  |  | Above normal, % |  | 7 |  |  | 14 |  |  | 7 |  |  | 8 |  |  | 7 |
| PCV | .37 | .47 | Total, n | 14 | 14 |  | 14 | 14 |  | 13 | 14 |  | 12 | 13 |  | 13 | 14 |
|  |  |  | Below normal, n |  |  |  |  |  |  |  |  |  | 1 |  |  | 1 | 1 |
|  |  |  | Below normal, % |  |  |  |  |  |  |  |  |  | 8 |  |  | 8 | 7 |
|  |  |  | Above normal, n | 3 | 1 |  | 2 | 2 |  | 1 | 1 |  | 1 | 2 |  |  | 2 |
|  |  |  | Above normal, % | 21 | 7 |  | 14 | 14 |  | 8 | 7 |  | 8 | 15 |  |  | 14 |
| MCH | 27 | 232 | Total, n | 14 | 14 |  | 14 | 14 |  | 13 | 14 |  | 12 | 13 |  | 13 | 14 |
|  |  |  | Below normal, n | 2 | 2 |  | 2 | 2 |  | 1 | 2 |  | 1 | 2 |  | 1 | 3 |
|  |  |  | Below normal, % | 14 | 14 |  | 14 | 14 |  | 8 | 14 |  | 8 | 15 |  | 8 | 21 |
|  |  |  | Above normal, n |  |  |  |  |  |  |  |  |  |  |  |  |  |  |
|  |  |  | Above normal, % |  |  |  |  |  |  |  |  |  |  |  |  |  |  |
| MCH2 | N/A | N/A | Total, n | 14 | 14 |  | 14 | 14 |  | 13 | 14 |  | 12 | 13 |  | 13 | 14 |
| RBCDW | 11.5 | 15 | Total, n | 14 | 14 |  | 14 | 14 |  | 13 | 14 |  | 12 | 13 |  | 13 | 14 |
|  |  |  | Below normal, n |  |  |  |  |  |  |  |  |  |  |  |  |  |  |
|  |  |  | Below normal, % |  |  |  |  |  |  |  |  |  |  |  |  |  |  |
|  |  |  | Above normal, n | 1 | 4 |  | 3 | 3 |  | 1 | 1 |  | 3 | 3 |  | 1 | 1 |
|  |  |  | Above normal, % | 7 | 29 |  | 21 | 21 |  | 8 | 7 |  | 25 | 23 |  | 8 | 7 |
| Albumin | 35 | 50 | Total, n | 14 | 14 |  | 14 | 14 |  | 13 | 14 |  | 12 | 13 |  | 13 | 13 |
|  |  |  | Below normal, n |  |  |  |  |  |  | 1 |  |  |  |  |  |  |  |
|  |  |  | Below normal, % |  |  |  |  |  |  | 8 |  |  |  |  |  |  |  |
|  |  |  | Above normal, n |  |  |  |  |  |  |  |  |  |  |  |  |  |  |
|  |  |  | Above normal, % |  |  |  |  |  |  |  |  |  |  |  |  |  |  |
| Bilirubin | 2 | 21 | Total, n | 14 | 14 |  | 14 | 14 |  | 13 | 14 |  | 12 | 13 |  | 13 | 13 |
|  |  |  | Below normal, n |  |  |  |  |  |  |  |  |  |  |  |  |  |  |
|  |  |  | Below normal, % |  |  |  |  |  |  |  |  |  |  |  |  |  |  |
|  |  |  | Above normal, n |  |  |  |  | 1 |  |  | 1 |  | 1 |  |  |  |  |
|  |  |  | Above normal, % |  |  |  |  | 7 |  |  | 7 |  | 8 |  |  |  |  |
| ALP | 30 | 130 | Total, n | 14 | 14 |  | 14 | 14 |  | 13 | 14 |  | 12 | 13 |  | 13 | 13 |
|  |  |  | Below normal, n |  |  |  |  |  |  |  | 1 |  |  | 1 |  |  |  |
|  |  |  | Below normal, % |  |  |  |  |  |  |  | 7 |  |  | 8 |  |  |  |
|  |  |  | Above normal, n | 1 |  |  | 1 |  |  |  |  |  |  |  |  | 1 | 1 |
|  |  |  | Above normal, % | 7 |  |  | 7 |  |  |  |  |  |  |  |  | 8 | 8 |
| ALT | N/A | 40 | Total, n | 14 | 14 |  | 14 | 14 |  | 13 | 14 |  | 12 | 13 |  | 13 | 13 |
|  |  |  | Above normal, n |  | 2 |  |  | 1 |  |  | 2 |  |  | 1 |  |  | 2 |
|  |  |  | Above normal, % |  | 14 |  |  | 7 |  |  | 14 |  |  | 8 |  |  | 15 |
| AST | N/A | 40 | Total, n | 14 | 14 |  | 14 | 14 |  | 13 | 14 |  | 12 | 12 |  | 12 | 11 |
|  |  |  | Above normal, n |  |  |  |  |  |  |  |  |  |  |  |  |  |  |
|  |  |  | Above normal, % |  |  |  |  |  |  |  |  |  |  |  |  |  |  |
| GGT_M | N/A | 90 | Total, n | 12 | 10 |  | 12 | 10 |  | 11 | 10 |  | 10 | 9 |  | 11 | 9 |
|  |  |  | Above normal, n | 1 | 1 |  | 1 | 1 |  | 1 | 1 |  | 1 |  |  | 1 |  |
|  |  |  | Above normal, % | 8 | 10 |  | 8 | 10 |  | 9 | 10 |  | 10 |  |  | 9 |  |
| GGT_F | N/A | 50 | Total, n | 2 | 4 |  | 2 | 4 |  | 2 | 4 |  | 2 | 4 |  | 2 | 4 |
|  |  |  | Above normal, n | 1 |  |  | 1 | 1 |  | 1 |  |  | 1 |  |  | 1 |  |
|  |  |  | Above normal, % | 50 |  |  | 50 | 25 |  | 50 |  |  | 50 |  |  | 50 |  |
| eGFR | 90 | N/A | Total, n | 14 | 14 |  | 14 | 14 |  | 13 | 14 |  | 12 | 13 |  | 13 | 14 |
|  |  |  | Below normal, n | 9 | 8 |  | 9 | 12 |  | 8 | 9 |  | 7 | 8 |  | 9 | 8 |
|  |  |  | Below normal, % | 64 | 57 |  | 64 | 86 |  | 62 | 64 |  | 58 | 62 |  | 69 | 57 |
| Sodium | 133 | 146 | Total, n | 14 | 14 |  | 14 | 14 |  | 13 | 14 |  | 12 | 13 |  | 13 | 14 |
|  |  |  | Below normal, n |  |  |  |  |  |  |  |  |  |  |  |  |  |  |
|  |  |  | Below normal, % |  |  |  |  |  |  |  |  |  |  |  |  |  |  |
|  |  |  | Above normal, n |  | 1 |  |  |  |  |  |  |  | 1 |  |  |  |  |
|  |  |  | Above normal, % |  | 7 |  |  |  |  |  |  |  | 8 |  |  |  |  |
| Potassium | 3.5 | 5.3 | Total, n | 14 | 14 |  | 14 | 14 |  | 13 | 14 |  | 12 | 13 |  | 13 | 12 |
|  |  |  | Below normal, n |  |  |  |  |  |  |  |  |  |  |  |  |  |  |
|  |  |  | Below normal, % |  |  |  |  |  |  |  |  |  |  |  |  |  |  |
|  |  |  | Above normal, n |  | 1 |  | 3 |  |  |  | 2 |  |  |  |  | 1 |  |
|  |  |  | Above normal, % |  | 7 |  | 21 |  |  |  | 14 |  |  |  |  | 8 |  |
| Urea | 2.5 | 7.8 | Total, n | 14 | 14 |  | 14 | 14 |  | 13 | 14 |  | 12 | 13 |  | 13 | 14 |
|  |  |  | Below normal, n |  |  |  |  |  |  |  |  |  |  |  |  |  |  |
|  |  |  | Below normal, % |  |  |  |  |  |  |  |  |  |  |  |  |  |  |
|  |  |  | Above normal, n | 3 | 5 |  | 4 | 5 |  | 3 | 4 |  | 2 | 7 |  | 3 | 5 |
|  |  |  | Above normal, % | 21 | 36 |  | 29 | 36 |  | 23 | 29 |  | 17 | 54 |  | 23 | 36 |
| Creatinine_M | 64 | 104 | Total, n | 12 | 10 |  | 12 | 10 |  | 11 | 10 |  | 10 | 9 |  | 11 | 10 |
|  |  |  | Below normal, n | 2 | 3 |  | 2 | 1 |  | 2 | 1 |  | 1 | 1 |  | 1 | 2 |
|  |  |  | Below normal, % | 17 | 30 |  | 17 | 10 |  | 18 | 10 |  | 10 | 11 |  | 9 | 20 |
|  |  |  | Above normal, n |  | 2 |  | 2 | 3 |  | 1 | 2 |  | 2 | 3 |  | 1 | 1 |
|  |  |  | Above normal, % |  | 20 |  | 17 | 30 |  | 9 | 20 |  | 20 | 33 |  | 9 | 10 |
| Creatinine_F | 49 | 90 | Total, n | 2 | 4 |  | 2 | 4 |  | 2 | 4 |  | 2 | 4 |  | 2 | 4 |
|  |  |  | Below normal, n |  |  |  |  |  |  |  |  |  |  |  |  |  |  |
|  |  |  | Below normal, % |  |  |  |  |  |  |  |  |  |  |  |  |  |  |
|  |  |  | Above normal, n |  |  |  |  |  |  |  |  |  |  | 1 |  |  | 1 |
|  |  |  | Above normal, % |  |  |  |  |  |  |  |  |  |  | 25 |  |  | 25 |
| Testosterone_M | 8 | 30 | Total, n | 12 | 10 |  | 12 | 10 |  | 11 | 10 |  | 10 | 9 |  | 11 | 10 |
|  |  |  | Below normal, n | 1 | 2 |  | 2 | 4 |  | 2 | 4 |  | 1 | 3 |  | 3 | 2 |
|  |  |  | Below normal, % | 8 | 20 |  | 17 | 40 |  | 18 | 40 |  | 10 | 33 |  | 27 | 20 |
|  |  |  | Above normal, n |  |  |  |  |  |  |  |  |  | 1 |  |  |  |  |
|  |  |  | Above normal, % |  |  |  |  |  |  |  |  |  | 10 |  |  |  |  |
| Testosterone_F | N/A | 2.8 | Total, n | 2 | 4 |  | 2 | 4 |  | 1 | 4 |  | 2 | 4 |  | 2 | 4 |
|  |  |  | Above normal, n |  |  |  |  |  |  |  |  |  |  |  |  |  |  |
|  |  |  | Above normal, % |  |  |  |  |  |  |  |  |  |  |  |  |  |  |
| DHEAS_M | 3.6 | 13 | Total, n | 12 | 10 |  | 12 | 10 |  | 11 | 10 |  | 10 | 9 |  | 11 | 10 |
|  |  |  | Below normal, n | 5 | 7 |  | 5 | 4 |  | 6 | 2 |  | 4 | 2 |  | 8 | 6 |
|  |  |  | Below normal, % | 42 | 70 |  | 42 | 40 |  | 55 | 20 |  | 40 | 22 |  | 73 | 60 |
|  |  |  | Above normal, n |  |  |  |  | 1 |  |  |  |  |  |  |  |  |  |
|  |  |  | Above normal, % |  |  |  |  | 10 |  |  |  |  |  |  |  |  |  |
| DHEAS_F | 2.7 | 11 | Total, n | 2 | 4 |  | 2 | 4 |  | 2 | 4 |  | 2 | 4 |  | 2 | 4 |
|  |  |  | Below normal, n | 2 | 3 |  | 2 | 1 |  | 2 | 2 |  | 2 | 2 |  | 2 | 3 |
|  |  |  | Below normal, % | 100 | 75 |  | 100 | 25 |  | 100 | 50 |  | 100 | 50 |  | 100 | 75 |
|  |  |  | Above normal, n |  |  |  |  |  |  |  |  |  |  |  |  |  |  |
|  |  |  | Above normal, % |  |  |  |  |  |  |  |  |  |  |  |  |  |  |
| fT4 | 10 | 20 | Total, n | 14 | 14 |  | 14 | 13 |  | 12 | 12 |  | 12 | 13 |  | 13 | 14 |
|  |  |  | Below normal, n |  |  |  |  |  |  |  |  |  |  |  |  |  |  |
|  |  |  | Below normal, % |  |  |  |  |  |  |  |  |  |  |  |  |  |  |
|  |  |  | Above normal, n |  |  |  |  | 1 |  |  |  |  |  |  |  |  |  |
|  |  |  | Above normal, % |  |  |  |  | 8 |  |  |  |  |  |  |  |  |  |
| TSH | .2 | 4 | Total, n | 14 | 14 |  | 14 | 13 |  | 12 | 12 |  | 12 | 13 |  | 13 | 14 |
|  |  |  | Below normal, n |  |  |  |  |  |  |  |  |  |  |  |  |  |  |
|  |  |  | Below normal, % |  |  |  |  |  |  |  |  |  |  |  |  |  |  |
|  |  |  | Above normal, n |  |  |  |  |  |  |  |  |  |  |  |  |  |  |
|  |  |  | Above normal, % |  |  |  |  |  |  |  |  |  |  |  |  |  |  |
| LLN=Lower limit of normal; ULN=Upper limit of normal. Suffix F=Female; M=Male. | | | | | | | | | | | | | | | | | |
