## Supplementary material for "A randomised controlled pilot trial of oral 11β-HSD1 inhibitor AZD4017 for wound healing in adults with type 2 diabetes mellitus": Table S16

### Table S16: Sample sizes for future trials

Based on estimates from available case data in full analysis set.

At alpha=0.05 (5% significance), 1-Beta=0.90 (90% power), accounting for 10% drop-out, sample sizes for a range of substantive between-group differences are presented below. For TEWL and integrity variables, means and SDs are presented on the log scale.

Note that for all variables, sample size has been based on mean and SD (and Pearson's r where applicable), as specified in the statistical analysis plan; issues with distributions for some variables may reduce the accuracy of these estimates. Sample sizes presented here should be considered preliminary.

| **Variable** | **Relative difference** | **PCB mean** | **AZD mean** | **Pooled SD** | **R*** | **N per arm** |
| --- | --- | --- | --- | --- | --- | --- |
| 11bHSD1 activity radioassay (% conv/24hrs): Day 28 | 10% | 12.38 | 11.14 | 5.13 | 0.07 | 399 |
|  | 20% | 12.38 | 9.90 | 5.13 | 0.07 | 100 |
|  | 30% | 12.38 | 8.66 | 5.13 | 0.07 | 46 |
| 11bHSD1 activity ELISA (% conv/24hrs): Day 28 | 10% | 15.27 | 13.74 | 25.85 | 0.36 | 5821 |
|  | 20% | 15.27 | 12.22 | 25.85 | 0.36 | 1456 |
|  | 30% | 15.27 | 10.69 | 25.85 | 0.36 | 648 |
| Sudomotor function Left Hand (micro S): Day 35 | 10% | 56.46 | 62.11 | 12.83 | 0.42 | 100 |
|  | 20% | 56.46 | 67.75 | 12.83 | 0.42 | 26 |
|  | 30% | 56.46 | 73.40 | 12.83 | 0.42 | 12 |
| Sudomotor function Right Hand (micro S): Day 35 | 10% | 54.08 | 59.48 | 14.07 | 0.61 | 100 |
|  | 20% | 54.08 | 64.89 | 14.07 | 0.61 | 26 |
|  | 30% | 54.08 | 70.30 | 14.07 | 0.61 | 11 |
| Sudomotor function Hands (micro S): Day 35 | 10% | 55.27 | 60.80 | 13.26 | 0.52 | 99 |
|  | 20% | 55.27 | 66.32 | 13.26 | 0.52 | 26 |
|  | 30% | 55.27 | 71.85 | 13.26 | 0.52 | 11 |
| Sudomotor function Left Foot (micro S): Day 35 | 10% | 68.69 | 75.56 | 15.26 | 0.67 | 63 |
|  | 20% | 68.69 | 82.43 | 15.26 | 0.67 | 16 |
|  | 30% | 68.69 | 89.30 | 15.26 | 0.67 | 8 |
| Sudomotor function Right Foot (micro S): Day 35 | 10% | 69.31 | 76.24 | 16.69 | 0.77 | 56 |
|  | 20% | 69.31 | 83.17 | 16.69 | 0.77 | 14 |
|  | 30% | 69.31 | 90.10 | 16.69 | 0.77 | 7 |
| Sudomotor function Feet (micro S): Day 35 | 10% | 69.00 | 75.90 | 15.82 | 0.72 | 59 |
|  | 20% | 69.00 | 82.80 | 15.82 | 0.72 | 14 |
|  | 30% | 69.00 | 89.70 | 15.82 | 0.72 | 7 |
| Sudomotor function Overall (micro S): Day 35 | 10% | 62.13 | 68.35 | 12.82 | 0.70 | 50 |
|  | 20% | 62.13 | 74.56 | 12.82 | 0.70 | 13 |
|  | 30% | 62.13 | 80.78 | 12.82 | 0.70 | 6 |
| Skin hydration (A.U): Day 35 | 10% | 38.19 | 42.01 | 9.90 | 0.55 | 110 |
|  | 20% | 38.19 | 45.83 | 9.90 | 0.55 | 28 |
|  | 30% | 38.19 | 49.65 | 9.90 | 0.55 | 12 |
| Epidermal thickness (micro m): Day 35 | 10% | 61.72 | 67.90 | 9.43 | 0.07 | 56 |
|  | 20% | 61.72 | 74.07 | 9.43 | 0.07 | 14 |
|  | 30% | 61.72 | 80.24 | 9.43 | 0.07 | 7 |
| Wound gap diameter (mm): Day 2 | 10% | 1.49 | 1.34 | 0.71 | N/A | 529 |
|  | 20% | 1.49 | 1.19 | 0.71 | N/A | 132 |
|  | 30% | 1.49 | 1.04 | 0.71 | N/A | 59 |
| Wound depth (mm): Day 7 | 10% | 0.60 | 0.54 | 0.20 | N/A | 262 |
|  | 20% | 0.60 | 0.48 | 0.20 | N/A | 66 |
|  | 30% | 0.60 | 0.42 | 0.20 | N/A | 30 |
| Wound gap diameter (mm): Day 30 | 10% | 1.44 | 1.30 | 0.60 | N/A | 409 |
|  | 20% | 1.44 | 1.16 | 0.60 | N/A | 102 |
|  | 30% | 1.44 | 1.01 | 0.60 | N/A | 46 |
| Wound depth (mm): Day 35 | 10% | 0.60 | 0.54 | 0.19 | N/A | 233 |
|  | 20% | 0.60 | 0.48 | 0.19 | N/A | 59 |
|  | 30% | 0.60 | 0.42 | 0.19 | N/A | 27 |
| Hour 3 TEWL (Set 1; Day 0) | 10% | 3.55 | 3.45 | 0.24 | 0.25 | 119 |
|  | 20% | 3.55 | 3.33 | 0.24 | 0.25 | 27 |
|  | 30% | 3.55 | 3.20 | 0.24 | 0.25 | 10 |
| Hour 48 TEWL (Set 1; Day 2) | 10% | 2.99 | 2.88 | 0.34 | 0.32 | 220 |
|  | 20% | 2.99 | 2.76 | 0.34 | 0.32 | 50 |
|  | 30% | 2.99 | 2.63 | 0.34 | 0.32 | 20 |
| Hour 168 TEWL (Set 1; Day 7) | 10% | 2.61 | 2.50 | 0.55 | 0.40 | 541 |
|  | 20% | 2.61 | 2.38 | 0.55 | 0.40 | 121 |
|  | 30% | 2.61 | 2.25 | 0.55 | 0.40 | 48 |
| Hour 3 TEWL (Set 2; Day 28) | 10% | 3.17 | 3.06 | 0.41 | 0.12 | 351 |
|  | 20% | 3.17 | 2.95 | 0.41 | 0.12 | 79 |
|  | 30% | 3.17 | 2.81 | 0.41 | 0.12 | 31 |
| Hour 48 TEWL (Set 2; Day 30) | 10% | 2.67 | 2.56 | 0.41 | 0.26 | 329 |
|  | 20% | 2.67 | 2.45 | 0.41 | 0.26 | 73 |
|  | 30% | 2.67 | 2.31 | 0.41 | 0.26 | 29 |
| Hour 168 TEWL (Set 2; Day 35) | 10% | 2.30 | 2.20 | 0.40 | 0.17 | 334 |
|  | 20% | 2.30 | 2.08 | 0.40 | 0.17 | 74 |
|  | 30% | 2.30 | 1.95 | 0.40 | 0.17 | 29 |
| Hour 0 TEWL (Set 3; Day 35) | 10% | 1.94 | 1.84 | 0.42 | -0.12 | 374 |
|  | 20% | 1.94 | 1.72 | 0.42 | -0.12 | 84 |
|  | 30% | 1.94 | 1.59 | 0.42 | -0.12 | 33 |
| N tapes required for barrier disruption: Day 28 | 10% | 3.63 | 3.73 | 0.49 | 0.42 | 500 |
|  | 20% | 3.63 | 3.82 | 0.49 | 0.42 | 138 |
|  | 30% | 3.63 | 3.90 | 0.49 | 0.42 | 66 |
| *Correlation with baseline value (not applicable for WH measures) | | | | | | |
