## Supplementary figures and images for "A randomised controlled pilot trial of oral 11β-HSD1 inhibitor AZD4017 for wound healing in adults with type 2 diabetes mellitus"

### Figure S1

Figure S1

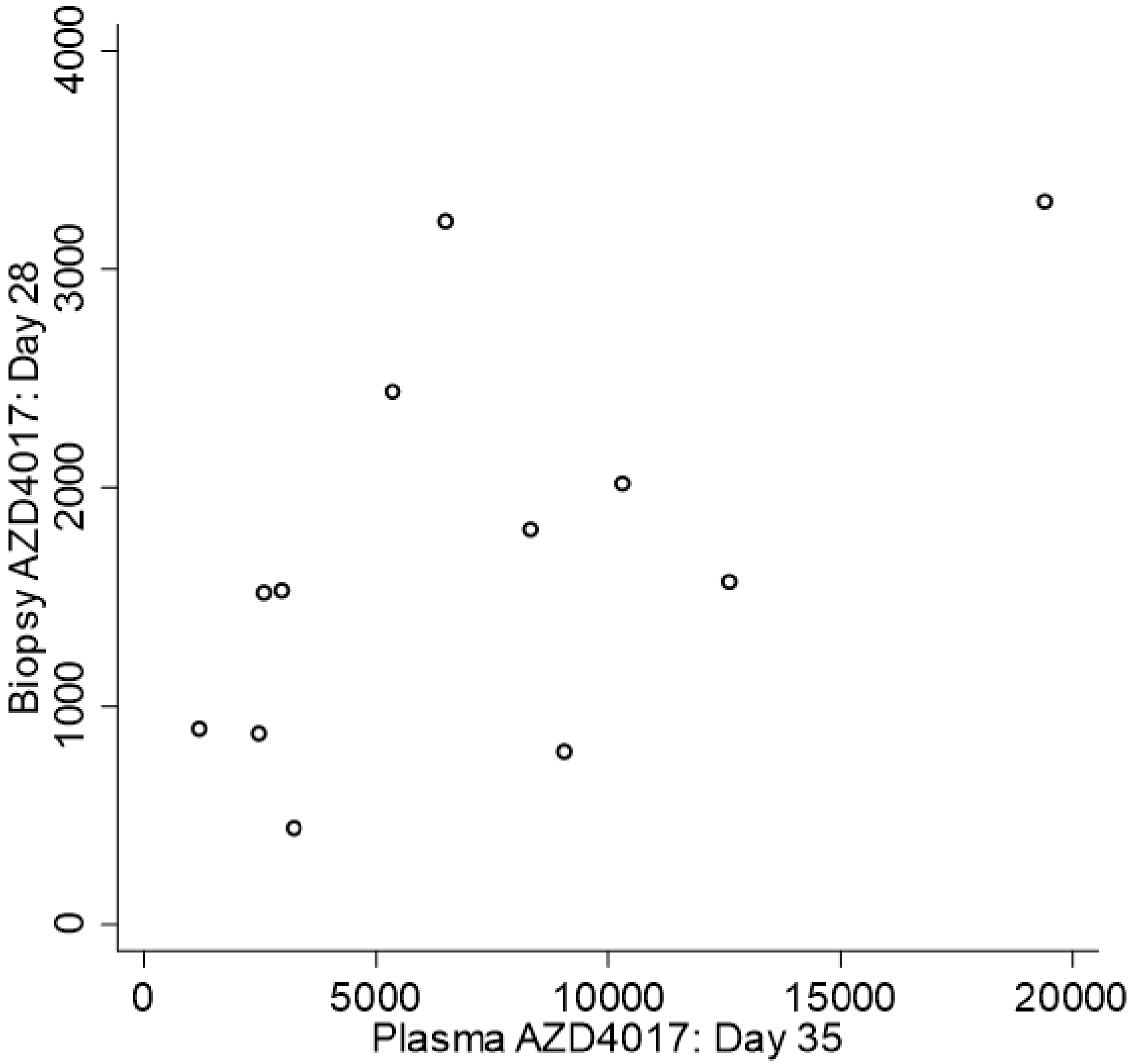


rho=0.54

### Figure S2

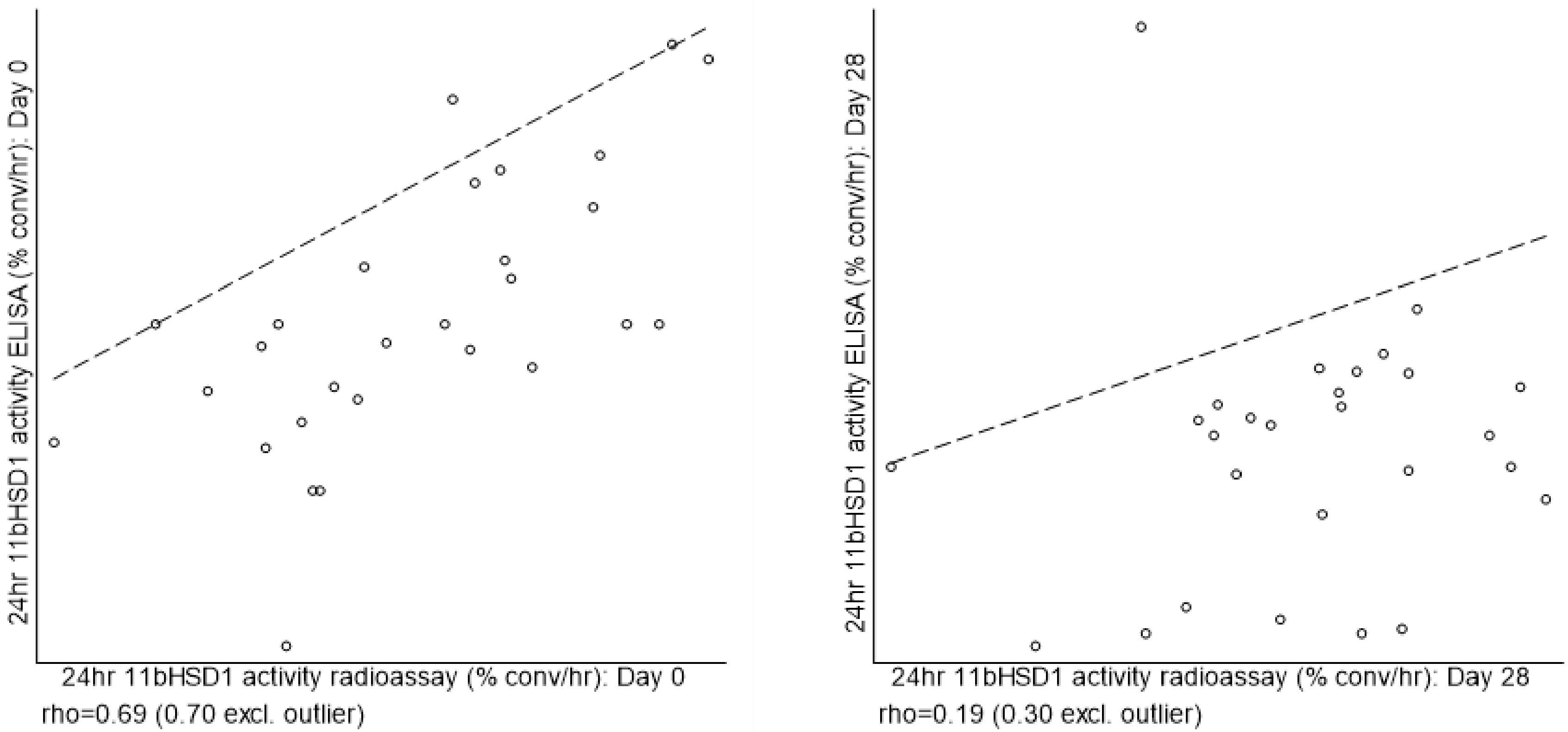
