## Appendix 1 for "A randomised controlled pilot trial of oral 11β-HSD1 inhibitor AZD4017 for wound healing in adults with type 2 diabetes mellitus"

### APPENDIX 1: Feasibility Results

#### 1.1 Recruitment

Limitations to recruitment were a moderate factor, requiring two extensions. The decision to stop recruitment at n=28 was justified on grounds that due to lower-than-anticipated drop-out at least 12 patients had completed follow-up.

The average recruitment rate was 2.9 patients per month; however, recruitment was intermittent rather than steady. Limited staffing (due to restricted resources for this early phase study) reduced the number of participants that could be processed simultaneously; some visits lasted ~4 hours and had a stringent schedule. This could be improved in future a future trial by focusing on a smaller number of outcome measures to reduce visit duration and increasing the number of assessors to include more participants simultaneously.

Despite this limitation, trial duration from funding confirmation to last patient last visit was 31 months.

#### 1.2 Eligibility and consent

Eligibility was a modest barrier to recruitment, with 8 screen failures (22%; 90% CI=13,35). Study uptake was also relatively low with approximately 300 study invitations being issued to attain 36 screening visits. Reasons for declining included inability to commit to the study schedule (e.g. childcare responsibilities or work) – this could be improved in future studies by coordinating study visits with clinic appointments. Of the 36 patients screened, 28 were eligible, all of whom consented to proceed with the trial.

#### 1.3 Data completeness

Overall data completeness was excellent, achieving 5,894/6,184 expected reportable data points (95.3%). The largest source of missing data (58.6%) was missed study visits, although these were rare (5/196 or 2.6%). Of the remaining missed data points 31.7% were due to incorrect blood sample processing. However, these were relatively low as a proportion of all blood data points - occurring in less than 1% of cases. A further 17.5% were due to patients forgetting to bring their diary card or tablets to the visit - affecting 7.5% of cases. The last missed data points affected a range of outcomes e.g. TEWL processing issues due to the warm weather, issues with OCT machine repair and more isolated incidents of human error. No particular outcome measure was deemed to be a cause of data incompleteness and all were deemed viable or future study inclusion.
