## Appendix 2 for "A randomised controlled pilot trial of oral 11β-HSD1 inhibitor AZD4017 for wound healing in adults with type 2 diabetes mellitus"

### Research Protocol

**Version 2.0 - 24/10/2018**

---

**Study Short Title: GC-SHealD (Glucocorticoids and Skin Healing in Diabetes)**

**Study Full Title: A double-blind, randomized, placebo-controlled phase II pilot trial investigating efficacy, safety and feasibility of 11β-hydroxysteroid dehydrogenase type 1 inhibition by AZD4017 to improve skin function and wound healing in patients with type 2 diabetes**

**Sponsor Name: University of Leeds**

**Sponsor Number: ED17/93260**

**EudraCT Number: 2017-001351-31**

**ISRCTN: 74621291**

---

**Protocol status:** FINAL

**Details of previous amendments**

**Version 1.2 11/07/2017**

---

**Confidential**. Intellectual property: University of Leeds.

### Key contacts

#### Chief Investigator

Dr Ramzi Ajjan

Associate Professor and Consultant in Diabetes and Endocrinology

Division of Cardiovascular and Diabetes Research

0113 343 7475

#### Scientific Lead

Ana Tiganescu

Postdoctoral Research Fellow

Leeds Institute of Rheumatic and Musculoskeletal Medicine

0113 34 38336

**Confidential**. Intellectual property: University of Leeds.

### INVESTIGATOR DECLARATION AND SIGNATURE(S)

**GC-SHealD Version 2.0, 24th October 2018**

**DECLARATION OF PROTOCOL ACCEPTANCE**

I confirm that I am fully informed and aware of the requirements of the protocol and agree to conduct the study as set out in this protocol.

| | |
|---|---|
| Ramzi Ajjan | |
| Chief Investigator | Date |

| | |
|---|---|
| Ana Tiganescu | |
| Scientific Lead | Date |

| | |
|---|---|
| Francesco Del Galdo | |
| Investigator | Date |

| | |
|---|---|
| Abd Tahrani | |
| Investigator | Date |

**Confidential**. Intellectual property: University of Leeds.

**Confidential**. Intellectual property: University of Leeds.

### ABBREVIATIONS

| Abbreviation | Term |
|---|---|
| 11β-HSD(1/2) | 11β-hydroxysteroid dehydrogenase (type 1/type 2) |
| ACTH | Adrenocorticotropic hormone |
| AE | Adverse event |
| ALP | Alkaline phosphatase |
| ALT | Alanine aminotransferase |
| AST | Aspartate aminotransferase |
| BP | Blood pressure |
| BMI | Body mass index |
| CBX | Carbenoxolone |
| CI | Chief Investigator |
| CK | Creatine kinase |
| CRF | Case report form |
| CTCAE | Common Terminology Criteria for Adverse Events |
| DFU | Diabetic foot ulcer |
| ECG | Electrocardiogram |
| ECM | Extracellular matrix |
| FBC | Full blood count |
| GC | Glucocorticoid |
| GC/MS | Gas chromatography/mass spectrometry |
| GCP | Good clinical practice |
| GMP | Good manufacturing practice |
| GP | General practitioner |
| GR | Glucocorticoid receptor |
| H6PDH | Hexose-6-phosphate dehydrogenase |
| HPA | Hypothalamic-pituitary-adrenal |
| IB | Investigator brochure |
| ICH | International conference on harmonisation |
| IMP | Investigational medicinal product |
| LFT | Liver function test |
| LIRMM | Leeds Institute of Rheumatic and Musculoskeletal Medicine |
| LMBRU | Leeds Musculoskeletal Biomedical Research Unit |

**Confidential**. Intellectual property: University of Leeds.

| Abbreviation | Term |
|---|---|
| MHRA | Medicines and healthcare products regulatory agency |
| MRI | Magnetic resonance imaging |
| PIL | Participant Information Leaflet |
| QA | Quality assurance |
| REC | Research ethics committee |
| SAE | Serious adverse event |
| SD | Standard deviation |
| SJUH | St James' University Hospital |
| SUSAR | Suspected unexpected serious adverse reaction |
| T2DM | Type 2 diabetes mellitus |
| [THF+alloTHF]/THE | Tetrahydrocortisol to tetrahydrocortisone urinary metabolites |
| TEWL | Trans-epidermal water loss |
| TMFTSH | Trial Master File |
| U+E | Thyroid stimulating hormone |
| ULN | Urea and electrolytes (kidney function test) |
| | Upper limit of normal |
| WH | Wound healing |
| WTBB | Wellcome Trust Brenner Building |

**Confidential**. Intellectual property: University of Leeds.

### PROTOCOL SYNOPSIS

| GENERAL INFORMATION | |
|---|---|
| Short Title | GC-SHealD (Glucocorticoids and Skin Healing in Diabetes) |
| Full Title | A double-blind, randomized, placebo-controlled phase II pilot trial investigating efficacy, safety and feasibility of 11β-hydroxysteroid dehydrogenase type 1 inhibition by AZD4017 to improve skin function and wound healing in patients with type 2 diabetes |
| Sponsor | University of Leeds |
| Sponsor ID | ED17/93260 |
| IRAS No. | 215411 |
| EudraCT No. | |
| MREC No. | |
| Chief Investigator | Ramzi Ajjan |
| Co-ordinating Centre | University of Leeds |
| National / International | Single site |
| **TRIAL INFORMATION** | |
| Phase | Phase II pilot |
| Indication | Type 2 diabetes mellitus |
| Design | Placebo-controlled, double-blind, parallel-group, randomised controlled pilot trial |
| Number of sites | 1 |
| Primary Objective | Evaluate AZD4017 efficacy |
| Secondary Objective(s) | 1) Evaluate safety of AZD4017<br>2) Examine effects of AZD4017 on skin function<br>3) Assess study feasibility |
| Primary Endpoint | 24 hour 11β-HSD1 activity in skin at baseline and day 28 |
| Secondary Endpoint(s) | Systemic 11β-HSD1 activity at baseline and day 35, AZD4017 detection in skin at day 28 and plasma at day 35, safety variables at baseline and days 7, 28 and 35, urinary cortisol to cortisone metabolite analysis at baseline and day 35, skin function variables at baseline and days 2, 7, 28, 30 and 35 and feasibility measures throughout the study |
| **TRIAL TIMELINES** | |
| Expected start date | Jun 2017 |

**Confidential**. Intellectual property: University of Leeds.

| | |
|---|---|
| Subject enrolment phase | Jul 2017 to Jan 2018 |
| Follow-up duration | One visit at least 7 days after the last treatment dose |
| End of Trial Definition | Entry of the last participant's last data item |
| Expected completion date | Feb 2018 |
| **TRIAL SUBJECT INFORMATION** | |
| Number of trial subjects | 30 |
| Age group of trial subjects | >18 years old |
| Inclusion criteria | 1) Able and willing to consent 2) T2DM with HbA1c ≤11% (≤97 mmol/mol) at screening while taking standard therapy at a stable dose for ≥10 weeks |
| Exclusion criteria | 1) WOCBP, 2) Active leg/foot ulceration, 3) Clinically relevant acute ECG anomalies 4) Uncontrolled hypertension, 5) Endocrine disorder (other than T2DM), including type 1 or secondary diabetes (except treated hypothyroidism), 6) Gilbert's disease, 7) Alanine aminotransferase and/or aspartate aminotransferase and/or alkaline phosphatase >1.5x ULN, 8) Bilirubin >1.5x ULN, 9) eGFR <45 ml/min/m$^2$, 10) CK >2x ULN, 11) Drug abuse within the last year, 12) Any GC treatment within 3 months of screening, 13) Anti-coagulant medication, 14) Probenecid therapy, 15) Medical/surgical procedure or trauma during IMP administration or one week after IMP cessation (excluding skin biopsies), 16) Involvement in trial planning and/or conduct, 17) Participation in other clinical study within 1 month, 18) Deemed inappropriate to participate by the trial team |
| **IMP** | |
| IMP name(s) | AZD4017 |
| IMP mode of administration | 400mg oral twice daily |
| Duration of IMP Treatment | 35 days |
| IMP Supplier(s) | AstraZeneca |
| Non IMP name(s) | Placebo |

**Confidential**. Intellectual property: University of Leeds.

#### SCHEMATIC DIAGRAM OF RECRUITMENT, RANDOMIZATION & TREATMENT

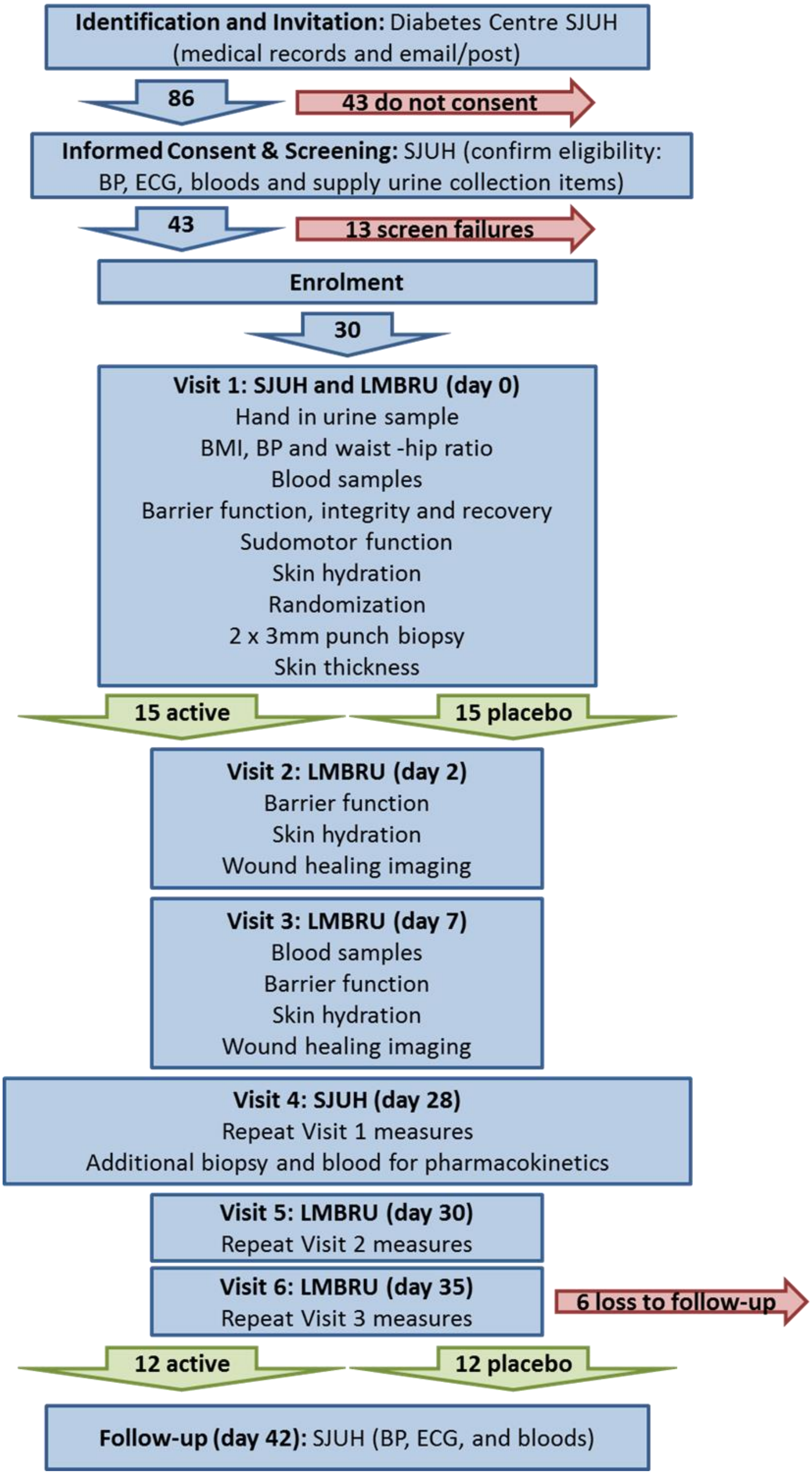

**Confidential**. Intellectual property: University of Leeds.

### 1. INTRODUCTION

#### 1.1. Background

Chronic, non-healing wounds e.g. diabetic foot ulcers (DFU) are a common worldwide health problem that have substantial medical and socioeconomic importance and represent a major unmet clinical need [1]. In Europe, 1-1.5% of the population has a problem wound at any one time. The average cost per episode is 6,650€ for leg ulcers and 10,000€ for foot ulcers, accounting for 2-4% of the healthcare budget and likely to escalate with an increasingly elderly and diabetic population [2]. In the Leeds/Bradford region the overall prevalence of wounds is 2.8-3.6 people per 1000 population [3], up to 50% of which are chronically inflamed, non-healing wounds. Costs for wound care in the UK are estimated at £2.03-3.8 million per 100,000 population [4] and diabetes currently accounts for approximately 10% of the total health resource expenditure and is projected to account for around 17% in 2035/2036 [5].

The profound atrophogenic effects of glucocorticoids (GC) on human skin structure and function are well documented, causing decreased collagen content, increased transepidermal water loss (TEWL), dermal and epidermal thinning, telangiectasia, impaired wound healing (WH) and increased infection risk [6-13]. Keratinocytes, melanocytes, fibroblasts and sebocytes play significant roles as GC targets in these processes [11, 14]. These effects arise from GC excess including systemic [7, 8] and topical [10] GC therapy, Cushing's disease [6] and psychological stress [13, 15-17].

The expertise of our group has focused on 11β-hydroxysteroid dehydrogenase (11β-HSD) isozymes which regulate local GC availability in many tissues largely independently of circulating levels [18]. In skin, 11β-HSD1 activates cortisol from cortisone, is expressed in epidermal keratinocytes, hair follicles, sebaceous glands and dermal fibroblasts and is upregulated by GC in a forward-feedback manner [19, 20]. Conversely, 11β-HSD2 converts cortisone to cortisol and, in skin, is predominantly expressed in eccrine (sweat) glands where it functions to protect the mineralocorticoid receptor from inappropriate activation by GC (as with other mineralocorticoid-responsive tissues e.g. kidney) [21]. We recently demonstrated increased 11β-HSD1 activity during the inflammatory phase of mouse skin WH [22] and faster healing in 11β-HSD1-null mice treated with oral corticosterone (active GC in mouse) and topical application of carbenoxolone (CBX), an 11β-HSD non-selective inhibitor (unpublished observations). Strikingly, mice with global deletion of 11β-HSD1 were protected from age-induced dermal atrophy, with improved collagen processing and accelerated WH [19]. Others have also reported accelerated WH by 11β-HSD1 blockade in animal models of diabetes and GC excess [23, 24].

These findings suggest that 11β-HSD1 mediates the effects of circulating GC in skin and drives the cutaneous consequences of GC excess. However, **the role of 11β-HSD1 in regulating skin function in man remains unexplored**, despite evidence that 11β-HSD1 is

**Confidential**. Intellectual property: University of Leeds.

upregulated by pro-inflammatory cytokines e.g. IL-1β and TNF-α (abundant in chronic wounds) [25-27] and reports of increased systemic GC levels in patients with type 2 diabetes mellitus (T2DM) [28, 29]. Moreover, pro-inflammatory cytokines and GC synergistically increase 11β-HSD1 expression [30] which may exacerbate GC availability in to further impede WH.

Targeting 11β-HSD1 through selective inhibitors has been a focus of major pharma for the last 5 years. In 2011, 11β-HSD1 was cited as the second most popular therapeutic target for patents filed (www.thomsonreuters.com/content/science/pdf/ls/iyc2011.pdf), the main target indication being metabolic syndrome to reduce hepatic gluconeogenesis, steatosis and visceral adipogenesis. Proof of concept was established in a number of phase II studies but effect size may prevent progression to phase III [31-34]. Selective 11β-HSD1 inhibitors, such as AZD4017, are now being released for repurposing studies funded through schemes such as the MRC Asset Sharing Initiative. **The ability of systemic 11β-HSD1 inhibitors to target enzyme activity in peripheral tissue (e.g. skin) in man is unknown**.

Our proposed pilot trial will generate efficacy, safety and study feasibility data for the application of selective 11β-HSD1 inhibitors as novel therapies to improve skin function and WH in diabetes. Our skin-specific study variables are a combination of validated disease-related outcomes and **measures previously unexplored in T2DM** that will greatly improve our knowledge of GC metabolism in skin. The potential to accelerate WH in patients with T2DM would contribute to a significant **improvement in patient quality of life and reduction in costs of care**.

#### 1.2. Investigational medicinal product (IMP)

##### 1.2.1. Investigator brochure (IB) updates

Please refer to the current IB (edition 9 Oct 2016) for more information.

##### 1.2.2. Summary of product characteristics

Not applicable

##### 1.2.3. Non IMP(s)

A placebo tablet containing microcrystalline cellulose and sodium stearyl fumarate is also available to match the active tablets in size, shape and colour. Matched placebo tablets will be provided by AstraZeneca.

#### 1.3. Rationale for the proposed study

The project is well-aligned with the research strategies of the School of Medicine at the University of Leeds and compliments a key platform of the Leeds Institute of Rheumatic and Musculoskeletal Medicine: Regeneration and Repair. This preliminary study aims to investigate efficacy and safety of 11β-HSD1 inhibition on skin function in patients with

**Confidential**. Intellectual property: University of Leeds.

T2DM. Study feasibility will also be assessed; if successful, data from this pilot study will inform power calculations for a future trial to investigate the ability of 11β-HSD1 inhibition to promote foot ulcer healing in T2DM.

### 2. STUDY AIM AND OBJECTIVES

#### 2.1. Study aims

To evaluate efficacy of oral AZD4017 in skin, determine safety of AZD4017 in patients with T2DM, examine effects of AZD4017 on skin function and asses study feasibility.

#### 2.2. Primary objective

Evaluate AZD4017 efficacy

#### 2.3. Secondary objectives

1) Evaluate safety of AZD4017

2) Examine effects of AZD4017 on skin function

3) Assess study feasibility

### 3. STUDY ENDPOINTS

#### 3.1. Primary endpoint

24 hour 11β-HSD1 activity in skin (efficacy) at baseline and day 28

#### 3.2. Secondary endpoints

Systemic 11β-HSD1 activity at baseline and day 35 and AZD4017 quantification in skin at day 28 and plasma at day 35 (to support efficacy), safety variables at baseline and days 7, 28 and 35, blood pressure, blood safety variables, EGC and biopsy inspection at day 42, urinary cortisol to cortisone metabolite analysis at baseline and day 35 (to assess systemic GC levels), skin function variables at baseline and days 2, 7, 28, 30 and 35 and feasibility variables at throughout the study.

### 4. STUDY VARIABLES

#### 4.1. Primary variable

**<u>AZD4017 efficacy</u>**

- 24 hour 11β-HSD1 activity in skin

#### 4.2. Secondary variables

**AZD4017 efficacy**

- 24 hour urinary cortisol to cortisone metabolite ratio
- AZD4017 levels in skin and plasma

**Safety**

- Body mass index (BMI)
- Waist-hip ratio
- Blood pressure (BP)
- Blood HbA1c
- Blood lipids
- Full blood count (FBC)
- Liver function test (LFT)
- Estimated glomerular filtration rate (eGFR)
- Kidney function test (U+E)
- Adrenal function (testosterone and dehydroepiandrosterone sulphate)
- Thyroid function
- Adverse event (AE) reporting
- Number of patients discontinuing study therapy due to safety

**T2DM-related skin variables**

- Sudomotor function (in each hand or foot, average in hands, average in feet, overall average)
- Skin hydration

**Exploratory skin variables**

- Epidermal barrier function
- Epidermal barrier recovery
- Epidermal barrier integrity
- Skin thickness
- WH
- RNA-seq gene expression profiling

**Other secondary variables**

- 24 hour urinary free cortisol

**Feasibility**

- Data on eligibility (proportion of patients assessed for eligibility deemed to be eligible and for each inclusion/exclusion criterion, proportion of patients eligible)
- Data on recruitment success (recruitment rate per month and week by week)
- Data on consent success (proportion of eligible patients consented)

**Confidential**. Intellectual property: University of Leeds.

- Assessment of randomization (evenness of numbers randomised to the two groups and balance of baseline characteristics between the two groups)
- Data on adherence to intervention (number of patients completing diary card, percentage compliance throughout treatment phase and reasons for missed doses)
- Data on retention to trial (number of patients completing final study assessment, reasons for discontinuation and number discontinuing in each case)
- Data on completeness of outcome measure reporting (for each outcome, number of patients with data available at each visit)

### 5. STUDY DESIGN

#### 5.1. Study description

This study aims to conduct a double-blind, randomised, parallel group, placebo-controlled phase II pilot trial of 35 days′ duration with 400mg oral AZD4017 twice daily (n=15) or placebo (n=15) in patients with T2DM.

Eligible participants will be identified using medical records from the Diabetes Clinic at St. James' University Hospital (SJUH) by the CI or an authorised member of the direct care team. Once identified, patients will be invited to participate in the trial by email and/or post and provided with a Participant Information Leaflet (PIL) and PIL Feedback Form (returnable by self-addressed prepaid envelope).

Patients will be contacted by telephone to confirm willingness to participate and eligibility after 72 hours since sending the PIL.

#### 5.2. Study duration

The protocol treatment duration is 35 days with follow-up visit at least 7 days after the last treatment dose.

Skin, plasma, serum and 24 hour urine samples will be stored for future biochemical analyses but storage will not exceed 5 years after the end of the trial. At 5 years after the end of the trial any residual samples will either be destroyed or placed in an approved Research Tissue Bank.

#### 5.3. Rationale for study design and selection of dose

**AZD4017 efficacy**

The ability of 11β-HSD1 inhibitors to reduce 11β-HSD1 activity in peripheral tissues has not been determined in man. Our primary variable will therefore evaluate the ability of AZD4017 to inhibit 24 hour 11β-HSD1 activity in skin. In addition to assessment of efficacy, this outcome measure will be used to provide preliminary data on associations between 11β-HSD1 activity levels and skin function variables. To support this outcome measure,

**Confidential**. Intellectual property: University of Leeds.

secondary variables will include systemic AZD4017 efficacy by measuring urinary cortisol to cortisone metabolite ratios ([THF+alloTHF]/THE) and pharmacokinetic analyses to confirm AZD4017 levels in plasma and skin biopsies.

**Safety**

AZD4017 has been studied in healthy human volunteers and is tolerated when given in single doses up to 1500mg and in multiple doses up to 900mg for periods of up to 9 days. A Phase IIa study of 28 days duration has been conducted in patients with raised intraocular pressure; 7 subjects received 200 mg once daily AZD4017 for up to 28 days and 19 subjects received 400 mg twice daily AZD4017 for up to 28 days. The most commonly reported AE in the studies with AZD4017 were gastrointestinal disorders and headache and were of mild to moderate intensity. Previous safety studies in healthy volunteers also reported an increase in ACTH and dehydroepiandrosterone sulfate levels. However, cortisol and testosterone levels were not changed. As the proposed pilot trial is relatively short, we will measure dehydroepiandrosterone sulfate and testosterone. However, blood samples will be stored allowing ACTH and cortisol to be measured in any patient showing evidence of hypothalamic-pituitary-adrenal (HPA) axis activation.

Other safety laboratory analyses showed no clinically relevant changes. No clinically relevant changes or trends were seen in vital signs or in ECG results in previous safety studies.

Although other 11β-HSD1 inhibitors have been trialled in patients with T2DM and were safe and well-tolerated [32, 33], AZD4017 has not been tested in this patient cohort. This preliminary pilot trial will therefore evaluate safety by monitoring validated safety variables of T2DM (e.g. BMI, waist-hip ratio, BP, HbA1c and lipids) and AZD4017 (e.g. FBC, LFT, eGFR, U+E, adrenal and thyroid function). Further information regarding non-clinical data and previous clinical studies can be found in the AZD4017 IB.

Data collected will be used to inform powering and feasibility of a future confirmatory trial.

**Treatment duration**

The study design is suitable for the time and funding constraints available. The study duration (35 days) is sufficient to detect changes in all T2DM-related skin secondary outcome measures [35-39].

**Dose**

AZD4017 is a novel orally bioavailable small molecule inhibitor of 11β-HSD1 enzyme activity. It is potent and highly selective in vitro and in vivo. The half maximal inhibitory concentration (IC50) for inhibition of 11β-HSD1 activity (cortisone to cortisol conversion) is 2nM. AZD4017 is selective (> 2000x) for 11β-HSD1 over human recombinant 11β-HSD2 and the closely-homologous enzymes 17β-hydroxysteroid dehydrogenase 1 (17β-HSD1) and 17β-hydroxysteroid dehydrogenase 3 (17β-HSD3) *in vitro*.

**Confidential**. Intellectual property: University of Leeds.

The dose to be used in this study is 400mg twice daily. This dose results in a decrease in the urinary ratio of tetrahydrocortisol to tetrahydrocortisone metabolites ([THF+alloTHF]/THE) from 0.99+0.27 to 0.11+0.06 and decreases the generation of plasma prednisolone following oral prednisone by 80%. Pharmacokinetic and pharmacodynamic data from single and multiple ascending dose studies in healthy volunteers suggest that a dose of 400mg twice daily may achieve approximately 90% inhibition of 11β-HSD1 over 24 hours. The safety and tolerability profile of AZD4017 has been studied in animals and in single and multiple ascending dose studies in human volunteers and supports the use of AZD4017 twice daily to 900 mg/day. The 35-day treatment period is considered sufficient to see sustained inhibition of 11β-HSD1 by AZD4017, and therefore this dose and duration is appropriate. Please refer to IB for more information.

**Other secondary variables**

Additional secondary study variables have been selected to represent a combination of clinically validated disease-related and/or GC-regulated exploratory outcome measures of skin function. Peripheral diabetic neuropathy in diabetes is characterized by a decrease in intra-epidermal nerve fiber density and sudomotor function leading to decreased skin hydration which and an increased risk of DFU [40-47]. Sudomotor function analysis has superseded intra-epidermal nerve fiber density as the “gold standard” for evaluating peripheral diabetic neuropathy and measures sweat gland innervation as a direct indicator of cutaneous C-fiber function [48]. Although the regulation of sudomotor function by GC is poorly understood, skin hydration is known to be reduced by GC treatment [49] and will also be investigated as a secondary study variable. The proposed study will generate novel data on the regulation of DFU risk factors by 11β-HSD1-mediated GC activation in skin.

Previous studies have indicated increased TEWL following treatment with topical GC [50] and acute systemic stress [51]. Conversely, acute systemic stress (physical restraint) decreased TEWL in rodent models of inflammatory diseases [52]. Therefore, we anticipate that limiting local GC reactivation by 11β-HSD1 inhibition may regulate TEWL. Impaired barrier recovery has been reported following systemic GC excess in humans [53] and rodents [54-56]. The number of tapes required to induce a standardized degree of barrier disruption will be used to evaluate barrier integrity, which is also impaired by GC excess [55, 56]. Skin thinning is a frequently reported side-effect of GC excess driven by both dermal and epidermal thinning [8, 9, 12, 57, 58], however, regulation of skin thinning by 11β-HSD1 in man remains unexplored. Impaired WH is a common side-effect of GC excess [11, 59] and 11β-HSD1 has been implicated in mouse WH models by us [19, 22] and others [23, 24]. However, this remains to be investigated in man. Although chronic wounds are a widely reported feature of T2DM, acute WH has not been fully investigated. Moreover, TEWL, barrier recovery, epidermal integrity and skin thickness remain unexplored in T2DM. Our study will provide a new insight into the regulation of these key skin functions by 11β-HSD1 in T2DM. RNA-seq will also be used for transcriptomic profiling of gene expression to identify key pathways involved in 11β-HSD1-mediated regulation of skin function.

**Confidential**. Intellectual property: University of Leeds.

Systemic GC levels have been shown to be important in determining peripheral GC exposure by regulating 11β-HSD1 substrate (cortisone) availability [18]. Therefore, systemic GC levels in combination with peripheral 11β-HSD1 activity may affect AZD4017 responsiveness and will also be monitored by measuring 24 hour urinary free cortisol levels.

Study feasibility will be assessed using a range of quantifiable measures including eligibility, recruitment, consent, randomization, adherence, retention and data completeness as in accordance with Consolidated Standards of Reporting Trials guidance [60, 61].

### 6. SELECTION AND WITHDRAWAL OF SUBJECTS

#### 6.1. Target population

Adults aged 18 years or older, with T2DM

#### 6.2. Estimated number of eligible participants

Current data from the NIHR Yorkshire and Humber Clinical Research Network identified >45,000 patients with T2DM from 11 general practitioner (GP) practices, indicating a large target population pool. Therefore, we estimate a trial invitation requirement of 86 participants of whom an estimated 50% will consent and of whom an estimated 70% will be eligible (using the criteria below) to reach our target of 30 participants (allowing a 20% post-randomization drop-out rate to achieve a total of 24 completers with 12 in each arm).

#### 6.3. Eligibility criteria

##### 6.3.1. Inclusion criteria

1) Able and willing to provide informed consent

2) Clinical diagnosis of T2DM with HbA1c <11% (<97 mmol/mol) at screening while taking standard therapy at a stable dose for ≥10 weeks

##### 6.3.2. Exclusion criteria

1) Women of child-bearing potential (WOCBP). Note: WOCBP include any female who has experienced menarche and who has not undergone successful surgical sterilization (hysterectomy, bilateral tubal ligation, or bilateral oophorectomy) or is not postmenopausal [defined as amenorrhea >/= 12 consecutive months or women on hormone replacement therapy with documented serum follicle stimulating hormone level >35 mIU/mL].

Additionally, male study participants who are sexually active with a female partner of childbearing potential must be surgically sterilized or agree, along with their partner, to use a highly effective method of birth control (as defined below) from the time of screening until 3 weeks after final dose of study drug (5 drug elimination half-lives plus 2 weeks). Male

**Confidential**. Intellectual property: University of Leeds.

study participants must also **not** donate sperm from the time of screening until 3 weeks after final dose of study drug (5 drug elimination half-lives plus 2 weeks).

Highly effective methods of contraception are defined as combined (estrogen and progestogen containing) hormonal contraception associated with inhibition of ovulation (either oral, intravaginal or transdermal), progestogen-only hormonal contraception associated with inhibition of ovulation (either oral [such as Cerazette™], injectable or implantable), intrauterine device (IUD), intrauterine hormone-releasing system (IUS), bilateral tubal occlusion, vasectomized partner or true sexual abstinence: When this is in line with the preferred and usual lifestyle of the subject. [Periodic abstinence (e.g., calendar, ovulation, symptothermal, post-ovulation methods), declaration of abstinence for the duration of a trial, and withdrawal are not acceptable methods of contraception].

2) Active leg/foot ulceration

3) Acute electrocardiogram (ECG) anomalies deemed to be clinically relevant by the CI

4) Systemic hypertension (BP >150/90), on 3 successive measurements at the screening visit (patients with controlled hypertension can be included in the trial) with the first acceptable measurement recorded and either systolic or diastolic BP exceeding these values requiring re-measurement

5) Any endocrine disorder (other than T2DM), including type 1 or secondary diabetes, except treated hypothyroidism with normal thyroid function tests for at least 3 months

6) Suspicion of or known Gilbert's disease

7) Alanine aminotransferase (ALT) and/or aspartate aminotransferase (AST) and/or alkaline phosphatase (ALP) > 1.5x upper limit of normal (ULN)

8) Bilirubin (total) > 1.5x ULN

9) An eGFR calculated by the Modification of Diet in Renal Disease equation of <45 ml/min/$m^2$

10) Creatine kinase (CK) >2 x ULN

11) Participant is, at the time of signing the informed consent, a user of recreational or illicit drugs (including marijuana) or has had a recent history (within the last year) of drug or alcohol abuse or dependence on questioning or clinical history

12) Receiving any topical, systemic (including vaginal/rectal) or inhaled GC treatment at the time of or within 3 months prior to the screening visit

13) Taking any anticoagulant medication (blood thinning e.g. warfarin)

14) Taking probenicid or similar (for gout) at the time of inquiry

15) Any medical/surgical procedure (excluding skin biopsies) or trauma during IMP administration or within one week following the last administration of the IMP as judged by the CI

**Confidential**. Intellectual property: University of Leeds.

16) Involved in the planning and/or conduct of the study (applies to both AstraZeneca staff and/or staff at the study site)

17) Participated in any other clinical study within 1 calendar month prior to the screening visit (except registry-only participation)

18) Deemed otherwise inappropriate to participate by the trial team

###### 6.3.3. Exclusions for general safety

Not applicable

###### 6.3.4. Screen failures

Participants who sign an Informed Consent Form and fail to meet the inclusion and/or exclusion criteria are defined as screen failures. For all screen failures, the investigator is to maintain a Screening Log that documents the screening number, participant initials, and reason(s) for screen failure. A copy of the log will be retained with the Case Report Form (CRF) and screening will be recorded as pass or fail in the CRF. Screening Log originals will be stored in the TMF.

##### 6.4. Recruitment, consent and randomization processes

###### 6.4.1. Recruitment

Eligible participants will be identified using medical records from the Diabetes Clinic at St. James's University Hospital by the Chief Investigator or an authorized member of the direct care team. Potentially eligible participants will be selected using a computerized search and review of medical records.

A PIL and PIL Feedback Form will be provided by a qualified member of staff who has signed/dated the staff delegation log for suitable candidates to consider. This will include detailed information about the rationale, design and personal implications of the study. Following information provision, patients will be contacted by telephone after 72 hours to ask whether they would be willing to take part in the trial and to check eligibility. This process will be clearly documented into the patient's medical notes.

###### 6.4.2. Consent

Assenting patients will then be invited to attend a screening visit where they will be formally assessed for eligibility and asked to provide written, informed consent. Where English is not the patient's first language every effort will be made to provide a Trust interpreter according to normal Trust procedures. The right of the patient to refuse consent without giving reasons will be respected. The original Informed Consent Form will be filed in the Trial Master File (TMF), with one copy given to the patient and one filed in the hospital notes. Consent pass or fail will also be recorded in the CRF. The written consent will be taken by an

**Confidential**. Intellectual property: University of Leeds.

eligible member of staff who has signed/dated the staff delegation log. The process of obtaining written consent will be clearly documented in the patient's medical notes.

A separate Informed Consent Form will be attained prior to the skin biopsy procedure which will be stored with the general Informed Consent Form in the TMF and recorded as pass or fail in the CRF.

##### 6.4.3. Randomization process

Treatment groups will be allocated on a fully randomised basis. A randomization schedule will be generated by the dedicated trials pharmacy representative who has signed/dated the staff delegation log and is not otherwise associated with this study. Patients will be randomised in a 1:1 treatment allocation ratio to either AZD4017 or placebo. The randomization schedule will be stored in a password protected file accessible only to the dedicated trials pharmacy (the CI and Scientific Lead will be blinded to the randomization process) until all samples have been collected.

##### 6.4.4. Definition for the end of the trial

The end of the trial will be defined as entry of the last participant's last data item.

#### 6.5. Withdrawal criteria

Participants are at any time free to withdraw from study (IMP and assessments), without prejudice to further treatment (withdrawal of consent) at any point. Such participants will always be asked about the reason(s) and the presence of any AE. Participants are not under obligation to give a reason for withdrawal. If possible, they will be invited to attend a follow-up visit, ideally, at least 7 days after stopping the IMP. They will undergo the follow-up visit safety measurements and be formally discharged by telephone. Any AE will be followed up wherever possible and all remaining IMP should be returned by the participant.

Should the participant need to be withdrawn from the active treatment phase of the study between study visits for safety reasons e.g. serious adverse event (SAE) or laboratory abnormalities, study medication will be stopped immediately. If withdrawn from IMP, participants will be free to complete the remaining protocol if they choose to do so unless deemed clinically unfit by the CI, in which case they will be withdrawn from the remainder of the protocol.

IMP withdrawal criteria are as follows:

1) Participant decision. The subject is at any time free to discontinue treatment, without prejudice to further treatment

2) The participant has a clinically significant or serious AE that would not be consistent with treatment continuation, as determined by the CI or the participant; including but not limited to hospitalisation or disability, requirement for medical or surgical intervention to prevent

**Confidential**. Intellectual property: University of Leeds.

hospitalisation or disability; or abnormalities of liver, muscle or thyroid function that are considered unacceptable by the CI

3) Hepatotoxicity: ALT and/or ALP and/or AST >2.5x ULN or bilirubin >2x ULN. This is in keeping with Common Terminology Criteria for AE (v5.0) Grade 2 recommendations from the National Institute of Health.

4) Renal toxicity: eGFR <45

###### 6.5.1. Patients who withdraw consent

Unless the participant specifically withdraws consent for their data and samples to be stored, all data and samples that have already been collected from them will continue to be stored as per the original participant consent. This will also apply to participants who are unable to confirm enduring consent due to loss of capacity.

###### 6.5.2. Managing/replacing patients who withdraw early

Participants who decide to withdraw will be invited to complete a withdrawal form which will be stored in the TMF (although refusal to do so will be respected). Participants who are withdrawn by the CI will be recorded in a Withdrawal Log that documents the enrolment number, participant initials, reason(s) for withdrawal and treatment offered (if applicable). A copy of the log will be retained in the CRF which will also directly capture information on withdrawal outcome (Discontinuation due to Adverse Event or Elective Withdrawal). Withdrawal Log originals will be stored in the TMF. Participants who withdraw or are withdrawn pre-randomization will be classified as screening failures (see section 6.3.4). Participants who withdraw or are withdrawn post-randomization (see section 6.5) will be classified as withdrawals and will not be replaced.

### 7. STUDY BLINDING

#### 7.1. Type of blinding

Treatment groups will be allocated in a double-blind manner. Participants will be blinded to the treatment they receive (placebo or IMP) throughout all stages of the study. Investigators will also be blinded to the treatment until all samples have been collected and processed. Blinding will be generated by the dedicated trials pharmacy representative who has signed/dated the staff delegation log and is not otherwise associated with this study.

#### 7.2. Procedure for production and maintenance of blind

Blind production and maintenance will be according to dedicated trials pharmacy guidelines.

#### 7.3. Breaking the blind in an emergency

**Confidential**. Intellectual property: University of Leeds.

In an emergency (e.g. participant AE) the blind may be broken to determine whether this may be treatment-related. Blinding for previously collected or future treatments will not be compromised.

### 8. STUDY TREATMENTS

#### 8.1. General information on the products (trial drugs) to be used

| IMP | Dosage form and strength | Manufacturer |
|---|---|---|
| AZD4017 | Tablets 200 mg | AstraZeneca |
| Placebo to match AZD4017 | Tablets | AstraZeneca |

Please refer to the IB for more information.

#### 8.2. Frequency and duration of the trial drugs

Participants will be randomised to receive either AZD4017 400mg twice daily (as close to 12 hours apart as possible) or placebo. AZD4017 or placebo will be dosed as 2 tablets orally twice daily for 35 days starting at the baseline visit (Visit 1).

#### 8.3. Administration/handling of the trial drugs

The AZD4017 tablets and matching placebo will be supplied to the dedicated trial pharmacy by Almac in bottles of 32, 200mg tablets with a unique kit number (sufficient for 8 days per bottle); 4 bottles will be dispensed at Visit 1 and 1 bottle will be dispensed at Visit 4 (sufficient for 5 days overage per participant). Almac will provide the pharmacy with a kit list to refer to when dispensing for each patient and a set of code breaks for emergency unblinding.

Participants will be instructed to take the AZD4017 or placebo tablets 30min before or at least 2 hours after a meal. In the case of vomiting or diarrhoea after taking the tablets, participants will be instructed not to repeat the dose but to wait for their next scheduled dose.

##### 8.3.1. Handling, storage and supply

Labels will be prepared in accordance with Good Manufacturing Practice (GMP) and local regulatory guidelines. The labels will fulfil GMP Annex 13 requirements for labelling. Label text will be in English. The labels will have a peel-off portion, which will be inserted into the participant's source data verification document at dispensing.

All study medications will be kept in a secure place under appropriate storage conditions. The IMP label on the pack specifies the appropriate storage.

##### 8.3.2. Drug accountability

**Confidential**. Intellectual property: University of Leeds.

The IMP provided for this study will be used only as directed in the study protocol. The study personnel will account for all IMP dispensed to and returned from the participants.

AstraZeneca personnel or its representative CTRU will ensure accountability logs are maintained for all study drugs received at the site, unused study drugs and for appropriate destruction. If possible, the study drug will be destroyed locally by the site pharmacy. Certificates of delivery and destruction will be signed and stored in the pharmacy site file.

#### 8.4. Prior and concomitant illnesses

Participants and their GP will be requested to report any prior or concomitant illness and these will be assessed on an individual basis as to the most appropriate subsequent actions to be taken (if any) based on the exclusion and withdrawal criteria described above.

We do not anticipate that ADZ4017 will adversely affect any prior or concomitant illness.

#### 8.5. Prior and concomitant medications

We do not anticipate that AZD4017 will adversely affect any prior or concomitant medications; however, certain medications are prohibited as they may interfere with study outcome measures.

##### 8.5.1. Permitted prior medications

Participants will be allowed to receive any treatment that is judged to be in their best medical interests. Participants and their GP will be requested to report any new treatment and participants may be withdrawn according to exclusion and withdrawal criteria.

##### 8.5.2. Prohibited prior medications

No specific drug interactions have been reported for AZD4017. However, the following medications are listed in the exclusion criteria as they may interfere with data integrity:

- Any topical, systemic (including vaginal/rectal) or inhaled GC treatment at the time of or within 3 months prior to the screening visit as this may affect 11β-HSD1 activity and secondary outcome measures
- Any anticoagulant medication (blood thinning e.g. warfarin) as this may reduce blood clotting and delay WH
- Probenicid or similar (for gout) as this may be increase AZD4017 excretion

##### 8.5.3. Permitted concomitant medications

Participants will be allowed to receive any treatment that is judged to be in their best medical interests.

##### 8.5.4. Prohibited concomitant medications

Please refer to exclusion criteria.

**Confidential**. Intellectual property: University of Leeds.

##### 8.5.5. Surgical procedures

We do not anticipate that AZD4017 will adversely affect any prior or concomitant surgical procedures but as a precaution patients will be ineligible to participate in the event of any medical/surgical procedure (excluding skin biopsy) or trauma during IMP administration or within one week following the last administration of the IMP as judged by the CI.

#### 8.6. Special warnings and precaution for use

AZD4017 has been studied in healthy human volunteers some patient groups and is generally well tolerated.

The most commonly reported AE amongst healthy volunteers were gastrointestinal disorders and headache and were of mild to moderate intensity. Please refer to IB for more information.

In a multiple ascending dose study (see IB for details) 30 healthy male subjects received single doses of AZD4017 followed by repeated doses of 75 mg once daily up to 900 mg twice daily for 9 days. The expected inhibition on 11β-HSD1 enzyme activity in liver measured by prednisolone generation in plasma after an oral prednisone challenge as well as on urinary GC metabolites was observed. An activation of the HPA axis was also indicated as an increase in both adrenocorticotropic hormone (ACTH) and dehydroepiandrosterone sulphate levels while cortisol and testosterone levels were unchanged. A few subjects had transient increases in ALT and AST at 1-1.5xULN. One subject taking 1200mg once daily experienced an increase in ALT to 3x ULN. In all cases, liver transaminases returned to baseline on withdrawal of study drug. There is **no clear relationship with plasma exposure as the subjects with the highest ALT did not have the highest plasma area under the curve of either AZD4017** or the glucuronic acid metabolite.

In a Phase IIa trial of subjects with raised intraocular pressure, 7 subjects receiving 200mg AZD4017 once daily and 19 subjects receiving 400mg AZD4017 twice daily for up to 28 days, **no deaths or SAE were reported**; in the group receiving AZD4017 400 mg twice daily three subjects had ALT between 1 and 1.5x ULN, four subjects had AST between 1 and 1.5x ULN, two subjects had gamma-glutamyl transferase between 1 and 1.5x ULN and one subject had GGT between 1.5 and 2x ULN. An activation of the HPA-axis was demonstrated by an increase in ACTH and dehydroepiandrosterone sulfate levels. However, cortisol and testosterone levels were unaffected. No AEs associated with these findings were reported. In the clinical studies performed so far no obvious muscle effects have been seen.

Non-clinical data and observations in previous clinical studies with similar and even higher repeated doses of AZD4017 support dose administration according to the proposed clinical study protocol. Further information regarding non-clinical data and previous clinical studies can be found in the AZD4017 IB.

Blood samples will be taken at screening, baseline, days 7, 28, 35 and follow-up to monitor HbA1c, lipids, liver, kidney, thyroid and adrenal function.

**Confidential**. Intellectual property: University of Leeds.

#### 8.7. Dose modifications

We do not anticipate any requirements for dose modifications.

#### 8.8. Assessing subject compliance with study treatment(s)

Participants will be asked to complete a daily Diary Card indicating the time of treatment (at approximately 12h intervals).

The administration of all study medication will be recorded directly into the CRF as the expected number of tablets remaining, the actual number of tablets remaining, the overall percentage compliance ((140-(actual number of doses remaining-expected number of tablets remaining))/140)*100 and the cumulative percentage compliance (1-((actual number of tablets remaining-expected number of tablets remaining) / (visit day x 4)))*100 at Visits 2-6. Any missed tablets will be documented in the medical notes, along with reasons for the missed tablets.

Percentage Diary Card completion will also be recorded in the CRF (number of doses recorded divided by the total available number of doses and multiplied by 100) along with a copy of the Diary Card. Diary Card originals will be stored in the TMF.

#### 8.9. Withdrawal of treatment

##### 8.9.1. Subject compliance

Participants will not be withdrawn on grounds of non-compliance in accordance with recent European Medicines Agency to avoid the presence of unobserved measurements as much as possible.

##### 8.9.2. Lack of efficacy

Efficacy will be evaluated after termination of recruitment and a lack of efficacy will therefore not affect participant withdrawal.

### 9. METHODS OF ASSESSMENT

#### 9.1. Assessment of primary variable (AZD4017 efficacy)

##### 11β-HSD1 activity in skin

To evaluate efficacy of oral AZD4017 on 24 hour 11β-HSD1 activity in skin, 3mm punch biopsies will be obtained at Visits 1 and 4 from lower outer forearm (midpoint between wrist and elbow) performed under local anaesthetic (e.g. lidocaine). This procedure will be conducted by authorised trial personnel according to the staff delegation log and does not require sutures. Administration of local anaesthetic may involve a momentary stinging sensation after which the procedure is pain-free. 11β-HSD1 activity assays will be conducted by the Scientific Lead within 8h as previously described [19]. To prevent accidental

**Confidential**. Intellectual property: University of Leeds.

unblinding, 11β-HSD1 activity samples will be stored frozen until the blind is broken at the end of the trial and will then be batch processed. Approximately 10% of the sample will be used for validation using a cortisol Enzyme Linked Immunosorbent Assay. Raw data (enrolment number, visit number and date) will be copied in its original form to secure server storage and recorded in the CRF as % conversion of cortisone to cortisol per hour.

#### 9.2. Assessment of secondary variables

##### 9.2.1. Efficacy

###### 9.2.1.1. Global 11β-HSD1 activity

Systemic 11β-HSD1 activity will be inferred from urinary [THF+alloTHF]/THE ratios. These will be measured in 24 hour urine samples at Visits 1 and 6 by liquid chromatography–mass spectrometry. Samples will be taken using commercially available collection containers, frozen in aliquots and stored at -80°C in the WTBB designated storage area before batch shipping for analyses by the Institute of Metabolism and Systems Research (University of Birmingham) at the end of the trial. The original report generated for each sample will be stored in the TMF and recorded in the CRF at the end of the trial.

Measurements will be used to examine associations between participant systemic GC levels and outcome measures of skin function.

###### 9.2.1.2. AZD4017 in plasma and skin

Skin samples will be collected at Visit 4 and blood plasma at visit 6 and stored at -80°C in the WTBB designated storage area before batch shipping for AZD4017 pharmacokinetic analysis by York Bioanalytical Solutions Ltd after the end of the trial. Blood samples will be collected by authorised research nurse support. Shipping contents, collection and delivery will be recorded and stored in the TMF. The original report generated for each sample will be stored in the TMF and recorded in the CRF.

##### 9.2.2. Safety

AE-related participant withdrawals (i.e. number of Discontinuations due to Adverse Events) e.g. exceeding hepatic/renal stopping criteria, myocardial infarction or stroke, inpatient hospitalization or any other AE deemed by the clinical care team to require trial withdrawal will be recorded as a Discontinuation due to Adverse Event under withdrawal outcome in the CRF. The following safety measures will also be used to assess AZD4017 safety:

###### 9.2.2.1. Clinical evaluation of biopsy site

Following punch biopsies, participants will be requested to monitor the wound for any excessive adverse reaction (e.g. redness, rash, swelling or pain) and report any concerns to their GP. During Visits 2, 3, 4, 5 and 6, the application site will also be assessed by authorised trial personnel for signs of infection, detailed in the medical notes and recorded directly into the CRF as pass or fail.

**Confidential**. Intellectual property: University of Leeds.

###### 9.2.2.2. Body mass index

BMI is an attempt to quantify the amount of tissue mass (muscle, fat, and bone) in an individual, and then categorize that person as underweight, normal weight, overweight, or obese based on that value. BMI is defined as the body weight divided by the square of the body height, and is universally expressed in units of $kg/m^2$, resulting from mass in kilograms and height in metres. Height will be measured in metres to the nearest centimetre (e.g. 1.63m) and weight will be measured in kilograms to the nearest 100 grams (e.g. 60.2kg). Split values will be rounded up (e.g. 1.625m to 1.63m and 60.15kg to 60.2kg). BMI will be calculated using the formula: BMI = weight (kg)/height ($m^2$) and rounded up to two decimal place (e.g. BMI = $60.2/1.63^2$ = 22.66). Commonly accepted BMI ranges are underweight: under 18.5 $kg/m^2$, normal weight: 18.5 to 25, overweight: 25 to 30, obese: over 30. Measurements will be conducted at Visits 1 and 6 by a qualified member of staff who has signed/dated the staff delegation log and weight, height and BMI will be recorded directly into the CRF.

###### 9.2.2.3. Waist-hip ratio

Waist-hip ratio is used in combination with BMI to assess obesity and can be more accurate as BMI can be skewed by muscle mass [62]. The waist-hip ratio is the ratio of the circumference of the waist to that of the hips. This is calculated as waist measurement (to the nearest cm) divided by hip measurement (to the nearest cm). Measurements will be conducted at Visits 1 and 6 by a qualified member of staff who has signed/dated the staff delegation log and waist circumference, hip circumference and waist-hip ratio will be recorded directly into the CRF.

###### 9.2.2.4. Blood pressure

Blood pressure is usually expressed in terms of the systolic (maximum) pressure over diastolic (minimum) pressure and is measured in millimeters of mercury (mm Hg) via a non-invasive sphygmomanometer. Measurements will be conducted at screening, Visits 1, 6 and follow-up by a qualified member of staff who has signed/dated the staff delegation log and recorded directly into the CRF.

###### 9.2.2.5. Blood tests

Blood samples collected at screening, baseline, study Visits 3 (day 7), 4 (day 28), 6 (day 35) and follow-up (day 42 onwards) will be analysed immediately through the Leeds Teaching Hospitals NHS Trust Pathology Service or processed and stored for future research.

Blood samples at each of these visits will evaluate HbA1c (mmol/mol) and lipids [total and high density lipoprotein cholesterol and triglycerides (mmol/l)] for T2DM disease markers and FBC [haemoglobin (g/l), white cells and platelets ($x10^9/l$), red cells ($x10^{12}/l$), mean corpuscular volume (fl), haematocrit (%/100), mean corpuscular haemoglobin (pg), mean corpuscular hemoglobin concentration (g/l) and red blood cell distribution width (%)], LFT

**Confidential**. Intellectual property: University of Leeds.

[ALT, AST and gamma-glutamyl transpeptidase (iu/l), ALP (U/l), albumin (g/l) and bilirubin (umol/l)], eGFR (ml/min/1.73m$^2$), U+E [sodium, potassium and urea (mmol/l) and creatinine (umol/l)], adrenal [testosterone (nmol/l) and dehydroepiandrosterone sulphate (umol/l)] and thyroid [thyroid stimulating hormone (mIU/l) and free thyroxine (pmol/l)] function (AZD4017 safety markers). CK (iu/l) will also be measured at the screening visit. Samples will be collected by authorised research nurses.

Results will be sent to the CI or authorised delegated personnel for immediate monitoring signature and dating. Signed and dated result reports will be stored with the patient's medical notes for each visit and recorded in the CRF as pass or fail.

Blood will also be taken at baseline and day 35 for separation to serum and plasma and stored at -80°C in the WTBB designated storage area for plasma AZD4017 detection and future studies (up to 5 years). At 5 years after the end of the trial any residual samples will either be destroyed or placed in an approved Research Tissue Bank.

###### 9.2.2.6. Adverse event reporting

Any reported AE will record the following information directly into the CRF at Visits 2, 3, 4, 5 and 6

- Type of AE defined by the Common Terminology Criteria for Adverse Events (CTCAE) v5.0 preferred term
- Date of onset
- Severity (defined by CTCAE v5.0)
- Relation to study intervention (definite, probable, possible, unlikely, unrelated)
- Date of resolution

##### 9.2.3. Skin function

###### 9.2.3.1. Sudomotor function

The test is conducted using a Sudoscan device consisting of a simple desktop computer connected to two sets of large surface stainless steel electrodes: two for application of the palms, and two for the soles. The test will be conducted at Visits 1 and 6 and performed by the Scientific Lead at the SJUH Diabetes Centre. The procedure is non-invasive, pain-free and requires approximately 10min.

The patient places both hands and feet simultaneously on the designated electrodes and a painless scanning process ensues over the course of 2–3 min. A low voltage (1-4V) is incrementally applied to the electrodes, with the left and right electrodes serving alternatively as cathode and anode. At voltages <10V, the *stratum corneum* is electrically insulating; the sweat glands, however, consist of a cellular bilayer and therefore can transmit electrically charged ions to the electrodes through the skin's surface. The current of chloride ions generated is quantified and reflects C-fiber innervation. This chloride ion current is reported as electrochemical skin conductance measured in microSiemens (µS)

**Confidential**. Intellectual property: University of Leeds.

[48]. The results will be recorded directly into the CRF. Raw data on the Sudoscan device (including enrolment number, visit number and date) will be copied in its original form to secure server storage.

###### 9.2.3.2. Skin hydration

*Stratum corneum* hydration will be measured using a Corneometer CM 825 device. The device measures the change in the dielectric constant due to skin surface hydration changing the capacitance of a precision capacitor and can detect even slight changes in hydration (reported in arbitrary units).

The measurement will be taken at Visits 1 and 6 with a small portable probe that is applied to the skin, is non-invasive and pain-free. The test requires approximately 5min and will be conducted by the Scientific Lead. The results will be recorded in the CRF and stored as image files in a temporary folder (including enrolment number, visit number and date) on the secure (encrypted) laptop used for the measurements before transfer to a secure server.

###### 9.2.3.3. Epidermal barrier function

TEWL is a validated measure of skin epidermal permeability barrier function.

Evaporation of water from the skin occurs as part of normal skin metabolism. As barrier function is disrupted, water loss increases. The Tewameter TM 300 probe measures the density gradient of the water evaporation from the skin by two pairs of sensors (temperature and relative humidity) inside a hollow cylinder. The open chamber measurement method is the only method to assess the TEWL continuously without influencing its microenvironment. A microprocessor analyses the values and expresses the evaporation rate in $g/h/m^2$.

The procedure requires 5min to perform, is non-invasive, pain-free and will be conducted by the Scientific Lead at Visits 1, 2, 3, 4, 5 and 6. The results will be recorded in the CRF and stored as image files in a temporary folder (including enrolment number, visit number and date) on the secure (encrypted) laptop used for the measurements before transfer to a secure server.

###### 9.2.3.4. Epidermal barrier recovery

In addition to resting TEWL, epidermal function can be assessed by measuring TEWL recovery over time following barrier disruption by repeated tape stripping.

Following the initial TEWL measurement, adhesive patches (D-Squame tapes) will be used to gently remove the *stratum corneum* layers to a pre-specified TEWL rate from the lower inner forearm at Visits 1 and 4. Recovery of TEWL at this location will be evaluated by repeated TEWL readings at 3 hour, 48 hour and 7 days post-disruption at Visits 1, 2, 3, 4, 5 and 6.

**Confidential**. Intellectual property: University of Leeds.

This procedure will be conducted by the Scientific Lead and may cause a temporary mild discomfort or transient irritation which should subside within a few hours. The results will be recorded in the CRF and stored as image files in a temporary folder (including enrolment number, visit number and date) on the secure (encrypted) laptop used for the measurements before transfer to a secure server.

###### 9.2.3.5. Epidermal barrier integrity

The number of tapes required is an indication of *stratum corneum* integrity. Tapes will be stored at -80°C in the WTBB designated storage area for future molecular biology studies. The results will be recorded directly in the CRF.

###### 9.2.3.6. Skin thickness

Epidermal thickness will be evaluated by OCT at Visits 1 and 6 according to the manufacturer's instructions. Image files (including enrolment number, visit number and date) will be stored on the OCT machine until the end of the trial. They will then be transferred to a secure server, compiled, analysed and values for skin thickness entered into the CRF.

###### 9.2.3.7. Wound healing

Both biopsies from visit 1 and two biopsies from visit 3 will be imaged by OCT at Visits 2, 3, 5 and 6. Recent studies suggest OCT may be a useful tool to evaluate wound re-epithelialization [63]. This imaging modality is already being utilised by our department [64]. OCT uses light to capture sub-micrometer resolution, three-dimensional images from within biological tissue (e.g. skin). The method is based on low-coherence interferometry, typically employing near-infrared light. The use of relatively long wavelength light allows it to penetrate 1-2mm into the tissue. In addition to wound diameter, collected images capture information on vascularization which may be used for future studies.

The procedure takes approximately 2min using a small probe applied to the skin, is non-invasive, pain-free and will be conducted by the Scientific Lead or delegated authorised personnel. Image files (including enrolment number, visit number and date) will be stored on the OCT machine until the end of the trial. They will then be transferred to a secure server, compiled, analysed and values for wound diameter entered into the CRF.

###### 9.2.3.8. RNA-seq gene expression profiling

Freshly snap-frozen 3mm skin biopsies collected at Visit 1 and 4 will be used to examine differences in mRNA expression by RNA-seq between baseline measurement and following 35 days of AZD4017. Skin samples will be stored -80°C in the WTBB designated storage area until all samples have been collected prior to processing for RNA extraction and RNA-seq. Lists of expressed and differentially expressed genes will be stored on a secure sever (including enrolment number and visit number). Genes and pathways found to be regulated

**Confidential**. Intellectual property: University of Leeds.

by AZD4017 will be quantified by qPCR and the data (including enrolment number and visit number) will be stored on a secure server.

##### 9.2.4. Feasibility

Data for powering of secondary variables and feasibility of a future confirmatory trial will be captured on the CRF. Data on eligibility, recruitment, consent, randomization, adherence, retention and data completeness will be captured though a range of sources including eligibility, enrolment, screening and withdrawal logs, Informed Consent Forms, pharmacy records and diary cards (stored in the TMF) to enable assessment of future confirmatory trial feasibility.

### 10. STUDY PROCEDURES BY VISIT

#### 10.1. Summary schedule of study assessments

Table 1. Summary schedule of study assessments

| **Assessment (day)** | **Screening (-7 to -2)** | **Visit 1 (0)** | **Visit 2 (2)** | **Visit 3 (7)** | **Visit 4 (28)** | **Visit 5 (30)** | **Visit 6 (35)** | **Follow-up (>42)** |
|---|---|---|---|---|---|---|---|---|
| Eligibility | X | | | | | | | |
| Informed consent* | X | X | | | X | | | |
| ECG | X | | | | | | | X |
| BMI, BP, waist-hip ratio | BP only | X | | | | | X | BP only |
| Blood tests** | X | X | | X | X | | X | X |
| 24 hour urine collection | | X | | | | | X | |
| Skin hydration | | X | | | | | X | |
| Barrier function | | X | X | X | X | X | X | |
| Barrier recovery | | X | X | X | X | X | X | |
| Skin integrity | | X | | | X | | | |
| Sudomotor test | | X | | | | | X | |
| Biobank serum and plasma | | X | | | | | X | |
| Randomization | | X | | | | | | |
| Tablet supply | | X | | | X | | | |
| 11β-HSD1 activity (biopsy) | | X | | | X | | | |
| RNA-seq | | X | | | X | | | |

**Confidential**. Intellectual property: University of Leeds.

| | | | | | | | | |
|---|---|---|---|---|---|---|---|---|
| (biopsy) | | | | | | | | |
| Skin thickness | | X | | | | | X | |
| AE monitoring | | | X | X | X | X | X | X |
| Evaluation of biopsy site | | | X | X | X | X | X | X |
| Wound healing | | | X | X | | X | X | |
| Compliance monitoring | | | X | X | X | X | X | |
| AZD4017 in plasma and skin (biopsy)*** | | | | | | | X | |

* Informed consent will also be obtained prior to each biopsy

** HbA1c, lipids, FBC, LFT, eGFR, U+E, adrenal and thyroid function (CK will also be analysed at screening). For further detail see section 9.2.2.5

*** Active (AZD4017) arm only after unblinding at the end of the trial

#### 10.2. Screening visit (day -7 to -2) SJUH Diabetes Clinic

Patients invited to participate in the trial will be contacted by telephone after 72 hours and asked to confirm willingness to enrol, eligibility and identify a suitable study schedule. Two to seven days before the baseline visit (Visit 1), eligibility will be confirmed at screening by a medically qualified doctor and will be recorded in both the medical notes and CRF. Written informed consent will be obtained before testing for other exclusion criteria (clinically relevant acute ECG anomalies, hypertension and blood analyses). Potential participants will also be supplied with 24 hour urine collection devices. If all screening criteria are met, potential participants will be contacted by telephone when the results are ready to confirm enrolment and arrange a visit schedule.

#### 10.3. Visit 1 (day 0) SJUH Diabetes Clinic and CAH LMBRU

Participants will return 24 hour urine samples for storage until processing. Participants will be asked if anything has changed since the previous visit.

**Baseline measures** will be conducted as follows: 1) BMI, BP, waist-hip ratio 2) skin hydration 3) epidermal barrier function and integrity 4) sudomotor function 5) blood samples for HbA1c, lipids, FBC, LFT, eGFR, U+E, thyroid and adrenal function and serum/plasma biobank storage 6) Randomization to 400mg oral AZD4017 twice daily or placebo 7) collection of IMP from pharmacy 8) obtain biopsy informed consent 9) 2 x 3mm biopsies for 11β-HSD1 activity RNA-seq 10) 3 hour barrier recovery time-point 11) skin thickness 12) payment

Pre-paid transport between SJUH and CAH for the skin thickness measurement will be arranged by a member of the trial team.

#### 10.4. Visit 2 (day 2) at CAH LMBRU

We will monitor the biopsy site and participants will be asked if anything has changed since the previous visit and if they have experienced any AE. Measures will be conducted as follows: 1) epidermal barrier function 2) 48 hour barrier recovery time-point 3) biopsy site inspection and wound imaging by optical coherence tomography (OCT) 4) compliance monitoring 5) payment

#### 10.5. Visit 3 (day 7) at CAH LMBRU

We will monitor the biopsy site and participants will be asked if anything has changed since the previous visit and if they have experiences any AE. Measures will be conducted as follows: 1) epidermal barrier function 2) 7 day barrier recovery time-point 3) blood samples for HbA1c, lipids, FBC, LFT, eGFR, U+E, thyroid and adrenal function 4) biopsy site inspection and wound imaging by OCT 5) compliance monitoring 6) payment

#### 10.6. Visit 4 (day 28) at SJUH Diabetes Clinic

We will monitor the biopsy site and participants will be asked if anything has changed since the previous visit and if they have experiences any AE. Biopsy sites will be inspected and measures will be conducted as follows 1) epidermal barrier function and integrity 2) blood samples for HbA1c, lipids, FBC, LFT, eGFR, U+E, thyroid and adrenal function 3) collection of IMP from pharmacy 4) obtain biopsy informed consent 5) 3 x 3mm biopsies for 11β-HSD1 activity, RNA-seq and AZD4017 quantification 6) 3 hour barrier recovery time-point 7) compliance monitoring 8) payment

#### 10.7. Visit 5 (day 30) at CAH LMBRU

We will monitor the biopsy site and participants will be asked if anything has changed since the previous visit and if they have experiences any AE. Measures will be conducted as follows: 1) epidermal barrier function 2) 48 hour barrier recovery time-point 3) biopsy site inspection and wound imaging by OCT 4) participants will be supplied with 24 hour urine collection devices 5) compliance monitoring 6) payment

#### 10.8. Visit 6 (day 35) at CAH LMBRU

We will monitor the biopsy site and participants will return 24 hour urine samples for storage until processing. Participants will be asked if anything has changed since the previous visit and if they have experiences any AE. Measures will be conducted as follows: 1) BMI, BP, waist-hip ratio 2) skin hydration 3) epidermal barrier function 4) 7 day barrier recovery time-point 5) sudomotor function 6) blood samples for HbA1c, lipids, FBC, LFT, U+E, thyroid and adrenal function, serum/plasma biobank storage and AZD4017 quantification 7) remaining tablets and diary cards will be returned to the pharmacy 8) wound imaging by OCT 9) skin thickness 10) payment

**Confidential**. Intellectual property: University of Leeds.

Blood tests results from all visits will be sent to the CI or authorised delegated personnel for immediate consultation, signature and dating. Signed and dated blood reports will be stored in the medical notes and recorded in the CRF as pass or fail.

#### 10.9. Follow-up visit (day 42 onwards) at SJUH Diabetes Clinic

The follow-up visit will take place at least 7 days following cessation of trial treatments (day 42 onwards). At the follow-up visits, all biopsy sites will undergo clinical evaluation for signs of infection. ECG, BP and AE will be recorded. Blood samples will be collected for HbA1c, lipids, FBC, LFT, U+E, thyroid and adrenal function. Measurements will be conducted by a qualified member of staff who has signed/dated the staff delegation log and recorded directly into the CRF (ECG will be recorded in the CRF as pass or fail with original reports stored in the medical notes). Participants will be contacted by telephone to confirm follow-up findings and to be discharged from the trial (this will be recorded in the CRF). Dates of screening visits and Visits 1-6 will also be recorded directly into the CRF.

#### 10.10. Unscheduled visits

Presently, there is no information regarding overdose of AZD4017 in man and there is no known antidote for AZD4017. If an overdose is suspected any ongoing administration of AZD4017 should be stopped and the subject should be monitored closely and treated symptomatically. To determine the level of AZD4017 a blood sample must be drawn as soon as possible in proximity to the event. In addition, the time and extent of overdose must be ascertained. Since activation of the HPA-axis is a suspected pharmacological effect of excessive levels of AZD4017, blood sampling for testosterone and dehydroepiandrosterone sulfate should be carried out.

Test findings will be recorded in the CRF and participants may be withdrawn from the study.

### 11. PHARMACOVIGILANCE

#### 11.1. Defining adverse events

An AE is any untoward medical occurrence in a patient during or following administration of an investigational product and which does not necessarily have a causal relationship with treatment. An AE can therefore be any unfavourable and unintended sign (including an abnormal laboratory finding), symptom, or disease temporarily associated with the use of the trial drugs, whether or not considered related to the trial drugs.

For the purposes of this study, an AE can include an undesirable medical condition occurring at any time between the Baseline and Completion Visits. As this is a blinded trial, AE will be assessed for expectedness and causal relationship assuming that the participant has been receiving AZD4017.

**Confidential**. Intellectual property: University of Leeds.

#### 11.2. Defining serious adverse events

A SAE is an AE which is defined as serious, i.e. that it:

- Results in death. Death may occur as a result of the basic disease process. Nevertheless, all deaths occurring within 7 days of the last administration of the study agent must be treated as SAEs and reported as such. All deaths which may be considered as related to the trial agent, regardless of the interval, must be treated as a SAE and reported as such
- Is life-threatening
- Requires inpatient (overnight) hospitalization or prolongation of an existing hospitalization
- Results in a persistent or significant disability or incapacity
- Additionally, important medical events that may not result in death, be life-threatening, or require hospitalization may be considered SAE when, based on appropriate medical judgment, they may jeopardize the subject and may require medical or surgical intervention to prevent one of the outcomes listed in this definition
- Any other significant clinical event, not falling into any of the criteria above, but which in the opinion of the CI requires reporting

**Other Reportable Information:** Certain information, although not considered an SAE, must be recorded, reported, and followed up as indicated for an SAE. This includes the following:

- Overdose of IMP as specified in this protocol, with or without an AE
- Inadvertent or accidental exposure to IMP, with or without an AE

#### 11.3. AE of special interest

##### 11.3.1. Pregnancy

WOCP are excluded from this study. Should a pregnancy still occur, study medication will be discontinued immediately and the pregnancy reported to the University of Leeds and to AstraZeneca.

Pregnancy is considered a form of SAE. If a pregnancy is confirmed, use of the IMP must be discontinued immediately. Both maternal and paternal exposures are considered other reportable information. For exposure involving the female partner of a male subject, the necessary information must be collected from the subject, while respecting the confidentiality of the partner. All pregnancies will be followed up until birth.

#### 11.4. Defining suspected unexpected serious adverse reactions

A Suspected Unexpected Serious Adverse Reaction (SUSAR) is a SAE suspected to have a reasonable causal relationship to the IMP where the nature or severity of the reaction is inconsistent with the available product information (mainly referring to the IB). All SAE

**Confidential**. Intellectual property: University of Leeds.

assigned by the CI or delegated clinician as both *suspected* to be related to the trial drugs and *unexpected* are subject to expedited reporting.

#### 11.5. Exemptions from safety reporting

Not applicable

##### 11.5.1. Efficacy endpoints and disease progression events

Not applicable

##### 11.5.2. Other expected events

Not applicable

#### 11.6. Recording and reporting of AE

##### 11.6.1. Recording and reporting of all AE

Determination of AE will be based on the signs or symptoms detected during the physical examination and on clinical evaluation of the subject, and will be assessed and recorded at every visit. Signs and symptoms must be recorded using standard medical terminology. Subjects considered incapable of giving consent would not be considered for this study.

AE and SAE will be monitored from the first dose of protocol treatment to 7 days after the last dose of treatment with a protocol IMP. The Scientific Lead must instruct the subject to report AE and SAE during this time period.

During the time period specified above, the Scientific Lead will:

- Record all AE and SAE on source documents
- Record all AE and SAE in the CRF for subjects who are not screen failures
- Report all SAE on a 'CTT21 Serious Adverse Event Form'. Instructions on where to send this form will be provided by the Sponsor

The Scientific Lead must follow up on all AE and SAE until the events have subsided, returned to baseline, or, in case of permanent impairment, until the condition has stabilized. The Sponsor will maintain detailed records of all AE and SAE reported by the Scientific Lead in accordance with Good Clinical Practice (GCP – section 14.1) and applicable local regulations.

##### 11.6.2. Sponsor SAE and SUSAR reporting requirements

All SAE assigned by the Scientific Lead as both suspected to be related to protocol treatment and unexpected will be reviewed by the CI. The CI or other qualified and delegated individual may declare a SAE a SUSAR. In the absence of the CI the event will be reported as a SUSAR. If upon receipt of follow-up information the causality relation is changed, this information will be submitted to the MHRA as part of the SUSAR follow-up reporting

**Confidential**. Intellectual property: University of Leeds.

procedures. All Investigators will refer to the IB when determining whether a SAE is expected.

All SAE, other information reportable as SAE and follow-up information must be reported to the Sponsor within 24 hours of the research team becoming aware of them, by emailing a completed ‘CTT21 Serious Adverse Event Form’ to the email address. The Sponsor will confirm, by phone or e-mail, that the email was received. SUSAR are subject to expedited reporting to the Research Ethics Committee (REC) and Medicines and Healthcare products Regulatory Agency (MHRA).

Identifiable patient data, other than linked anonymised data required by the SAE form, must not be included when reporting SAE and SUSAR.

The Sponsor[1] then will inform the MHRA[2,3] via the MHRA eSUSAR web portal and the Main REC[2,3] of SUSAR within the required expedited reporting timescales.

2. SUSAR must be reported to the REC/MHRA within 7 calendar days of the CI (or their research team) being informed of the event, if they result in Death or are deemed to be life-threatening. Follow-up information must be reported within 8 calendar days
3. Any SUSAR not resulting in death or deemed to be life-threatening must be reported to the REC/MHRA within 15 calendar days of the CI (or their research team) being informed of the event. Follow-up information must be reported within 8 calendar days

SUSAR will be reported in accordance with the requirements and provisions of the applicable national laws. They will all be signed off by the CI or, in their absence, by a delegated individual.

##### 11.6.3. AstraZeneca SAE and SUSAR reporting requirements

The Sponsor will:

• Report unblinded SUSAR to AstraZeneca as individual case reports as they occur

• Report blinded listings of SAE and Suspected Serious Adverse Reactions to AstraZeneca on a quarterly basis

• Inform AstraZeneca within 24 hours of knowledge of the event of any emerging safety data or actions that the Sponsor is considering as a result of a safety signal with the IMP. This includes but is not limited to:

1. Urgent safety measures to be implemented in the study

2. Safety amendments to protocol/patient information and informed consent

**Confidential**. Intellectual property: University of Leeds.

3. Open reports from Independent Data Monitoring Committees excluding confidential reports to and meeting minutes

4. Interactions with Regulatory Authorities / Ethics Committees

5. Inform AstraZeneca on an ongoing basis of any new safety trends or signals observed during routine safety surveillance activities

• Report SUSAR / SAE through the AstraZeneca Contract Research Organization Kinapse

• Include the following essential information in SUSAR, Suspected Serious Adverse Reactions and SAE reports provided to AstraZeneca (initial and follow-up):

1. AstraZeneca Reference number

2. Sponsor trial number

3. Centre number

4. Patient trial number

5. Year of birth or age

6. Sex

7. IMP dose, start and stop date

8. AE onset and stop date

9. Event term as reported by the investigator (and/or the CTCAE v5.0 term and grade)

10. Investigator's assessment of seriousness (International Council for Harmonisation definitions)

11. Investigator's assessment of causality

12. SAE Outcome

#### 11.7. Urgent safety measures

If the research team becomes aware of information affecting the risk/benefit balance of the trial they may take immediate action to ensure patient safety. Urgent safety measures deemed necessary must be reported immediately by telephone to the MHRA (in conjunction with the Sponsor) and to the Main REC for the trial, and must be followed within three days by notice in writing setting out the reasons for the urgent safety measures and the plan for further action. The REC co-ordinator will acknowledge within 30 days.

#### 11.8. Serious breaches of protocol

A serious breach is a breach which is likely to effect to a significant degree either:

- The safety or physical or mental integrity of the subjects of the trial

**Confidential**. Intellectual property: University of Leeds.

- The scientific value of the trial

Serious breaches of GCP, the trial protocol and the Clinical Trial Authorisation will be reported to the Sponsor QA office within 24 hours from the time the research team becomes aware of the incident. A member of the research team must complete form 'CTT20: UoL/LTHT CTIMP Protocol Deviations, Violations and Potential GCP Breaches' and email it to.

#### 11.9. Other breaches of protocol

Although minor non-compliances from the protocol do not need to be reported to the MHRA as a serious breach, only the Sponsor QA office are permitted to make this assessment. A member of the research team must complete form 'CTT20: UoL/LTHT CTIMP Protocol Deviations, Violations and Potential GCP Breaches' and email it to within 24 hours of discovery.

If the research team have any doubt or uncertainty if what they have identified is a suspected serious breach, it must be discussed with the Sponsor QA office without delay, in accordance with Sponsor QA guidance and Standard Operating Procedure.

#### 11.10. Laboratory measurements

The laboratory staff will have access to a laboratory manual which will provide detailed descriptions of collection, preparation and labelling requirements for all laboratory samples for the study.

**Blood**

A maximum of 70 ml of blood will be collected at each study visit. The blood will be drawn into a combination of tubes, according to the planned experiments. After collection, blood samples will be analysed immediately or processed for serum and plasma and stored at -80°C in freezers at the WTBB SJUH for future biomarker and pharmacokinetic analyses, respectively.

Sample labels containing appropriate identification information will be provided.

The blood measurements to be collected in this study include T2DM-specific (HbA1c and lipids) and AZD4017-related (FBC, LFT, eGFR, U+E, thyroid and adrenal function) safety measures as presented in Table 2.

**Urine**

24 hour urine samples will be collected by the participant and brought in to appropriate study visits (see section 10). Samples will be aliquoted and stored at -80°C in the WTBB SJUH.

**Skin biopsies**

**Confidential**. Intellectual property: University of Leeds.

For 11β-HSD1 activity: sample will be placed in 1ml assay media and stored at room temperature until all participants for that day have been processed. Samples will be assayed overnight by incubating at 37°C with 200nM cortisone and ~1500cpm of [3H] cortisone. After 24 hours, tissue will be weighed and destroyed and media will be stored frozen. At the end of the trial, samples will be thawed to RT (an aliquot of 50µl will be retained for cortisol Enzyme Linked Immunosorbent Assay following manufacturer's recommendation) and steroids will be extracted from the media by vortexing with 3ml dichloromethane. The aqueous layer will be aspirated and the dichloromethane evaporated under air at 60°C. Dried steroids will be resuspended in 40µl dichloromethane, spotted onto aluminium-silica plates with 2µl 10mM cortisone/cortisol standards and separated by thin layer chromatography in a 186:14ml chloroform: ethanol mobile phase for 90min. Steroids are visualised under UV, marked, excised and extracted in scintillation fluid overnight. Following detection by scintillation, the % conversion of cortisol from cortisone is calculated and expressed as pmol/mg tissue/h. In the event of radioassay failure, Enzyme Linked Immunosorbent Assay data will be used for the calculation.

For RNA-seq: sample will be snap-frozen and stored at -80°C in freezers at the WTBB SJUH until batch processing to RNA after all samples have been collected.

Table 2. Summary of laboratory measurements. For more details please consult Section 9.2.2.5 and the IB

| **Measurement (day)** | **Screening (-7 to -2)** | **Visit 1 (0)** | **Visit 2 (2)** | **Visit 3 (7)** | **Visit 4 (28)** | **Visit 5 (30)** | **Visit 6 (35)** | **Follow-up (>42)** |
|---|---|---|---|---|---|---|---|---|
| Blood HbA1c, lipids, FBC, LFT, eGFR, U+E, adrenal and thyroid (and CK at screening) | X | X | | X | X | | X | X |
| Blood plasma for AZD4017 detection | | | | | X | | | |
| Blood serum and plasma for biobank (2 each) | | X | | | | | X | |
| 24 hour urine samples | | X | | | | | X | |
| Skin biopsies | | X | | | X | | | |

#### 11.11. Other safety measurements

Not applicable

#### 11.12. Annual reports

**Confidential**. Intellectual property: University of Leeds.

An annual report describing the general progress and any relevant safety data related to the trial must be submitted to the Main REC, MHRA and the Sponsor on the anniversary of the Clinical Trial Authorisation being granted. The annual report will follow the format of a Developmental Safety Update Report. A template and guidance for the completion of this report is available from the Sponsor office. The CI must review and sign/date the report. Any findings in the Developmental Safety Update Report that are inconsistent with the IB should be communicated to AstraZeneca during report production but at the latest in parallel to the report being sent to the regulatory authorities. The production of any Developmental Safety Update Report with no new safety concerns should be confirmed in writing to the AstraZeneca operational representative.

##### 11.13. End of trial report

A declaration of end of trial form must be submitted to the MHRA within 90 days of the end of the trial (or 15 days for a premature termination). Upon completing the trial, as defined in section 6.4.4, an end of trial report must be submitted to the MHRA within one year of the end of the trial by the Sponsor or Sponsor-delegated individual. A copy of this end of trial report will also be supplied to all support departments involved in the study, for example pharmacy and or radiology. The CI must review and sign/date the report. AstraZeneca require an unblinded listing of SAE and Suspected Serious Adverse Reactions to enable unblinding of these events on the AZ safety database. For convenience and completeness SUSAR should also be included and easily identifiable. This will be provided to AstraZeneca at 'clean file' (when all study queries have been answered and the database is locked) at the following time points:

1. At primary analysis

2. After last patient has completed study treatment

For all safety reporting communications with AstraZeneca the AstraZeneca reference number ESR-16-12321 and IMP name should be included in email headers and emails should be sent in an encrypted file e.g. WinZip.

#### 12. STUDY MANAGEMENT AND ADMINISTRATION

##### 12.1. Training of study site personnel

All Investigators will have completed GCP training within 2 years prior to study commencement. All procedures included in this study are already conducted at the Leeds sites as part of other on-going studies. Staff contributing to the study will be invited to a launch meeting to be briefed on the study background, objectives, eligibility criteria, procedures and safety protocols.

##### 12.2. Good clinical practice and regulatory compliance

**Confidential**. Intellectual property: University of Leeds.

This clinical trial, which involves the use of an IMP has been designed and will be run in accordance with the Principles of GCP and the current regulatory requirements, as detailed in the Medicines for Human Use (Clinical Trials) Regulations 2004 (UK S.I. 2004/1031) and any subsequent amendments of the clinical trial regulations.

#### 12.3. Adherence to protocol

The Investigators will not deviate from the protocol. In medical emergencies, the CI may use his medical judgment and may remove a study participant from immediate hazard before notifying the Sponsor, the MHRA and the REC in writing regarding the type of emergency and the course of action taken.

#### 12.4. Monitoring, audit and inspection

The Sponsor reserves the right to audit any site involved in the trial and authorisation for this is given via the study contract or agreement. A site may be audited, by LIRMM, an independent contractor working for LIRMM or may be subject to inspection by the MHRA in order to ensure compliance with International Conference on Harmonisation (ICH)-GCP, and the Scientific Lead will allow direct access to trial documentation.

##### 12.4.1. Procedures for monitoring subject compliance

The administration of all study medication (including IMP) will be recorded in the appropriate sections of the CRF.

The administration of all study medication will be recorded directly into the CRF as the expected number of tablets remaining, the actual number of tablets remaining, the overall percentage compliance ((140-(actual number of doses remaining-expected number of tablets remaining))/140)*100 and the cumulative percentage compliance (1-((actual number of tablets remaining-expected number of tablets remaining) / (visit day x 4)))*100 at Visits 2-6. Any missed tablets will be documented in the medical notes, along with reasons for the missed tablets.

Percentage Diary Card completion will also be recorded in the CRF (number of doses recorded divided by the total available number of doses and multiplied by 100) along with a copy of the Diary Card. Diary Card originals will be stored in the TMF.

##### 12.4.2. Definition of source data

Source documents are original records in which raw data are first recorded. These may include, e.g. hospital/clinic/general practitioner records, charts, diaries, laboratory results, printouts, pharmacy records, care records, ECG or other printouts. Source documents will be kept in a secure, limited access area.

Some data will be recorded directly in the CRF and will not appear in a source document as defined in the Source Data Location Sheet (e.g. BMI, BP, waist-hip ratio).

**Confidential**. Intellectual property: University of Leeds.

Source documents that are computer-generated and stored electronically will if possible/practical be printed for review by the monitor. Once printed, these copies will be signed and dated by the Scientific Lead or CI where appropriate and become a permanent part of the subject's source documents.

The Scientific Lead will authorize the monitor to compare the content of the print out and the data stored in the computer to ensure all data are consistent. If electronically stored and impractical to print, each timely review of the electronically-stored data will be annotated in the patient's notes.

###### 12.4.3. Source data verification

Source data verification ensures accuracy and credibility of the data obtained. During monitoring, reported data are reviewed with regard to being accurate, complete, and verifiable from source documents (e.g. subject files, recordings from automated instruments, ECG tracings, laboratory notes). All data reported on the CRF will be supported by source documents, unless otherwise specified in section 12.4.2. Data Verification and will be carried out by the Scientific Lead and members of the study team who will check the CRF for completeness and clarity, and crosscheck them with source documents.

###### 12.4.4. Quality assurance

Investigators will promptly notify the Sponsor Quality Assurance Office of the following within the required timeframe:

- Serious breaches of GCP
- Urgent safety measures
- Protocol violations
- Any amendments to the trial
- Any changes the Clinical Trial Risk Assessment (form A).
- Any other issues as stated in the study contract or agreement

###### 12.4.5. Trial oversight

###### 12.4.6. Data monitoring, ethics and trial steering committee

Independent oversight of the study will be conducted by the Independent Data Monitoring, Ethics and Trial Steering Committee. Amongst its members will be an independent chair, a lay individual (from the Leeds Musculoskeletal Biomedical Research Unit Public and Patient Advocacy Group), a clinician who is independent of the study research team, and a representative of the LIRMM study management group. They are expected to meet at least quarterly. A copy of the Independent Data Monitoring, Ethics and Trial Steering Committee minutes will be forwarded to the Sponsor QA office for review.

The daily running of the trial will be co-ordinated by Dr Tiganescu who will lead the Trial Management Group (also comprising of Dr Ajjan, Dr Del Galdo and Dr Tahrani).

**Confidential**. Intellectual property: University of Leeds.

#### 12.5. Data handling

##### 12.5.1. CRF completion

The research team is responsible for prompt reporting of accurate, complete, and legible data in the CRFs and in all required reports. Any change or correction to the paper CRF will be dated, initialled, and explained (if necessary) and will not obscure the original entry. Use of correction fluid is not permitted. The Scientific Lead will maintain a list of personnel authorized to enter data into the CRF. Detailed instructions will be provided in the CRF Instructions.

##### 12.5.2. Database entry and reconciliation

CRF/external electronic data will be captured on an electronic spreadsheet, accessible only to the CI and delegated personnel and stored on a secure server with regular backups. Periodic manual reviews will be conducted to check for discrepancies and to ensure consistency with CRF/external electronic data. Upon completion, the database will be locked for editing and subjected to final data inspection as recommended by the Data Management Committee before analysis.

##### 12.5.3. Screening and enrolment logs

Subject's Screening will be recorded in the Subject Screening Log.

The Scientific Lead will keep a list containing all subjects enrolled into the study. This list remains with the Scientific Lead and is used for unambiguous identification of each subject. The list contains the subject identification number, full name, date informed consent signed and the hospital number or National Health Security number, if applicable.

The subject's consent and enrolment in the study must be recorded in the subject's medical record. These data will identify the study and document the dates of the subject's participation.

#### 12.6. Archiving and data retention

In line with the principles of GCP/UK Clinical Trial Regulations, at the end of the trial, essential documents will be securely archived at each participating centre for a minimum of 15 years. However, because of international regulatory requirements, the Sponsor may request retention for a longer period. Arrangements for confidential destruction will then be made. **If a patient withdraws consent for their data to be used, it will be confidentially destroyed immediately.** No records/study documentation/data may be destroyed without first obtaining written permission from the Sponsor.

Essential documents include (this list is not exhaustive):

- Signed informed consent documents for all subjects
- Subject identification code list*, screening and enrolment log

**Confidential**. Intellectual property: University of Leeds.

- Record of all communications between the CI, the REC and the Sponsor
- Composition of the REC, and the Sponsor (or other applicable statement as described in section 14.6).
- List of sub-Investigators and other appropriately qualified persons to whom the Scientific Lead/CI has delegated significant trial-related duties, together with their roles in the study and their signatures
- Copies of CRF and documentation of corrections for all subjects
- Investigational product accountability records
- Record of any body fluids or tissue samples retained
- All other source documents (subject medical records, hospital records, laboratory records, etc.)
- All other documents as listed in section 8 of the ICH E6 Guideline for GCP (Essential Documents for the Conduct of a Clinical Trial)

*European Union legislation requires this list to be maintained for a minimum of 15 years.

Normally, these records will be held in the Scientific Lead's archives. If the Scientific Lead is unable to meet this obligation, he or she must ask the Sponsor for permission to make alternative arrangements. Details of these arrangements will be documented.

##### 12.7. Study suspension, termination and completion

Suspension or termination of the study may occur at any time for any reason, following discussion between the Investigators and the Sponsor. In the case of early study termination the Sponsor or Sponsor-delegated individual will be responsible for completing a premature end of study report to the MHRA and the REC within 15 days. Upon study completion, the Sponsor or Sponsor-delegated individual will be responsible for sending the Declaration of the End of a Clinical Trial to the MHRA within 90 days. The Sponsor or Sponsor-delegated individual will be responsible for providing the end of trial report to the MHRA within 1 year of the end of the trial.

#### 13. DATA EVALUATION

##### 13.1. Responsibilities

Data analysis, report preparation and study dissemination will be conducted by the Scientific Lead and other delegated authorised personnel.

##### 13.2. Hypotheses

In this pilot study, analysis will be descriptive throughout and hence no specific inferential hypotheses are being formally tested. We seek preliminary descriptive evidence that:

- Oral AZD4017 inhibits 11β-HSD1 activity in skin
- AZD4017 is safe and well-tolerated in patient with T2DM

**Confidential**. Intellectual property: University of Leeds.

- Oral AZD4017 regulates the skin function outcome measures described in this study
- Systemic GC levels and skin 11β-HSD1 activity, independently or in combination correlate with the skin function outcome measures described in this study

#### 13.3. General statistical considerations

Unless otherwise specified, the last valid measurement before study medication administration will be utilized as the baseline value. In general, summary statistics [n (number of available measurements), arithmetic mean, standard deviation, median, minimum, and maximum] for quantitative variables and absolute and relative frequency tables for qualitative data will be presented.

Wherever possible the trial will be reported in accordance with the recommendations of the Consolidated Standards of Reporting Trials statement.

An initial blind review of the data will be completed prior to locking the database to identify patients to be excluded from per-protocol analysis.

#### 13.4. Planned analyses

##### 13.4.1. Primary endpoint analysis

Skin 11β-HSD1 activity will be summarized descriptively in each treatment group at each time-point; the primary analysis will be on an intention-to-treat basis, will all patients included, as randomised. This will be supplemented by a per-protocol analysis that includes patients whose compliance with protocol is deemed satisfactory. Unadjusted and adjusted summaries (adjusted for gender, age and baseline HbA1c) for absolute values and changes from baseline will be presented; between-group differences in absolute values and changes from baseline will be presented together with 90% confidence intervals – deemed acceptable for pilot studies [65]. Adjusted summaries will be obtained via a linear regression model which mirrors analysis of covariance. Where necessary, transformations to a normal distribution will be performed. Preliminary proof-of-concept will be considered to have been achieved if adjusted mean 11β-HSD1 activity in skin at 28 days is lower in the active treatment arm compared to the placebo arm. Estimated sample sizes for future trials will be produced based on the pooled standard deviations from both treatment arms for the following outcomes: 11β-HSD1 activity in skin (at baseline and 28 days), sudomotor function, skin hydration, epidermal barrier function (at baseline and 35 days), integrity (at baseline and 28 days) and recovery (at baseline and 28 days – 3 hour, 2 day and 7 day), skin thickness (at baseline and 35 days) and wound healing (at baseline and 28 days – 2 day and 7 day). Sample sizes for a range of plausible and clinically-meaningful between-group differences will be presented, assuming analysis of covariance controlling for baseline values would be used. Correlation (Pearson's r) between baseline and follow-up values will be estimated in the combined treatment groups to aid in the sample size estimates. If the

**Confidential**. Intellectual property: University of Leeds.

correlation between the measurements at the two time-points is ρ, ANCOVA comparing groups of $(1-\rho^2)n$ subjects has the same power as a t-test comparing groups of n subjects.

##### 13.4.2. Secondary endpoint analyses

###### 13.4.2.1. Efficacy

Systemic 11β-HSD1 activity and skin function analyses will be conducted as for the primary endpoint (above). The strength of association between skin AZD4017 at day 28 concentration and plasma AZD4017 at day 35 in the active treatment arm will be assessed using Pearson's product moment correlation coefficient; for severely skewed variables for which a suitable transformation cannot be found, Spearman rank correlation will be used. For all correlation analyses, absolute correlation coefficients with a value of r(ho)≥0.3 will be considered preliminary evidence of substantive association.

###### 13.4.2.2. Safety

Safety analyses will be conducted in all patients who received any study treatment, according to treatment received. Continuous safety variables (BMI, waist-hip ratio, BP, HbA1c levels, lipids, FBC, LFT, eGFR, U+E, adrenal and thyroid function test) will be summarized descriptively by patient group at each timepoint. Unadjusted and adjusted summaries (adjusted for gender, age and baseline HbA1c) for absolute values and changes from baseline will be presented; between-group differences in absolute values and changes from baseline will be presented together with 90% confidence intervals. Adjusted summaries will be obtained via a linear regression model which mirrors analysis of covariance. Where necessary, transformations to a normal distribution will be performed. Where applicable, numbers of patients in each group whose values exceeded ULN (or were below the lower limit of normal) during the trial will be presented, corrected for exposure by 100 patient years.

Line listings of all AEs will be provided in the end of trial report. The frequency of all AE during the study period will be presented for each treatment group separately within classes of AEs defined by CTCAE preferred term. The data will be displayed as number of subjects experiencing the AE, percentage of subjects, and number of AE. AE will also be summarized by severity and relation to IMP. For recurrent AEs, the most severe occurrence will be recorded per AE per patient. AE data will also be corrected for exposure by 100 patient-years. For expected AEs the recorded rates will be compared against the rates outlined in the IB.

In addition to the above, the proportions of patients who passed the overall assessment of blood safety will be summarised using absolute and relative frequencies at each visit, in all patients and by treatment group at days 0, 7, 28, 35 & 42.

No inferential testing will be conducted; safety will be assessed in terms of the clinical relevance of the AEs reported and of any observed differences between groups.

**Confidential**. Intellectual property: University of Leeds.

###### 13.4.3. Skin function

The strengths of associations among systemic GC level, skin 11β-HSD1 activity and skin outcome measures will be assessed using Pearson's product moment correlation or, where a bivariate association needs to adjust for a third variable, partial correlation. For severely skewed variables for which a suitable transformation cannot be found, Spearman rank correlation will be used.

###### 13.4.4. Feasibility

Feasibility variables will be summarised descriptively in all patients. They will be interpreted in terms of whether sufficient data were obtained to determine feasibility and if so, whether the data obtained indicate that a larger phase II trial is feasible and what adjustments to protocol, if any, may be needed.

##### 13.5. Safety analyses

See section 13.4.2.2

##### 13.6. Definition of 'per protocol' set

In addition to an intention-to-treat analysis including all patients, a supplemental 'per protocol' analysis (including only those who adhered to the treatment regimen of the programme they were allocated, did not violate the trial protocol in any substantial way, and have data available) will be performed. Prior to unblinding of the study, patients will be allocated to or excluded from the 'per protocol' set by a panel including the clinical project manager, trial statistician, and other appropriate clinical study team members.

##### 13.7. Handling of dropouts and missing data

The level of missing data is one of the feasibility endpoints of this trial. Effective means of imputation such as multiple imputation may prove impractical if the sample size is too small for the imputation model to converge successfully. If multiple imputation cannot be performed, available case analysis will be used, supplemented by last-observation-carried forward single imputation as a sensitivity analysis. Multiple imputation by chained equations will be attempted, using multiple regression as the imputation model. If multiple imputation is viable, both available case and LOCF will be performed as sensitivity analyses.

##### 13.8. Planned interim analysis and data monitoring

In this small study with short follow-up no formal interim efficacy analysis is planned. Data monitoring will be carried out during the trial by the study management team and the sponsor (see section 12.4). . Interim safety monitoring will be conducted by the study Data Management and Ethics Committee. The DMEC independent statistician will obtain the randomisation list from pharmacy and will code this as treatment A or B before passing to the trial statistician. The trial statistician will produce both open (blinded) and closed

**Confidential**. Intellectual property: University of Leeds.

(unblinded) reports for the DMEC, the latter using the treatment codes. Only safety data values will be presented by treatment group; data completeness will be summarised for all secondary variables and for biopsy success. Feasibility data will also be presented. The primary outcome will not be available for interim analysis as this will not be calculated until after the final patient completes follow-up. The trial statistician will not be present when closed reports are discussed by the DMEC with knowledge of the codebreak, which will be supplied to committee members by the DMEC independent statistician. The DMEC will examine at least one set of interim safety data after the first 8 patients have completed the trial up to day 35 (or have been withdrawn); timings of subsequent analyses will be informed by the findings of the first analysis.

There will be a blind review of the data after the last patient has completed follow-up.

#### 13.9. Determination of sample size and randomization method

Sample size is small as this is a preliminary pilot study and the first use in this patient group. Published rules of thumb for pilot studies recommend a sample size of 12 participants per arm completing the protocol [66]. To ensure this minimum number complete the protocol we will randomise 15 per arm to allow for 20% drop-out.

#### 13.10. Procedure for unblinding the study prior to analysis

The final database is not to be locked or unblinded until a review of the data has been completed, the per protocol sample identified, and the statistical analysis plan has been finalised. The dates of each of these milestones will be recorded in the study documentation prior to the full unblinding of the data.

### 14. ETHICS AND REGULATORY REQUIREMENTS

#### 14.1. Good Clinical Practice

This study will be conducted in accordance with applicable laws and regulations including, but not limited to, the ICH Guideline for GCP and the recommendations guiding ethical research involving human subjects adopted by the 18th World Medical Assembly, Helsinki, Finland, 1964, amended at the 48$^{th}$ General Assembly, Somerset West Republic of South Africa, October 1996. The REC and MHRA must review and approve the protocol and Informed Consent Form before any subjects are enrolled. Before any protocol-required procedures are performed, the subject must sign and date the REC-approved Informed Consent Form. The right of a patient to refuse participation without giving reasons must be respected. The patient must remain free to withdraw at any time from the study without giving reasons and without prejudicing his/her further treatment. The study will be submitted to and approved by a Main REC and the appropriate regulatory authorities prior to entering patients into the study.

**Confidential**. Intellectual property: University of Leeds.

#### 14.2. Delegation of Investigator duties

The Investigators will ensure that all persons assisting with the trial are adequately qualified and informed about the protocol, any amendments to the protocol, the study treatments, and their trial-related duties and functions.

The Scientific Lead will maintain a delegation log of co-Investigators and other appropriately qualified persons to whom he or she has delegated significant trial-related duties.

#### 14.3. Subject information and informed consent

Before being enrolled in the clinical study, subjects must consent to participate after the nature, scope, and possible consequences of the clinical study have been explained in a form understandable to them.

A PIL that includes information about the study will be prepared and given to the subject at least 24 hours prior to the screening visit. This document will contain all the elements required by the ICH E6 Guideline for GCP and any additional elements required by local regulations. The document must be translated (by an independent interpreter) into a language understandable to the subject and must specify who informed the subject. Where required by local law, the person who informs the subject must be a physician.

During verification of eligibility, patients will be given the opportunity to ask questions and the nature and objectives of the study will be explained. A research nurse may help in this process but the study doctor is responsible for the General Informed Consent discussions at the Screening Visit. The Skin Biopsy Informed Consent at Visits 1 and 4 can be received by a qualified member of staff who has signed/dated the staff delegation log (i.e. study doctor or research nurse).

After reading the informed consent document, the subject must give consent in writing. The subject's consent must be confirmed at the time of consent by the personally dated signature of the subject and by the personally dated signature of the person conducting the informed consent discussions, (i.e. the study doctor for General Informed Consent or qualified research nurse for Skin Biopsy Informed Consent).

The original signed Informed Consent Form will be retained in the TMF. Other copies of the consent form are required:

- One copy will be kept in the patient’s clinical notes
- One copy will be given to the patient

Consent is an ongoing process and will be reassessed at each study visit.

The Scientific Lead will not undertake any measures specifically required only for the clinical study until valid consent has been obtained.

The CI must inform the subject’s GP about the subject’s participation in the trial if the subject has a GP and if the subject agrees to the GP being informed.

**Confidential**. Intellectual property: University of Leeds.

#### 14.4. Subject confidentiality

Only the subject enrolment number will be recorded in the CRF, and if the subject name appears on any other document (e.g. laboratory report), it must be obliterated on the copy of the document to be supplied to anyone outside the clinical care team. The subjects will be informed that representatives of the Sponsor, REC or regulatory authorities may inspect their medical records to verify the information collected, and that all personal information made available for inspection will be handled in strictest confidence.

All information collected during the course of the trial will be kept strictly confidential.

Information will be held securely on paper and electronically.

The Diabetes Clinic at SJUH and LMBRU at CAH will comply with all aspects of the Data Protection Act 1998.

The Scientific Lead will maintain a personal subject identification list (subject numbers with the corresponding subject names) to enable records to be identified.

#### 14.5. Subject identification cards

All patients who receive at least one dose of study medication will be issued with identification cards which state the name of the IMP and an indication of the possibility that the patient may be receiving placebo or control treatment, in addition to details of whom to contact in the event of an emergency.

#### 14.6. Approval of clinical study protocol and amendments

Before the start of the study, the clinical study protocol, informed consent document, and any other appropriate documents will be submitted to the REC, the MHRA and the Sponsor with a cover letter or a form listing the documents submitted, their dates of issue, and the site (or region or area of jurisdiction, as applicable) for which approval is sought. If applicable, the documents will also be submitted to the authorities, in accordance with local legal requirements.

Investigational products can only be supplied to the Sponsor after documentation on all ethical and legal requirements for starting the study has been received by the product provider.

Before the first subject is enrolled in the study, all ethical and legal requirements must be met, including approval of the study by the NHS, the Sponsor Research and Development department, the REC and the MHRA.

Amendments must be evaluated to determine whether formal approval must be sought and whether the informed consent document should be revised, thus all protocol amendments and administrative changes must first be discussed with and approved by the Sponsor before being submitted to the REC and the MHRA, in accordance with legal requirements.

**Confidential**. Intellectual property: University of Leeds.

The Scientific Lead must keep a record of all communication with the REC, the MHRA, and the Sponsor. This also applies to any communication between the Scientific Lead and the authorities.

##### 14.7. Protocol amendments

Requests for any amendments to the study must be sent to the Sponsor by the Scientific Lead. The Sponsor will determine whether said amendments are substantial or non-substantial prior to their submission to the appropriate bodies for approval. Patients will be re-consented to the study if the amendments affect the information they have received, patient safety, or if the change alters the type or quality of the data collected for the study. Patients will only be re-consented AFTER an amendment has been fully approved.

##### 14.8. Ongoing information for MHRA/REC

Unless otherwise instructed by the MHRA, REC and the Sponsor, the Scientific Lead must submit to the MHRA, REC and the Sponsor:

- Information on SUSAR from the CI's site, as soon as possible and within 24 hours (one business day) of the research team becoming aware of them
- Expedited safety reports, as soon as possible
- Annual reports on the progress of the study
- The Declaration of the End of a Trial form

#### 15. FINANCE AND INSURANCE

##### 15.1. Indemnity and insurance

The University of Leeds is able to provide insurance to cover for liabilities and prospective liabilities arising from negligent harm. In certain circumstances we provide insurance cover for claims arising from non-negligent harm. Clinical negligence indemnification will rest with the participating NHS Trust or Trusts under standard NHS arrangements.

Further details of liability and insurance provisions for this study are given in separate agreements.

##### 15.2. Financial disclosure

None of the Investigators or members of the research team have any financial involvement with the sponsorship or funding bodies or will receive personal benefits, incentives or payment over and above normal salary.

#### 16. PUBLICATION

**Confidential**. Intellectual property: University of Leeds.

According to the GC-SHealD Research Collaboration Agreement with AstraZeneca, the University of Leeds and its employees, students and agents shall be entitled to publish the results of, or make presentations related to, this study to the extent that such publications or presentations are consistent with academic standards, are not false or misleading, and are not for commercial purposes, subject to the following:

The University of Leeds shall provide, or shall ensure that the Scientific Lead provides, AstraZeneca with copies of any materials relating to the study, study data or the developed technologies that it either intends to publish (or submit for publication, including, but not limited to, materials to be posted on clinical trial registries) or make any presentations relating to, at least forty five days in advance of publication, submission or presentation.

**Confidential**. Intellectual property: University of Leeds.

**Confidential**. Intellectual property: University of Leeds.

**Confidential**. Intellectual property: University of Leeds.

**Confidential**. Intellectual property: University of Leeds.

32. Rosenstock, J., et al., *The 11-beta-hydroxysteroid dehydrogenase type 1 inhibitor INCB13739 improves hyperglycemia in patients with type 2 diabetes inadequately controlled by metformin monotherapy.* Diabetes Care, 2010. **33**(7): p. 1516-22.
33. Feig, P.U., et al., *Effects of an 11beta-hydroxysteroid dehydrogenase type 1 inhibitor, MK-0916, in patients with type 2 diabetes mellitus and metabolic syndrome.* Diabetes Obes Metab, 2011. **13**(6): p. 498-504.
34. Shah, S., et al., *Efficacy and safety of the selective 11beta-HSD-1 inhibitors MK-0736 and MK-0916 in overweight and obese patients with hypertension.* J Am Soc Hypertens, 2011. **5**(3): p. 166-76.
35. Syngle, A., et al., *Minocycline improves peripheral and autonomic neuropathy in type 2 diabetes: MIND study.* Neurol Sci, 2014. **35**(7): p. 1067-73.
36. Roh, N.K., et al., *A Split-Face Study of the Effects of a Stabilized Hyaluronic Acid-Based Gel of Nonanimal Origin for Facial Skin Rejuvenation Using a Stamp-Type Multineedle Injector: A Randomized Clinical Trial.* Plast Reconstr Surg, 2016. **137**(3): p. 809-16.
37. Wanitphakdeedecha, R., S. Eimpunth, and W. Manuskiatti, *The effects of mucopolysaccharide polysulphate on hydration and elasticity of human skin.* Dermatol Res Pract, 2011. **2011**: p. 807906.
38. Kennedy, W.R., et al., *A randomized, controlled, open-label study of the long-term effects of NGX-4010, a high-concentration capsaicin patch, on epidermal nerve fiber density and sensory function in healthy volunteers.* J Pain, 2010. **11**(6): p. 579-87.
39. Gibbons, C.H., N. Wang, and R. Freeman, *Capsaicin induces degeneration of cutaneous autonomic nerve fibers.* Ann Neurol, 2010. **68**(6): p. 888-98.
40. Boyd, A.L., et al., *Topiramate improves neurovascular function, epidermal nerve fiber morphology, and metabolism in patients*

**Confidential**. Intellectual property: University of Leeds.

*with type 2 diabetes mellitus.* Diabetes Metab Syndr Obes, 2010. **3**: p. 431-7.
41. Casellini, C.M., et al., *Sudoscan, a noninvasive tool for detecting diabetic small fiber neuropathy and autonomic dysfunction.* Diabetes Technol Ther, 2013. **15**(11): p. 948-53.
42. Seite, S., et al., *Importance of treatment of skin xerosis in diabetes.* J Eur Acad Dermatol Venereol, 2011. **25**(5): p. 607-9.
43. Tahrani, A.A., et al., *Cutaneous structural and biochemical correlates of foot complications in high-risk diabetes.* Diabetes Care, 2012. **35**(9): p. 1913-8.
44. Dinh, T., et al., *Mechanisms involved in the development and healing of diabetic foot ulceration.* Diabetes, 2012. **61**(11): p. 2937-47.
45. Jiang, Y., et al., *A cohort study of diabetic patients and diabetic foot ulceration patients in China.* Wound Repair Regen, 2015. **23**(2): p. 222-30.
46. Gershater, M.A., et al., *Complexity of factors related to outcome of neuropathic and neuroischaemic/ischaemic diabetic foot ulcers: a cohort study.* Diabetologia, 2009. **52**(3): p. 398-407.
47. Prompers, L., et al., *Prediction of outcome in individuals with diabetic foot ulcers: focus on the differences between individuals with and without peripheral arterial disease. The EURODIALE Study.* Diabetologia, 2008. **51**(5): p. 747-55.
48. Vinik, A.I., M.L. Nevoret, and C. Casellini, *The New Age of Sudomotor Function Testing: A Sensitive and Specific Biomarker for Diagnosis, Estimation of Severity, Monitoring Progression, and Regression in Response to Intervention.* Front Endocrinol (Lausanne), 2015. **6**: p. 94.
49. Kim, H.J., et al., *Pseudoceramide-containing physiological lipid mixture reduces adverse effects of topical steroids.* Allergy Asthma Immunol Res, 2011. **3**(2): p. 96-102.
50. Sheu, H.M., et al., *Depletion of stratum corneum intercellular lipid lamellae and barrier function abnormalities after long-*

**Confidential**. Intellectual property: University of Leeds.

*term topical corticosteroids.* Br J Dermatol, 1997. **136**(6): p. 884-90.

**Confidential**. Intellectual property: University of Leeds.

**Confidential**. Intellectual property: University of Leeds.
