## Appendix 3 for "A randomised controlled pilot trial of oral 11β-HSD1 inhibitor AZD4017 for wound healing in adults with type 2 diabetes mellitus"

### APPENDIX 3 – Statistical Methods

#### 1.1 Primary outcome analysis

For the primary variable (24 hour 11β-HSD1 activity in skin at day 28) unadjusted and adjusted summaries (adjusted for gender, age and baseline 11β-HSD1 activity and HbA1c) for final values and changes from baseline are presented; between-group differences in final values and changes from baseline were presented together with 90% confidence intervals, supplemented with confidence intervals ranging from 75%-95% in 5% increments as pre-specified in the SAP. If the adjusted difference was in favour of the intervention arm, i.e. if 11β-HSD1 activity was lower, this was to be considered preliminary evidence of efficacy.

Because model residuals were non-normally distributed and a suitable data transformation could not be found, quantile (median) regression was used to obtain adjusted summaries and CIs, as pre-specified in the SAP.

The primary analysis was on an intention-to-treat basis, with all patients included, as randomised. Analysis was conducted in the full analysis set using multiple imputation by chained equations to address missing data. Five-nearest-neighbour predictive mean matching was used for all variables; for each outcome the imputation model included the repeated observations of the outcome, treatment assignment, age, sex, baseline HbA1c and overall IMP compliance. Variables which were correlated with the outcome or the likelihood of missingness, and which had fewer missing values at a given time-point, were also included. Twenty datasets were imputed, inspection of Monte Carlo errors indicated this was sufficient. Estimates were combined according to Rubin’s rules. Sensitivity analyses using available case and last observation carried forward were also performed. A planned sensitivity analysis in the per protocol set was not performed (see section 1.7 below).

#### 1.2 Secondary outcome analysis

The analyses for the secondary endpoints, systemic 11β-HSD1 activity and skin function, and continuous clinical laboratory safety variables were conducted as for the primary endpoint (above). Adjusted summaries were obtained via a linear regression model that mirrored the analysis of covariance approach for wound healing and laboratory safety variables. For all other variables visual inspection of linear regression model residuals indicated they were substantively non-normally distributed. For TEWL and epidermal integrity, log-transformation was performed prior to linear regression. For the remaining outcomes, quantile (median) regression was used.

Planned supplementary analyses used linear mixed modelling to allow the precise timing of measurements to be included as a covariate where relevant, to account for any differences in timings between groups. Likelihood ratio tests supported the inclusion of non-linear (quadratic) terms for change over time, and allowing changes over time to vary between patients.

#### 1.3 Additional analyses

Correlations between plasma AZD4017 concentration at day 35 and skin AZD4017 concentration at day 28, and between AZD4017 compliance and efficacy outcomes in the active treatment arm were estimated using Spearman rank correlation.

The strengths of associations among skin 11β-HSD1 activity and skin outcome measures controlling for systemic GC level were assessed using partial correlation following rank transformation. Correlation coefficients were transformed using Fisher’s z transformation prior to averaging across multiple imputed datasets. For all correlation analyses, absolute correlation coefficients with a value of r(ho)≥0.3 were considered preliminary evidence of substantive association.

The numbers of patients with clinical laboratory values below, within, or above normal ranges, pre-intervention versus each post-intervention time-point were tabulated for each test, for the Safety Population by treatment group. The proportions of patients who passed the overall assessment of blood safety at days 0, 7, 28, 35 & 42 were summarised.

For adverse events, summaries of incidence rates (frequencies and percentages), intensity, and relationship to study drug of individual AEs by System Organ Class and Preferred Term (CTCAE) were presented.

Feasibility variables were summarised descriptively.

Estimated sample sizes for future trials were produced based on the pooled standard deviations from both treatment arms for the following outcomes: 11β-HSD1 activity in skin (at 28 days), sudomotor function, skin hydration, epidermal barrier function, integrity (at 28 days) and recovery (at 28 days – 3 hour, 2 day and 7 day), skin thickness (at 35 days) and WH (at 28 days – 2 day and 7 day).

#### 1.4 Changes in the Conduct of the Study or Planned Analyses

##### Recruitment halted early

Study recruitment was intended to continue until a total of 30 patients had been randomised. This covered the 12 per group recommended for pilot studies and allowed for 20% drop-out. The recruitment period was extended twice; at the end of the second extension 28 patients had been recruited and the drop-out rate was found to be low (<5%), therefore we decided to halt recruitment rather than extend it further, risking delays to the reporting of the current trial and to the planning of future trials based on these results. This decision was made without reference to the primary outcome measure, which had not yet been processed, and prior to the breaking of the blind.

##### Primary outcome unit of measurement

Following the completion of the study and final database lock, the primary outcome was found to have been calculated as % conversion per *24 hours*, rather than *per hour* as stipulated in the protocol. As this was a matter of direct conversion and would not affect the conclusions, it was agreed with the Sponsor that the values would remain as they were and would be reported as % conversion per 24 hours.

##### Validation of primary outcome by ELISA

In the protocol amendment to v2.0, validation of the radioimmunoassay method of measuring 11β-HSD1 activity in the skin was added in the form of a cortisol Enzyme Linked Immunosorbent Assay (ELISA). This was added prior to the processing of the biopsy samples and prior to the breaking of the blind.

##### Measurement of wound depth instead of diameter at days 7 and 35

At 2 days post-wounding, maximal early granulation tissue width (a marker of early healing) was pre-specified as the standardised indicator of wound diameter. However, this had fully resolved by 7 days post-wounding. Therefore, maximal clot depth (a marker of later healing which was absent at 2 days post-wounding) was substituted as the standardised indicator of healing at this time point

##### Per protocol analysis

Only one patient was excluded from the per protocol set; this patient had withdrawn from follow-up due to work commitments after day 7. Because the per protocol analysis was intended to include only patients in the per protocol set with data available, on an outcome by outcome basis, this effectively meant that the per protocol analysis was identical to the planned available case sensitivity analysis.

##### Additional sensitivity analysis of multiply imputed data

An additional planned sensitivity analysis which would have increased or decreased imputed values in multiples of the baseline standard deviation in the observed data was not performed due to the overall low level of missing data.

#### 1.5 Analysis populations

##### Safety Population (SAFETY)

The Safety Population includes all patients who received any amount of planned study medication.

##### Full Analysis Set (EFFICACY)

The Efficacy Population includes all patients who were randomized and received at least one dose of planned study medication.

##### Per-Protocol Efficacy (PP-EFFICACY)

The PP-Efficacy includes all patients in the Efficacy Population excluding those who met the following criteria:

- Patients receiving prohibited prior and/or concomitant medications
- Patients not meeting the following inclusion / meeting exclusion criteria (i.e. those entered into the study in error)
- Patients whose overall compliance with study treatment during the course of the trial was <80%
- Patients who did not receive the study treatment to which they were randomised
- Patients who withdrew from study treatment for any reason

The above criteria defined the overall per protocol set; in addition, for each primary and secondary variable, at each visit, only patients with data available were included.
