## Supplementary material for "A randomised controlled pilot trial of oral 11β-HSD1 inhibitor AZD4017 for wound healing in adults with type 2 diabetes mellitus": CONSORT extension for Pilot and Feasibility Trials Abstracts Checklist

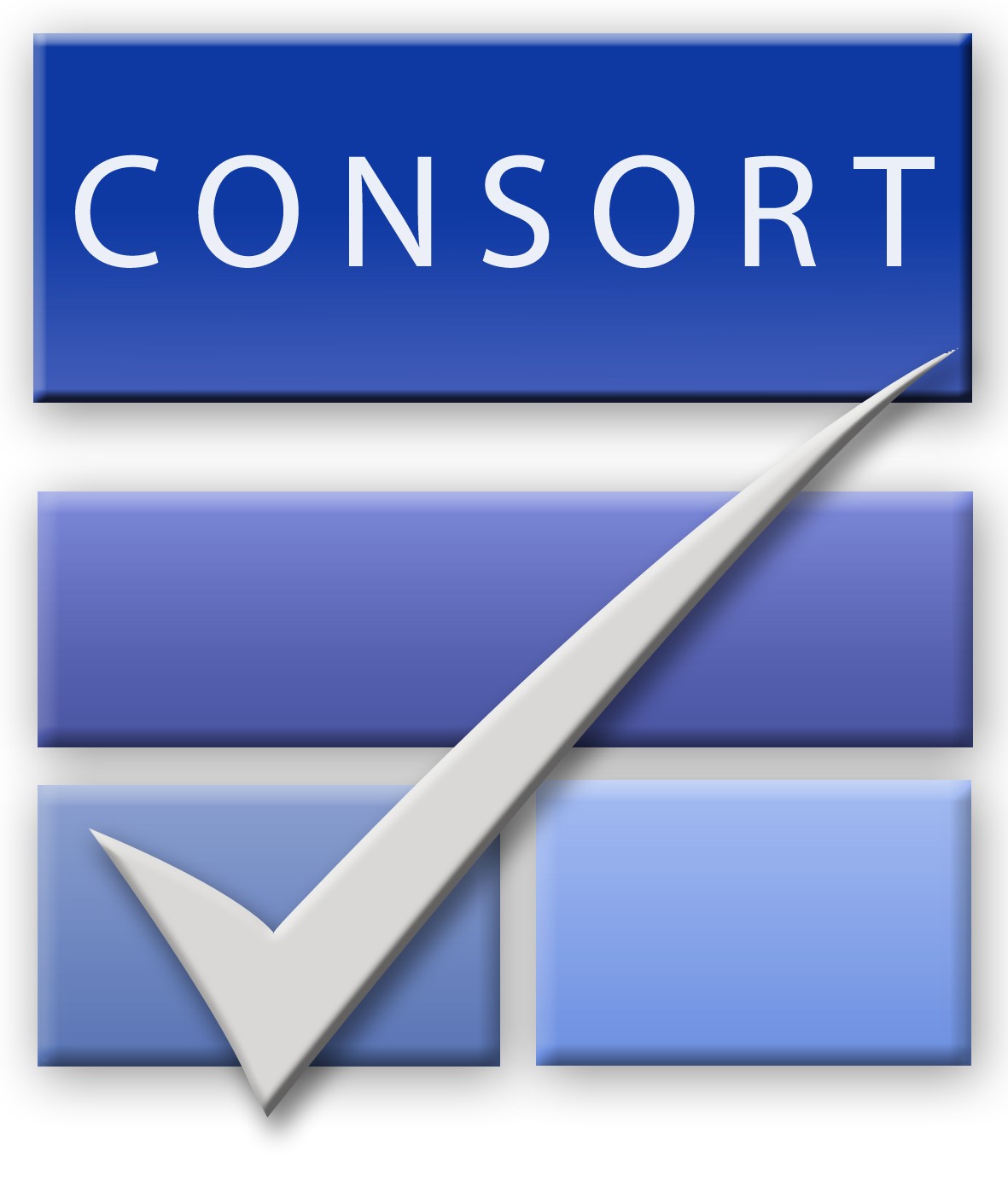
**CONSORT 2010 checklist of information to include when reporting a pilot or feasibility randomized trial in a journal or conference abstract**

| **Item** | **Description** | **Reported on line number** |
| --- | --- | --- |
| Title | Identification of study as randomised pilot or feasibility trial | 45 |
| Authors * | Contact details for the corresponding author | N/A |
| Trial design | Description of pilot trial design (eg, parallel, cluster) | 45 |
| Methods |  |  |
| Participants | Eligibility criteria for participants and the settings where the pilot trial was conducted | 46-47 |
| Interventions | Interventions intended for each group | 46-47 |
| Objective | Specific objectives of the pilot trial | 45 |
| Outcome | Prespecified assessment or measurement to address the pilot trial objectives** | 49-55 |
| Randomization | How participants were allocated to interventions | 48 |
| Blinding (masking) | Whether or not participants, care givers, and those assessing the outcomes were blinded to group assignment | 45 |
| Results |  |  |
| Numbers randomized | Number of participants screened and randomised to each group for the pilot trial objectives** | 48-49 |
| Recruitment | Trial status† | N/A |
| Numbers analysed | Number of participants analysed in each group for the pilot objectives** | 46-47 |
| Outcome | Results for the pilot objectives, including any expressions of uncertainty** | 50-65 |
| Harms | Important adverse events or side effects | 55-56 |
| Conclusions | General interpretation of the results of pilot trial and their implications for the future definitive trial | 57-59 |
| Trial registration | Registration number for pilot trial and name of trial register | 59 |
| Funding | Source of funding for pilot trial | 60 |

Citation: Eldridge SM, Chan CL, Campbell MJ, Bond CM, Hopewell S, Thabane L, et al. CONSORT 2010 statement: extension to randomised pilot and feasibility trials. BMJ. 2016;355.

**this item is specific to conference abstracts*

***Space permitting, list all pilot trial objectives and give the results for each. Otherwise, report those that are a priori agreed as the most important to the decision to proceed with the future*

*definitive RCT.*

*†For conference abstracts.*
